# Trajectories of physical activity components among community-dwelling older adults

**DOI:** 10.64898/2026.04.10.26350593

**Authors:** Bram Hoogerheide, Esther T. Maas, Marjolein Visser, Trynke Hoekstra, Laura A. Schaap

## Abstract

**Background/Objective:** Common measures of physical activity (PA) based on duration and intensity do not fully capture its complexity. Adding additional PA components of muscle strength, mechanical strain, and turning actions, can provide a more complete view of activity behavior. Furthermore, PA behaviors differ between men and women. Therefore, the goal of this study is to identify and cluster similar long-term PA patterns over time for each PA component, examined separately for men and women.

**Methods:** We used data from 4963 participants (52% women; mean age 66 years, SD = 8.6) of the Longitudinal Aging Study Amsterdam (1992–2019). PA component scores were assigned to self-reported activities, and Sequence Analysis with Optimal Matching was used to identify and cluster similar activity patterns over a period of 10 years, separately for each component and stratified by sex.

**Results:** PA components varied by sex and displayed a unique mix of trajectories, including predominately low, medium, or high activity, increasing or decreasing patterns, and trajectories characterized by early or late mortality. Importantly, trajectories remained independent, indicating that changes in one PA component were not linked to changes in others.

**Conclusion:** Older men and women follow distinct and independent long-term PA trajectories across components, underscoring that PA behavior cannot be described by a single dimension.

**Significance/Implications:** The observed independence and heterogeneity of trajectories suggest that muscle strength, mechanical strain, and turning actions capture meaningful and distinct aspects of PA that are not reflected by traditional measures alone. Future PA-strategies could incorporate these dimensions and acknowledge sex-specific patterns to better reflect natural movement. The independence of components suggests that future interventions should target multiple dimensions, as changes in one component may not translate to others. Such an approach may support more tailored and sustainable PA interventions in later life.

## 1. Introduction

### 1.1. Background

Physical activity (PA) levels tend to decline significantly with age [1,2]. This reduction in PA levels is concerning, as lower levels of PA are associated with a variety of adverse health conditions [2,3], worsening mental health [4], and cognitive decline [3,5,6]. Lower PA levels also contribute to decreased mobility [7,8] and deteriorating bone health [9], which heightens the risk of falls [3] and fractures [10], and subsequently decreases functional independence [3,8].

The most common measures to express PA levels in research are duration and intensity. However, these measures do not adequately take the more nuanced aspects of PA into account that can have both beneficial and potentially harmful effects on older individuals. For example, the high weight-bearing forces during tennis are associated with beneficial effects such as maintaining a sufficient bone density [11,12], preserving muscle strength [12] and reducing cardiovascular risk [12], but also with adverse effects such as increased risk of osteoarthritis (OA) [12,13] and heightened fall risk [14]. These contrasting associations demonstrate that the relationship between a specific activity and health conditions is complex and multifaceted, and cannot fully be captured by the common measurements (i.e., duration and intensity) for PA. This idea is further supported by recent findings that different types of PA also show different dose-response relations with mortality risk [15].

A promising approach to address the complexity of PA measurement was developed by Verweij et al.[16] and captured the PA components of intensity, muscle strength, mechanical strain, and turning actions. These four components were summarized into one scoring instrument, where each type of PA gets a score on all four components, based on data from the Longitudinal Aging Study Amsterdam (LASA) [17–20]. In a follow-up study within the same cohort, these four PA component scores were shown to have distinct associations with onset of OA, with high mechanical strain being a strong risk factor, while high muscle strength scores acted protectively [21], a finding in line with the literature [22,23]. Scoring individual physical activities on multiple components allows for better comparison of different types of PA and their expected health benefits, without stratification for each type of PA. However, the PA component scores have not yet been widely applied and their untapped potential remains.

The few existing studies that have incorporated these PA components have assessed them only cross-sectionally at baseline [16,21]. As a result, it remains unclear how these specific PA components change over time in older adults, and to what extent such changes differ between individuals and components. Studying long-term trajectories of PA components is essential for understanding how PA behavior evolves differently for different individuals during aging, beyond average trends or single time-point assessments. A trajectory-based approach can identify stable patterns, gradual changes, and pivotal points such as sudden declines or other shifts in PA behavior. These pivotal points may reflect emerging health problems, life events, or changing circumstances and could represent windows for timely intervention or even prevention of unfavorable changes. Insight into these dynamic and heterogeneous patterns is highly relevant for public health, as it supports the development of more realistic and sustainable PA strategies that align with how individual older adults actually move in daily life. Moreover, while sex differences in types of PA are well documented [24], little is known about whether men and women differ in the natural change in specific PA components over time. Aging processes differ between men and women, with women experiencing more years in poor health than men [25], which may translate into different PA component trajectories. Understanding these sex-specific patterns is therefore crucial for designing tailored and sustainable PA interventions in order to support healthy aging across the life course.

#### Aims and Objectives

The broader aim is to advance physical activity research by providing a more detailed and dynamic description of activity behavior in older adults. By adopting a component-specific, trajectory-based approach, this study seeks to contribute evidence that can ultimately support the development of better targeted and sex-sensitive public health guidelines and interventions that better reflect how older adults are physically active in daily life. Therefore, the goal of this study is to identify and cluster similar long-term PA patterns over time for each PA component, examined separately for men and women.

## 2. Methods

### 2.1. Data and Sample

We used data from the Longitudinal Aging Study Amsterdam (LASA; www.lasa-vu.nl), an ongoing population-based longitudinal study of adults aged 55+ in the Netherlands [17–20]. The first cohort was recruited in 1992/93 and comprised 3107 respondents aged 55–84 years. In 2002 and 2012, two additional cohorts of 55–64 year olds were added with 1002 and 1023 respondents, respectively, from the same sampling frame. Data were collected through face-to-face interviews and self-completed questionnaires with follow-up measurement waves conducted every three years. All procedures of LASA were in accordance with the ethical standards of the institutional and/or national research committee and with the 1964 Helsinki declaration and its later amendments or comparable ethical standards. All participants of the LASA study provided written informed consent.

### 2.2. Inclusion and exclusion criteria

The present study included all respondents from all the measurement waves from 1992/1993 up to 2018/2019, resulting in a total sample size of 5132 respondents. Respondents were excluded if they had no data on the LASA Physical Activity Questionnaire (LAPAQ) at any measurement wave, resulting in a study sample of 4963. The number of respondents varied per follow-up wave due to drop-outs, refusals, and mortality (see Appendix A).

### 2.3. Measurements

#### 2.3.1. Physical Activity (PA)

Information on PA has been obtained during the main interview of each wave using the LASA Physical Activity Questionnaire (LAPAQ). The LAPAQ is based on the questionnaires by Voorrips et al. [26] and Caspersen et al.[27], and has been validated for the LASA population [28]. Respondents were asked how often and for how long they performed each of the following activities in the last two weeks: walking outdoors, biking, gardening, light household activities, heavy household activities. Respondents could also report up to two of their most frequently performed sports activities.

Following the scoring instrument of Verweij et al.[16] (see also Appendix B), each activity in the LAPAQ was assigned a score for the PA components of *muscle strength*, *intensity*, *mechanical strain*, and *turning actions* of the extremities. Gardening was excluded from the calculation of these PA components [16]. Average duration of each reported activity per day was calculated and added as the fifth PA component of *duration*. Calculation and interpretation of the PA components is reported in Table 1. For each of the PA components, all corresponding scores were summarized into one mean score per respondent, resulting into one average score for each PA component.

**Table 1.** Overview of Physical Activity (PA) components and their operationalization.

| Component | Range * | Unit of measurement | Calculation |
| --- | --- | --- | --- |
| **Duration** | 1-960 | Average minutes per day spent on all types of PA combined. Duration is directly derived from information in the self-reported LASA PA Questionnaire (LAPAQ). | Frequency multiplied with average duration of each activity in the past two weeks, summed into total PA volume of past two weeks, and divided by 14 days for daily averages. |
| **Intensity** | 2.5 – 6 | MET scores. Literature-derived classification per activity, with adjustments for older adults.<br>2.5 MET correspond to least intensive activity of light household activities or billiards. 6 MET correspond to the most intensive activity of playing tennis. | Mean intensity score of all reported activities of the past two weeks. |
| **Muscle strength** | 1-4 | The required lower body muscle strength to perform an activity. Scoring was validated against measured muscle strength of the quadriceps. Additionally, scores were also validated against hand-grip strength, which is seen as an indicator of overall strength. | Mean muscle strength score of all reported activities of the past two weeks. |
|  |  | A score of 1 included standing and sitting activities. Score 2 included activities related to walking. Score 3 represented activities which include running or cycling. Score 4 contained activities involving jumping and squatting actions. |  |
| **Mechanical strain** | 1-4 | Ground Reaction Forces (GRF), expressing multitudes of body weight. Classification of activities is supported by literature.<br>A score of 1 was given to activities with GRF values up to one time the body weight (non weight bearing activities and standing activities). Score 2 represented those activities with GRF values between one and two times body weight (weight bearing activities). Score 3 described those activities with GRF values between two and four times body weight (activities including explosive actions such as sprinting and running) and score 4 indicated physical activities with GRF greater than four times body weight (activities including jumping). | Mean mechanical strain score of all reported activities of the past two weeks. |
| **Turning actions** | 1-3 | Degree of which transversal rotations in the lower extremities were present for each activity<br>The turning action scores ranged from 1 to 3, with the lowest score indicating few to no transversal rotations during the activities, such as walking and bicycling, and the highest score representing activities associated with numerous turning actions, such as dancing and tennis. | Mean turning actions score of all reported activities of the past two weeks. |

**Abbreviations:** PA: Physical Activity; LAPAQ: LASA Physical Activity Questionnaire; LASA: Longitudinal Aging Study of Amsterdam; MET: Metabolic Equivalent of Task; GRF: Ground Reaction Forces
\* **Note:** Presented ranges represent observed values in the study sample and may not correspond to the full theoretical range of possible values. The range of duration was capped at 960 minutes (16 hours), as higher values of average daily duration of all forms of PA were deemed unrealistic.
More details and information of the development of the scoring instrument for the components of intensity, muscle strength, mechanical strain, and turning actions can be found in the original article of Verweij et al. (2010)

The averaged PA component scores at each available measurement wave were subsequently categorized into tertiles, in line with the validation study of these PA components [21]. A moderate correlation was observed between the PA component scores of muscle strength and intensity (r = 0.73), whereas correlations between all other PA component combinations were weak (ranging from r = 0.01 to r = 0.33) (see Appendix C).

#### 2.3.2. Demographics

Data on age (in years) and sex (men/women) were derived from population registries. Partner status was self-reported as *no partner*, *living together*, or *partner outside household*. Education level was assessed as highest attained education, and subsequently categorized into three levels of *Lower education* (no education/elementary/lower vocational), *Intermediate education* (general intermediate/intermediate vocational/general secondary), and *Higher education* (higher vocational/college/university).

#### 2.3.3. Other variables

Given the multitude of associations between PA and health outcomes, additional variables on general health and physical- and cognitive functioning were considered for descriptive purposes. Chronic conditions were self-reported by respondents and subsequently summed into a count variable, representing the number of conditions, and ranges in our data from 0 to 10. Perceived limitations due to health conditions was self-reported as *“Yes, severely”, “Yes, slightly”*, and *“No”*. Self-perceived health was assessed on a five-point Likert scale ranging from *“Poor”* to *“Excellent”*. Physical functioning was assessed using self-reported functional limitations. Respondents were asked how well they could perform the activities of 1) walking up and down a staircase of 15 steps without resting, 2) using own or public transportation, and 3) cutting their own toenails, and were all reported on a four-point Likert Scale (0 *“Yes, without help”* – 3 *“No I cannot”*). These three items reflect activities of daily living, and were summed into one functional limitation scale (range 0-9) to asses physical functioning [29], with higher scores indicating more limitations and thus poor functioning. Cognitive functioning was assessed using the Mini-Mental State Examination (MMSE) [30]. The MMSE is widely used in epidemiological studies and community surveys, both as a screener for cognitive impairment and as a measure for global cognitive functioning in older persons. The MMSE consists of 23 items and scores range from 0-30, with higher scores indicating better cognitive functioning.

### 2.4. Analysis

The analyses of this study consisted of a data preparation phase, which included multiple imputations to address missing data, followed by a stratification on self-reported sex, and the subsequent analysis phases: 1) identifying distinct trajectories for each PA component, stratified for men and women, and 2) assessing differences between these trajectories within each PA component.

Individuals trajectories were constructed with Sequence Analysis (SA) and subsequently grouped with Optical Matching (OM) into longitudinal clusters of similar trajectory patterns for each of the five PA components over the course of 10 years (4 measures; T0 t/m T3). In order to asses the robustness of the trajectories, sensitivity analyses were performed by repeating the main analysis over the course of 20 years (7 measures; T0 t/m T6), 30 years (10 measures; T0 t/m T9), as well as in the subsets of survivors and complete cases for each of the three follow-up periods (10, 20, and 30 years).

#### 2.4.1. Multiple imputations

Missing responses on the LAPAQ were evaluated, with study drop-out, refusal at the current wave, or being bedridden or in a wheelchair, considered as missing not at random (MNAR). Missing responses on the LAPAQ items of duration and frequency of PA activities, where other items of the LAPAQ or any other questionnaire was present, were considered missing at random (MAR). Not performing any activity at all according to the LAPAQ, while not already assigned MNAR was deemed unrealistic, and these records were recoded as MAR as well. Similarly, mean duration of more than 16 hours a day was also deemed unrealistic and coded as MAR. Records that contained only MNAR and/or MAR responses on all measurements were dropped, whereas records with at least one real-world observation on the LAPAQ were retained. The number of MAR and MNAR differed by wave, and are reported in Appendix A.

The MAR responses were imputed in IBM SPSS version 29 using Predictive Mean Matching (PMM), using 5 donors and 10 iterations as settings for the PMM. Imputation took place on the lowest level; the raw variables of duration and frequency for each activity type. All imputed variables on frequency and duration were used both as missing data indicator and outcome in the PMM. Additional indicators for imputation were cohort, sex, age, education level, waist circumference, urbanization level, household size, self-reported limitations in activities of daily living, physical performance scores, comorbidities, self-rated health, depressive symptoms, self-efficacy, social network, and social participation.

#### 2.4.2. Sequence Analysis and Optimal Matching

Sequence Analysis (SA) is a method to analyze different states over time, where each pattern forms a sequence. Each state is assigned a unique code, and transitions between different states can be evaluated. States in our analyses correspond to the tertile scores of each PA component, and were thus coded *High, Medium,* and *Low*. To account for the natural life-course, *deceased* was added as an additional state. These four states are mutually exclusive, and were used across all five PA components. Respondents could transition to any of the four states during each measurement wave, unless respondents were already in the *Deceased* state.

Optimal Matching (OM) is a common algorithm in SA, which calculates the distance between all observed sequences. It starts with the full dataset, and subsequently splits the data into clusters of similar sequences based on the calculated distances. This process is then repeated for the distances between these clusters, until all observations are captured into their own cluster. Based on these distances between the clusters and their corresponding split, the optimal number of clusters can then be selected by hand. Both SA and OM were performed with the TramineR package (version 2.2.10) in R (version 4.4.1).

#### 2.4.3. Assessing differences between trajectories

All trajectories identified with OM were compared across the demographic and other variables. These comparisons were done within each of the five PA components and separately for older men and women. Differences in the characteristics of the trajectory clusters were tested with the appropriate tests: an ANOVA for normally distributed continuous variables, Kruskall-Wallis for non-normally distributed continuous variables, and Chi-square test or a Fischer’s exact test for categorical variables. Due to the size of the tables, only the tables for the PA component of duration within men and women are presented in the main article, with all other PA components in the appendices (see Appendix F).

## 3. Results

### 3.1. Sample descriptives

The study included 4963 participants with an equal sex distribution (52% women), and a mean follow-up duration of 11 years (SD = 8.1), as described in Table 2. At baseline, respondents had a mean age of 66 years (sd = 8.6) and most respondents lived with a partner (70%), though women were more often without a partner (37%) compared to men (16%). Regarding socioeconomic characteristics, 51% of the sample had a lower education level, with a higher proportion of women (56%) than men (46%) in this category.

**Table 2:** Sample characteristics at baseline.

|  | **Overall**<br>**(N=4963)** | **Men**<br>**(N=2395)** | **Women**<br>**(N=2568)** |
| --- | --- | --- | --- |
| **Recruitment cohort** |  |  |  |
| Cohort 1 (1992) | 2954 (60 %) | 1432 (60 %) | 1522 (59 %) |
| Cohort 2 (2002) | 991 (20 %) | 470 (20 %) | 521 (20 %) |
| Cohort 3 (2012) | 1018 (21 %) | 493 (21 %) | 525 (20 %) |
| **Years participating in the study** |  |  |  |
| Mean (SD) | 11 (8.1) | 9.9 (7.8) | 12 (8.2) |
| **Age at baseline** |  |  |  |
| Mean (SD) | 66 (8.6) | 66 (8.7) | 66 (8.6) |
| **Partner status** |  |  |  |
| No partner | 1315 (26 %) | 375 (16 %) | 940 (37 %) |
| Partner (co-residence) | 3483 (70 %) | 1924 (80 %) | 1559 (61 %) |
| Partner (outside household) | 164 (3 %) | 95 (4 %) | 69 (3 %) |
| Missing | 1 (0.0%) | 1 (0.0%) | 0 (0%) |
| **Education level** |  |  |  |
| Lower Education | 2551 (51 %) | 1110 (46 %) | 1441 (56 %) |
| Intermediate Education | 1528 (31 %) | 732 (31 %) | 796 (31 %) |
| Higher Education | 881 (18 %) | 553 (23 %) | 328 (13 %) |
| Missing | 3 (0.1%) | 0 (0%) | 3 (0.1%) |
| **Functional limitation** |  |  |  |
| Median [IQR] | 0 [0 - 1.0] | 0 [0 - 1.0] | 0 [0 - 2.0] |
| Missing | 4 (0.1%) | 3 (0.1%) | 1 (0.0%) |
| **MMSE-score** |  |  |  |
| Mean (SD) | 27 (2.6) | 27 (2.5) | 27 (2.6) |
| Missing | 13 (0.3%) | 7 (0.3%) | 6 (0.2%) |
| **Number of chronic comorbidities** |  |  |  |
| Median [IQR] | 1.0 [1.0 - 2.0] | 1.0 [0 - 2.0] | 2.0 [1.0 - 3.0] |
| **Health problems limit normal activities** |  |  |  |
| Yes, severely | 530 (11 %) | 255 (11 %) | 275 (11 %) |
| Yes, slightly | 1016 (20 %) | 435 (18 %) | 581 (23 %) |
| No | 3406 (69 %) | 1699 (71 %) | 1707 (66 %) |
| Missing | 11 (0.2%) | 6 (0.3%) | 5 (0.2%) |
| **Self-perceived health** |  |  |  |
| Excellent | 712 (14 %) | 365 (15 %) | 347 (14 %) |
| Good | 2512 (51 %) | 1265 (53 %) | 1247 (49 %) |
| Fair | 1103 (22 %) | 502 (21 %) | 601 (23 %) |
| Sometimes good/bad | 505 (10 %) | 196 (8 %) | 309 (12 %) |
| Poor | 117 (2 %) | 61 (3 %) | 56 (2 %) |
| Missing | 14 (0.3%) | 6 (0.3%) | 8 (0.3%) |
| **Number of reported sports** |  |  |  |
| 0 | 2043 (41 %) | 1028 (43 %) | 1015 (40 %) |
| 1 | 1511 (30 %) | 700 (29 %) | 811 (32 %) |
| 2 or more | 1409 (28 %) | 667 (28 %) | 742 (29 %) |
| **Total PA duration: min/day** |  |  |  |
| Mean (SD) | 170 (110) | 130 (100) | 200 (110) |
| Missing | 33 (0.7%) | 14 (0.6%) | 19 (0.7%) |
| **PA component: Duration** |  |  |  |
| Long duration | 1677 (34 %) | 512 (21 %) | 1165 (45 %) |
| Medium duration | 1792 (36 %) | 830 (35 %) | 962 (37 %) |
| Short duration | 1461 (29 %) | 1039 (43 %) | 422 (16 %) |
| Missing | 33 (0.7%) | 14 (0.6%) | 19 (0.7%) |
| **PA component: Intensity** |  |  |  |
| High Intensity | 2138 (43 %) | 1036 (43 %) | 1102 (43 %) |
| Medium Intensity | 2073 (42 %) | 1020 (43 %) | 1053 (41 %) |
| Low Intensity | 719 (14 %) | 325 (14 %) | 394 (15 %) |
| Missing | 33 (0.7%) | 14 (0.6%) | 19 (0.7%) |
| **PA component: Muscle strength** |  |  |  |
| High Muscle strength | 2148 (43 %) | 1123 (47 %) | 1025 (40 %) |
| Medium Muscle strength | 1591 (32 %) | 676 (28 %) | 915 (36 %) |
| Low Muscle strength | 1191 (24 %) | 582 (24 %) | 609 (24 %) |
| Missing | 33 (0.7%) | 14 (0.6%) | 19 (0.7%) |
| **PA component: Mechanical strain** |  |  |  |
| High Mechanical Strain | 889 (18 %) | 474 (20 %) | 415 (16 %) |
| Medium Mechanical Strain | 3074 (62 %) | 1393 (58 %) | 1681 (65 %) |
| Low Mechanical Strain | 967 (19 %) | 514 (21 %) | 453 (18 %) |
| Missing | 33 (0.7%) | 14 (0.6%) | 19 (0.7%) |
| **PA component: Turning actions** |  |  |  |
| High Turning | 913 (18 %) | 355 (15 %) | 558 (22 %) |
| Medium Turning | 2930 (59 %) | 1309 (55 %) | 1621 (63 %) |
| Low Turning | 1087 (22 %) | 717 (30 %) | 370 (14 %) |
| Missing | 33 (0.7%) | 14 (0.6%) | 19 (0.7%) |

**Note:** Missing is only added to variables with any missing observation. The Functional Limitation score is used as a proxy for impaired physical functioning, whereas the Mini Mental State Examination is used as score for cognitive functioning.
**Abbreviations:** PA: Physical Activity, MMSE: Mini Mental State Examination

The study sample has a low number of chronic diseases (median = 1.0, IQR = 1–2) and a self-reported health status that was predominantly rated as excellent or good (65%) and only 2% of participants reporting poor health. Functional limitation scores were generally low (median = 0, IQR = [0.0–1.0]), and overall cognitive functioning was high (mean MMSE score 27, SD = 2.6).

### 3.2. Baseline PA components

PA component levels varied by sex. Women reported a higher mean PA duration (200 min/day, SD = 110; median = 180, IQR = [120; 250]) compared to men (130 min/day, SD = 100; median = 110, IQR = [64; 180]). Accordingly, 45% of women fell into the high-duration category, compared to just 21% of men, while men were more often in the short-duration category (43% vs. 16%). Differences were also evident in turning actions: 30% of men were in the low-turning category, compared to 14% of women. Sex differences were less pronounced for muscle strength, intensity, and mechanical strain.

### 3.3. PA components over time

After examining the transitions between the states for all PA components (Appendix D and OSF), it became clear that approximately half of the respondents remain in the same state, indicating stable PA behavior. Furthermore, for all five PA components, when respondents do transition to a different state, they were about equally likely to transition to a higher or lower state. Respondents who transitioned into a *deceased* state generally did so from a *low* PA component state. These transitions between states were comparable between men and women, although the underlying distributions of states did differ between PA components and sex.

### 3.4. PA component trajectories

The natural progression of PA components was analyzed over ten years. Visualization of the PA component scores over time revealed the large heterogeneity of individual sequences in PA behavior. The number of unique individual trajectories differed by PA component and sex (men: 144-151; women: 157-165, depending on the PA component, see OSF), indicating sex-differences in the stability of PA behaviors. Higher numbers of unique trajectories reflect a higher number of transitions between states.

Across all five PA components, men and women were classified into comparable trajectory clusters: *deceased early*, *deceased late*, *predominantly low*, *predominantly medium*, *predominantly high*, and *increasing* or *decreasing* over time. While overall trajectory structure was similar across sexes and PA components (Figure 1), participant distribution, baseline levels, and health correlates differed (Table 3).

**Figure 1.**
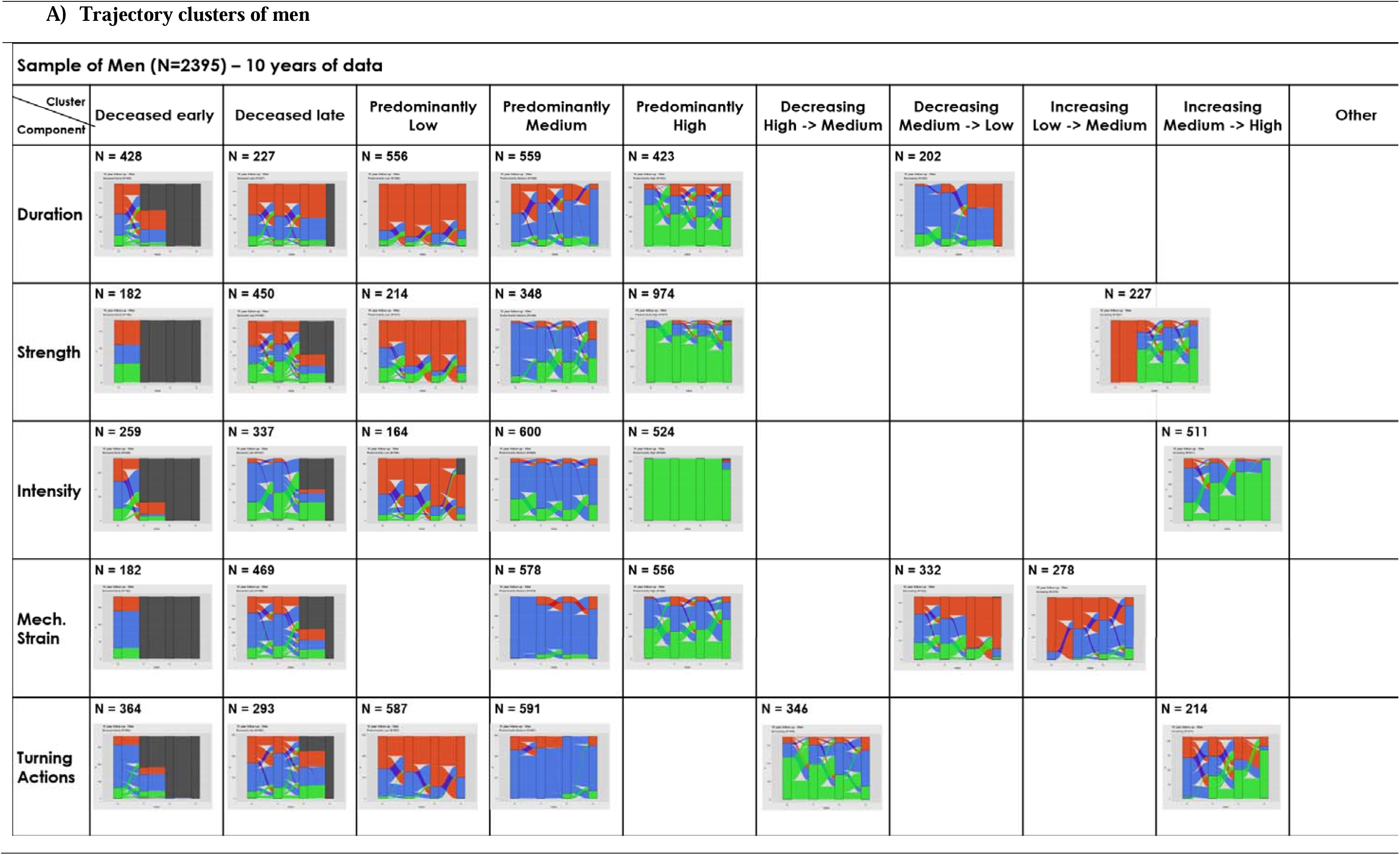
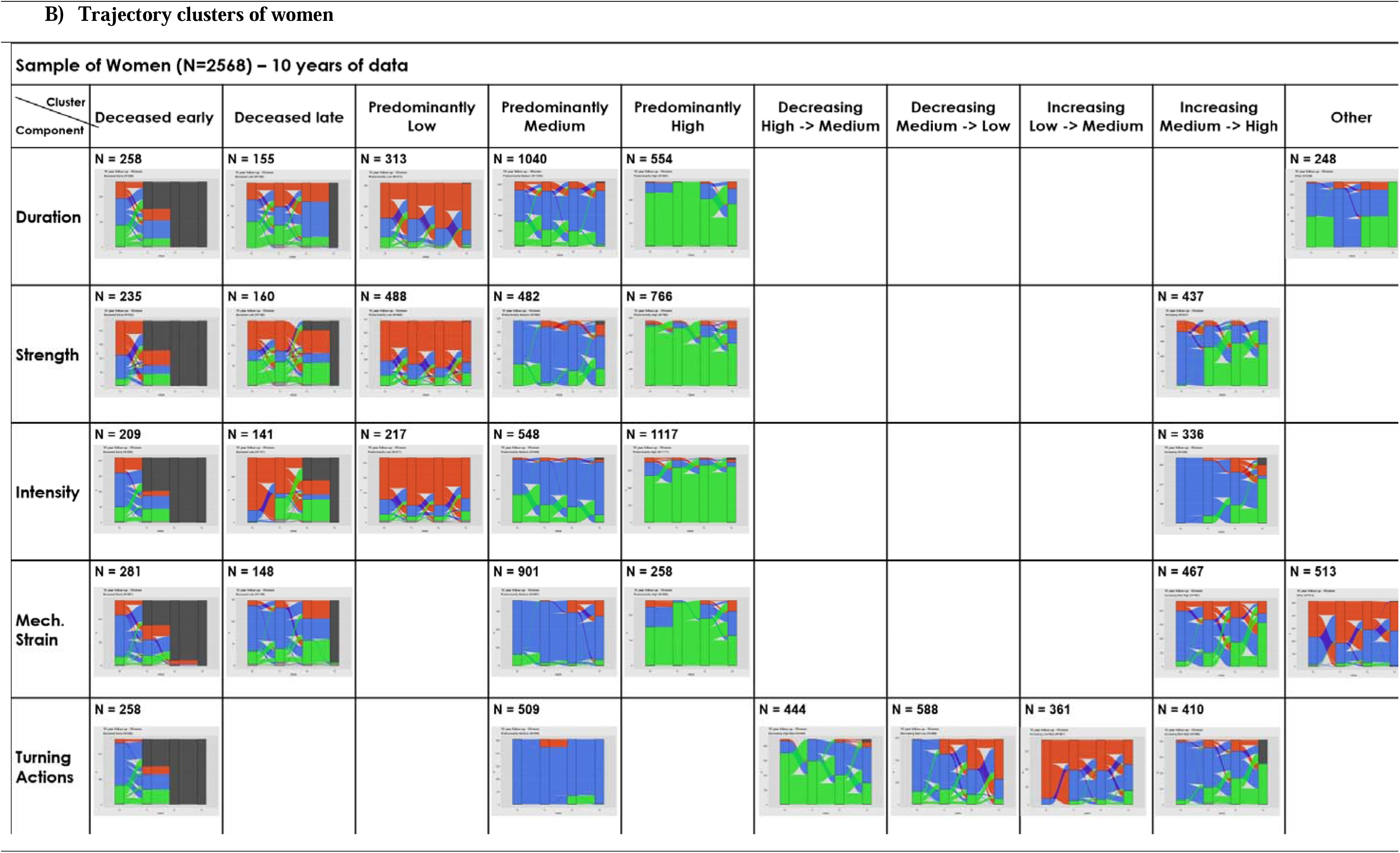
Sankey plots of the Physical Activity component trajectories in 4963 older men and women from the Longitudinal Aging Study Amsterdam **Note**: All Figures are stacked bar charts that represent the number of respondents within each state (red = low, blue = medium, green = high, black = deceased) and their transitions towards the states in the next time point. The transitions are calculated from one measurement tot the next, and do not necessarily reflect individual trajectories

**Table 3.**
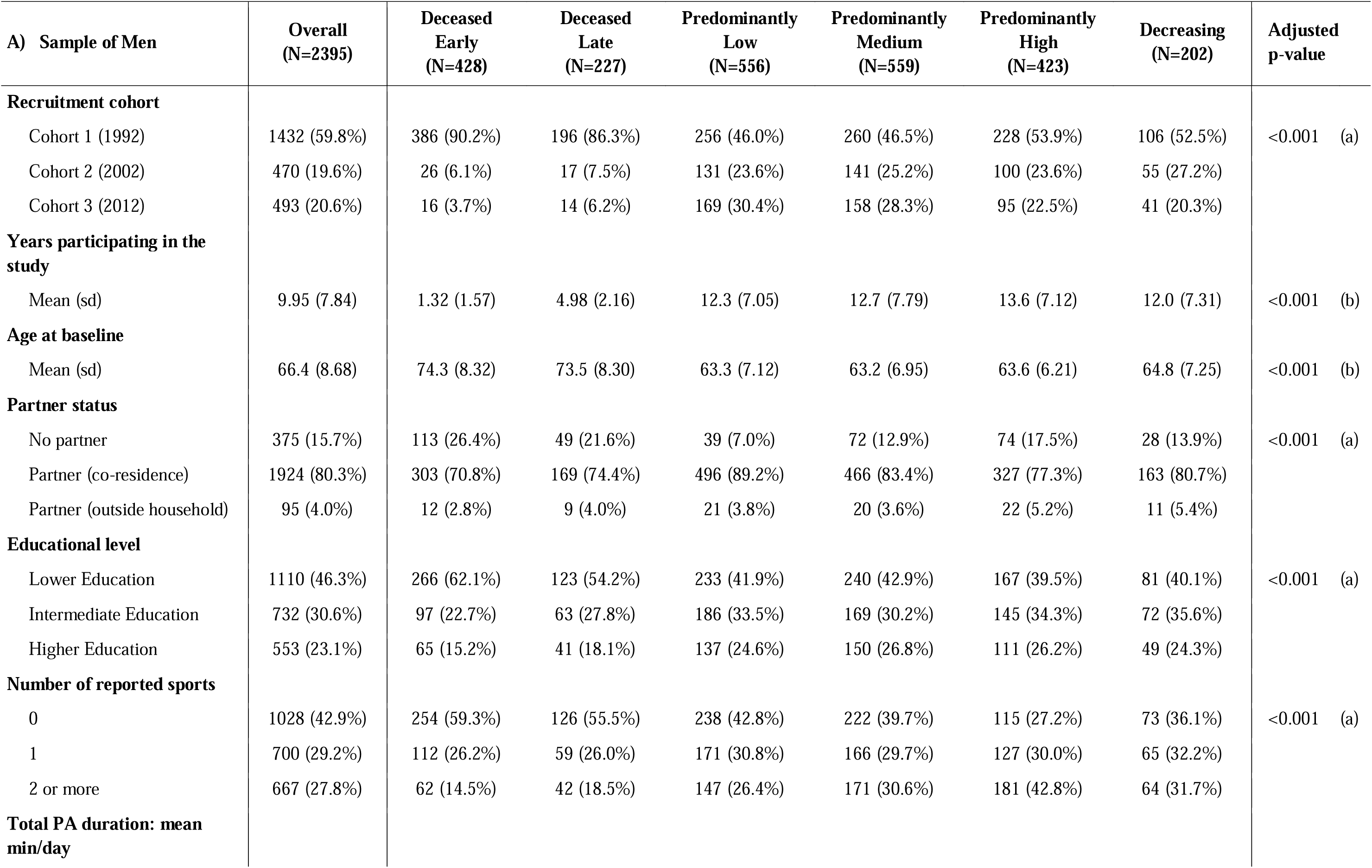
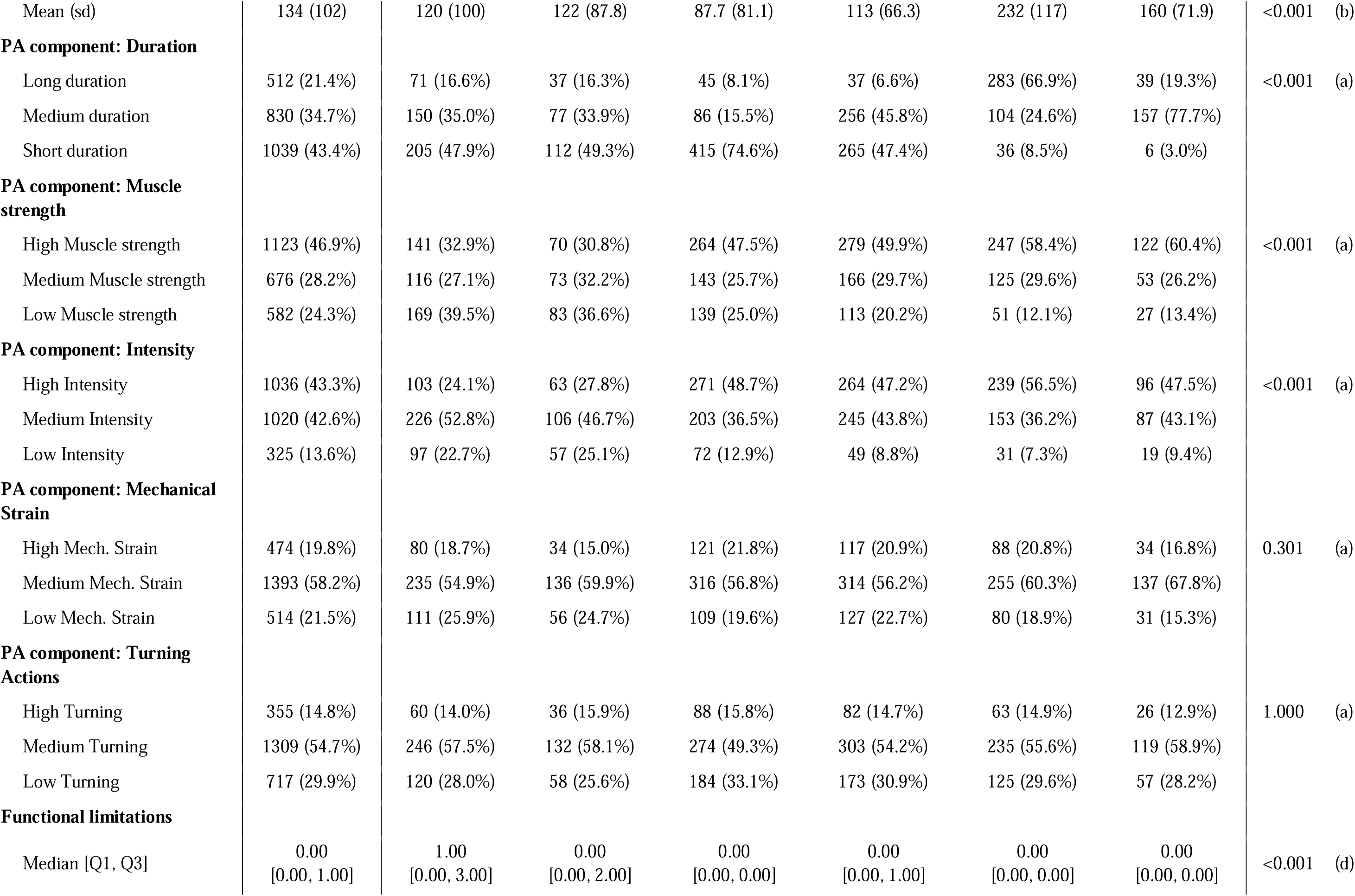
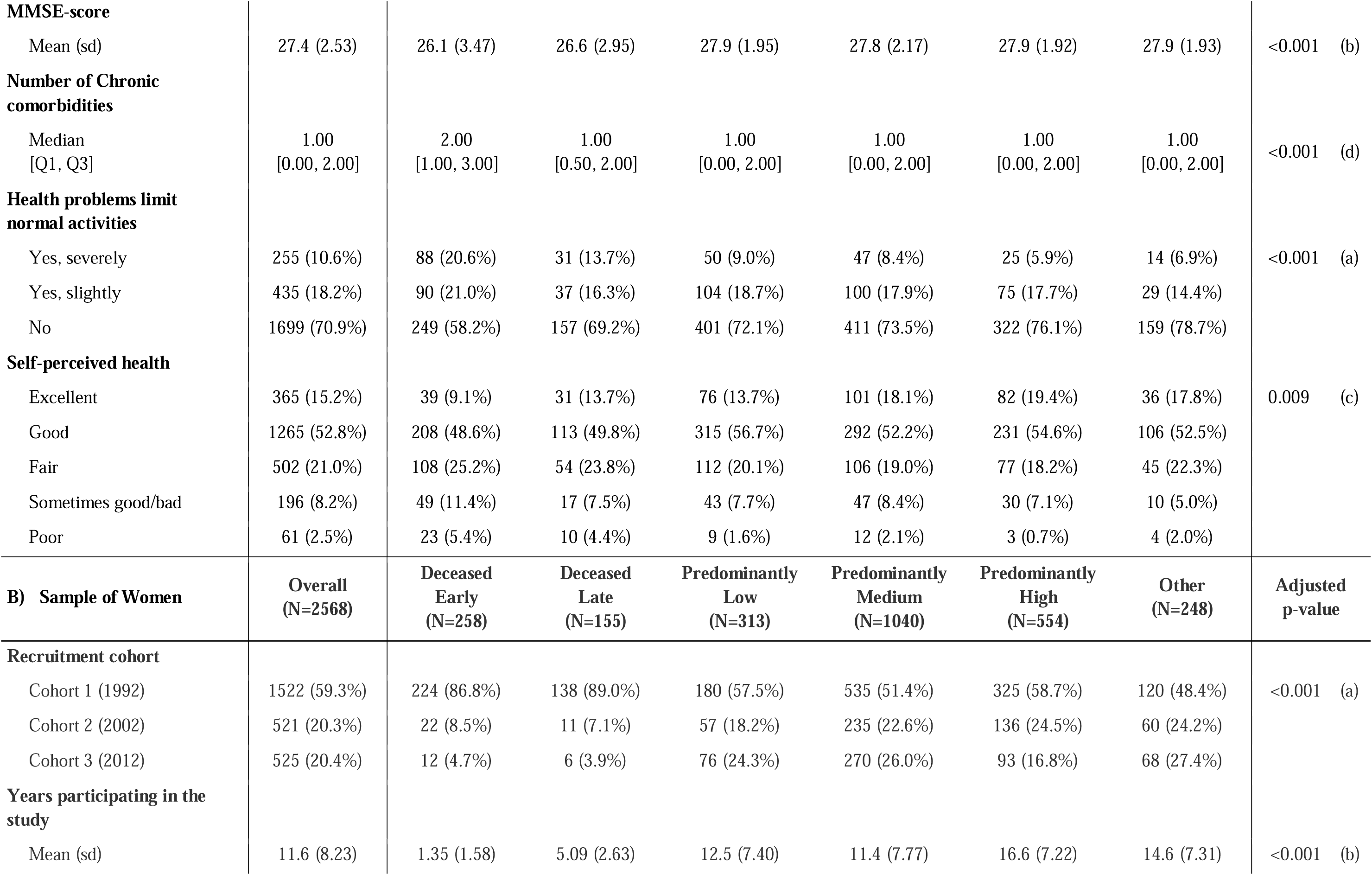
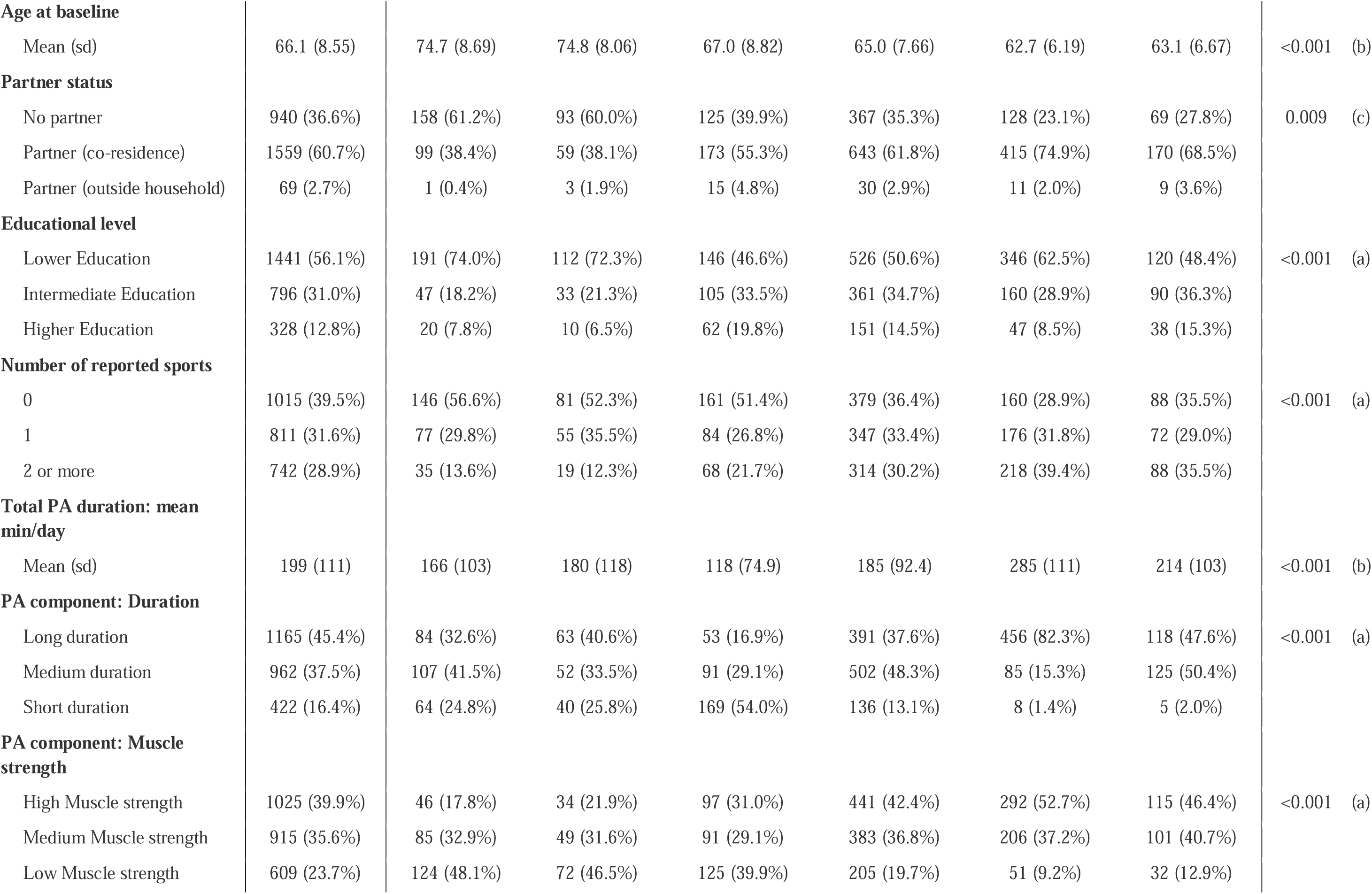
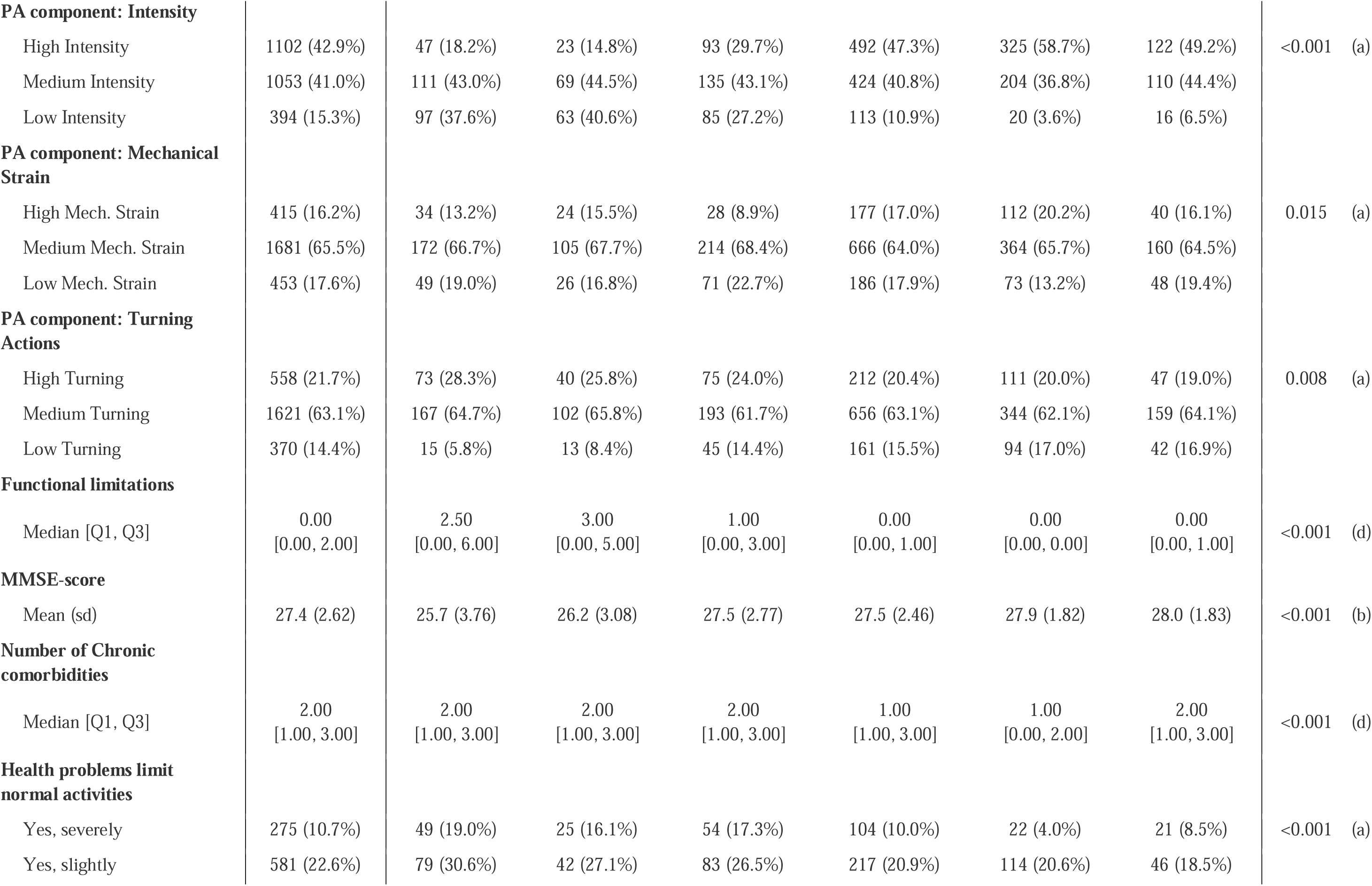
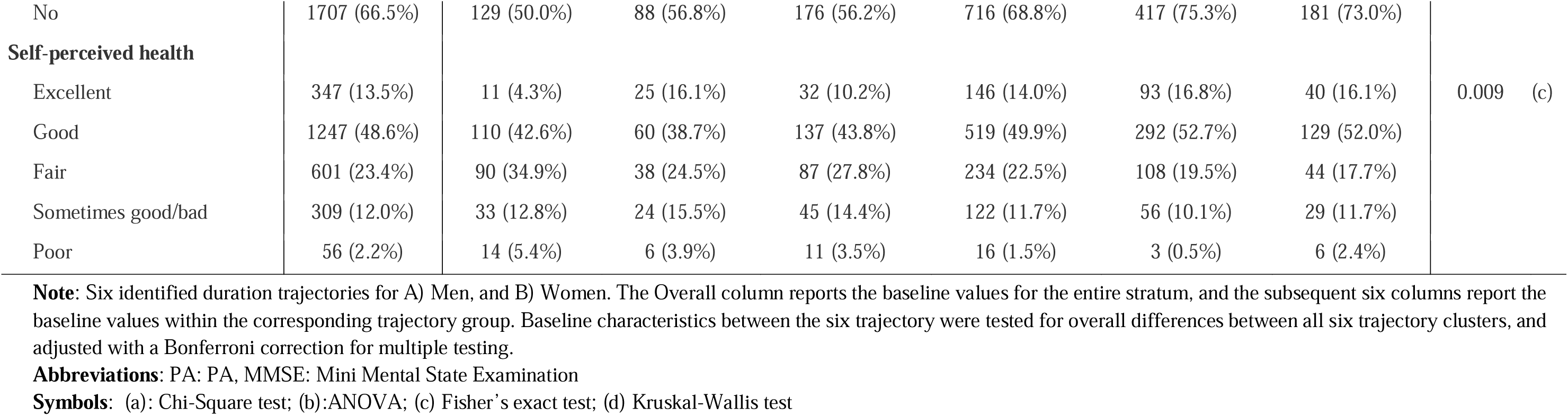
Baseline characteristics of the Duration trajectory clusters.

### 3.5. Shared patterns across PA components

Across all PA components and in both men and women, the *deceased early* and *deceased late* trajectories were consistently characterized by older baseline age, poorer cognitive functioning, more comorbidities, more functional limitations, and poorer self-perceived health than all other trajectories (Table 3). These mortality-related trajectories generally showed lower or intermediate PA component scores and minimal sports participation. This pattern was highly consistent across all PA components for both mortality-related trajectories.

Similarly to the mortality trajectories, *predominantly low* trajectories across PA components were also associated with less favorable health and more functional limitations at baseline, compared to *predominantly medium* or *predominantly high* trajectories. In contrast, members of the *predominantly high* trajectories were at baseline characterized by higher sports participation, better cognitive functioning, fewer functional limitations, and more favorable self-rated health. Although these gradients were observed in both sexes, women generally reported more comorbidities and functional limitations than men within comparable trajectories.

Despite these consistent cross-component patterns at the group level, trajectory memberships across PA components appeared largely independent. Initial correlations between trajectory memberships seemed substantial; however, these were largely attributable to the mortality trajectories. Participants classified into *deceased early* or *deceased late* trajectories naturally had fewer repeated measurements across all PA components, which systematically induced correlated trajectory assignment. After accounting for mortality status, correlations between trajectory memberships across PA components were attenuated, with correlation coefficients ranging from −0.01 to 0.24 in men and from −0.08 to 0.14 in women (Appendix C). This indicates that, beyond mortality, natural changes in PA components are independent of one another. For example, individuals in a *predominantly high* duration trajectory were not more likely to also follow a *predominantly high* intensity trajectory, and those in *predominantly low* muscle strength trajectories were not necessarily classified into *predominantly low* intensity or *predominantly low* mechanical strain trajectories. Thus, although trajectory structures and health gradients appeared broadly similar across PA components, trajectory memberships showed little overlap at the individual level.

### 3.6. Sex-specific differences

When comparing similar trajectory types between men and women, the *increasing* trajectories frequently differed by sex in their starting levels. For muscle strength and mechanical strain, *increasing* trajectories in women typically originated from medium baseline levels, whereas in men *increasing* trajectories starting from the low baseline levels was just as common. Another interesting comparison can be observed for turning actions, with more stable trajectories in men and more *increasing* and *decreasing* trajectories in women. Within turning actions, mortality driven and stable low trajectories were more common in men, but this differences was not observed for the other components.

Both had mostly stable trajectories, with main differences observed in the *other* trajectories. This trajectory group was harder to define due to its turbulent nature, with either many transitions back-and-forth (e.g. in the mechanical strain component for women), or with temporary declines in component scores (e.g. duration component in women). This category of trajectories was only observed in women, illustrating that changes in PA duration and mechanical strain scores over time were common enough to cluster together into their own trajectory. In other words, PA behavior might be more susceptible to changes over time for women as opposed to men.

### 3.7. Sensitivity Analyses

Sensitivity analyses using 20-year (N = 3945) and 30-year (N = 2954) follow-up data, complete case analyses (10 years: N = 2161, 20 years: N = 1608, 30 years: N = 1115), as well as analyses including only survivors of each follow-up period (10 years: N = 2068, 20 years: N = 1086, 30 years: N = 319), resulted in similar trajectory clusters and illustrate the robustness of our results (see Appendix E). At the longer follow-up periods, more trajectories were defined by becoming deceased, but both the 20-year and 30-year commonly reported trajectories of *predominantly low, predominantly medium, predominantly high*, and the occasional *decreasing* and *increasing* trajectory clusters.

Analyses on the subset of survivors within each follow-up period reported the same trajectory clusters as the main analyses, with logical exception of the *deceased early* and *deceased late* trajectories. Analysis of the trajectories over 20 and 30 years within the samples of survivors started to deviate more from the main results, likely due to the much smaller number of remaining respondents resulting in unstable clusters. Results of the complete case analyses were similar to the results of the main analysis.

## 4. Discussion

### 4.1. Main findings

The goal of this study was to identify and cluster similar long-term PA patterns over time for each PA component, examined separately for men and women. The identified trajectories share broadly similar structures across sexes and PA components, with members of *predominantly high* and *increasing* trajectories showing the most favorable health status at baseline. However, starting levels of the PA component scores, developmental patterns, and health correlates are strongly sex- and component-specific. Importantly, there was no systematic overlap between trajectory clusters across the different PA components. For example, individuals in the *predominantly high* duration trajectory were not more likely to also follow a *predominantly high* muscle strength or intensity trajectory, compared to any of the other duration trajectories. This finding highlights that different PA components behave differently and independently of each other over time at the individual level.

### 4.2. Comparison to literature

This study builds on previous research by Verweij et al.[16], who identified four distinct PA components; intensity, muscle strength, mechanical strain, and turning actions, and demonstrated their independent associations with health outcomes [21]. Our study extends these findings by adding duration as an additional PA component, and by showing how all five PA component scores change over time, revealing substantial individual variation and distinct trajectory patterns.

The large individual heterogeneity in PA behaviors led to an optimal data-driven solution of six main trajectories of PA behaviors, although the shapes of these trajectories differed by components and sex. Finding six trajectories of PA behavior in a cohort study of community dwelling older adults exceeds the number commonly found in the literature [31–34], where studies often report three to five trajectories of PA behaviors. However, some differences were expected due to differences in methods and analytical approach. For example, our study was the only one using Sequence Analysis with Optimal Matching. We also included deceased status as a natural state, which resulted in two of our identified trajectories being defined by the moment of death of the respondents. The others commonly handled deceased status as lost to follow-up or exclusion criteria. Furthermore, existing literature analyzed the changes in a summary score of duration, frequency, and/or intensity, while we investigated the PA components of duration, intensity, muscle strength, mechanical strain, and turning actions separately.

Despite these differences, the articles of Hassan et al. [34], SanchezℒSanchez et al. [31] and Wang et al.[33] are comparable due to the population of community-dwelling older adults, large sample size (1041 to 8227 respondents), fairly equal sex distributions (44% to 59% women), study duration (5 to 10 years), and number of measurements (2 to 4 measures). Direct comparison to Lounassalo et al. [32] could not be made, as this is paper is a review of multiple studies. Our identified trajectories align with the trajectories of *predominantly low* [31–34], *predominantly medium* [31–34], *predominantly high* [31–33], *decreasing* [31–34], and *increasing* [31–33] trends from the literature.

Women reported more *increasing* and *other* trajectories compared to men, while men reported more *decreasing*, *predominantly low,* and *deceased* trajectories. The latter one is not so surprising, as men tend to have a lower life-expectancy then women. However, the finding that men tend to become and stay less physically active than women is an important difference. Existing literature already showed that PA levels decline most notably in the leisure activities of older men, while for older women the declines in work-related activity are offset by an increase in household activities [35,36]. Although we did not identify a distinct *declining* trajectory for women in the duration component, several trajectories such as the *deceased late*, *predominantly low,* and *predominantly medium*, showed slight declines in duration over time. Combining this ‘hidden’ decline with the offset of work-related PA to household activities in women might explain the difference in trajectories. However, literature shows that women have lower intra-individual variability in PA behaviors [37], while we found that women had higher intra-individual variability (resulting in the *other* trajectories), but this difference could potentially be explained by the difference in measurement intervals (per minute vs. 3-yearly) and follow-up time (206 days vs. 10 years).

### 4.3. Strengths and limitations

Using data from LASA is a strength, as it provided extensive and high-quality data. The large number of respondents allowed for identification of uncommon trajectories, that could be overlooked in smaller samples such as having two different types of *decreasing* and *increasing* trajectories withing the same PA component. Furthermore, the large number of informative variables allowed us to impute missing LAPAQ values and allowed us to look more extensively into developments in PA behaviors. Another strength of the LASA dataset is the possibility to run extensive sensitivity analyses over different follow-up periods, hereby ensuring robust findings.

Another key strength of this study is the stratification by sex, which is often overlooked in PA research. While many studies typically adjust for sex in their analyses, stratification by sex allows for more sex-specific insights. A final strength of this study is the use of data-driven statistical clustering techniques to identify clusters of similar individual trajectories over time. This data-driven approach is not dependent on a-priori categorization of PA behaviors over time [38].

Despite being a strength, the analyzed data induces some limitations as well. First, the three-year intervals between LASA measurement waves may have resulted in missing important information about short-term changes in PA, particularly in later life when individuals’ health can decline more rapidly. Nevertheless, as the sensitivity analyses illustrated the robustness of the identified trajectory clusters, more measures would not necessarily have resulted in different trajectories.

Another limitation of the data lies in the use of self-reported PA data obtained by the LAPAQ. The LAPAQ reflects the two weeks prior to the interview, making it susceptible to recall bias. However, the LAPAQ is specifically designed for the LASA study, and validation of the LAPAQ against accelerometric data has shown that self-reported PA duration is equally reliable in older adults [28]. Furthermore, commonly used device-based PA measurements do not capture the specific type of activity performed, and therefore are not suitable to assess all five PA components simultaneously.

The use of multiple imputations posed a minor limitation, despite being one of the strengths. Since the multilevel imputation models failed to converge, we were forced to impute each observation independently. When comparing the imputed values to the complete cases, we observed an overestimation of the frequency of sport activities, specifically an overestimation of dancing and exercising on a home trainer. However, in next steps of our study, the PA component scores were first assigned to each of the six activity types (walking, cycling, light household, heavy household, and two most frequently performed sports), subsequently averaged over all these activities, and finally categorized into tertiles. Therefore, the effect of an overestimation of one of these six activity types will be minimal. This is supported by the observation that the distributions and transitions over time in each PA component of the imputed data closely match those of the complete cases (see Appendix E and OSF).

A point of discussion regarding the use of data-driven clustering techniques is the decision on the optimal number of clusters. In Sequence Analysis, this decision is based on the insertion and deletion costs of changing one sequence into another [39]. With these costs, distances between the sequences are calculated with Optimal Matching (OM). This process includes a subjective trade-off between statistical fit criteria and interpretability of the obtained clusters. To promote transparency on this trade-off procedure, and facilitate replication of our findings, we report all our code and findings in detail on the Open Science Framework.

### 4.4. Suggestions for future research

Future research could explore the determinants of the identified PA trajectories that influence the short-term and long-term change in the PA components over time in older men and women. These determinants could include major life events, such as illness, injury, or caregiving responsibilities. Cultural and social influences may also play a major role in shaping PA behaviors in older adults. Furthermore, it would be valuable to investigate the associations of the PA component trajectories with long-term health outcomes, in order to identify the optimal PA patterns which could subsequently support the development of more tailored and sustainable PA guidelines and interventions in later life.

## Conclusion

Later-life physical activity patterns changed in multiple, distinct ways over time, with differences between men and women and across PA components?. Concordance between components was limited; being active in one dimension (e.g. duration or intensity) did not necessarily coincide with change in other (e.g. muscle strength, mechanical strain or turning actions). The only clear overlap between activity patterns across components was related to participants who died earlier during follow-up. However, similar trajectories shapes did suggest generalizable associations across components with health status at baseline, with the *predominantly low* trajectories correlating with more health problems, and *increasing* and *predominantly high* trajectories being associated with better health status and more sport participation. These findings remained consistent even when we focused on longer follow-up, only completely observed records, or only included participants who survived the entire follow-up period. This shows that physical activity in older age is multidimensional and cannot be reduced to “how long” or “how intensively” people move. Recognizing that older adults follow different activity paths across these components can help health care professionals and policymakers design targeted and sustainable interventions that align with individual activity patterns and support healthy aging.

## Data Availability

All data analysed in the present study are owned by the Longitudinal Aging Study of Amsterdam (LASA), and are available upon upon reasonable request to this cohort study.

https://www.lasa-vu.nl/en/

## Availability of data and materials

All utilized data is owned by the Longitudinal Aging Study Amsterdam. As such, data can be requested through their website: www.lasa-vu.nl/en/. The design of this study, including all utilized datafiles, is registered at the Longitudinal Aging Study Amsterdam and can be requested through their website. R code and SPSS syntaxes for replicating analyses are available on request from the Longitudinal Aging Study Amsterdam and are publicly accessible at the Open Science Framework project page (https://doi.org/10.17605/OSF.IO/G42YB).

