## Appendices for "Trajectories of physical activity components among community-dwelling older adults"

Last updated at (dd-mm-yyyy): 28-01-2026

### Appendix A – Analytical set**.**

| Analytical sets of respondents with any physical activity data | | | | | | | | | | | | |
| --- | --- | --- | --- | --- | --- | --- | --- | --- | --- | --- | --- | --- |
|  | Block 1 (T0 t/m T3) | | | |  | | | | |  | | |
|  | Block 2 (T0 t/m T6) | | | | | | | | |  | | |
|  | Block 3 (T0 t/m T9) | | | | | | | | | | | |
|  | Time 0  Baseline | Time 1  Year 3 | Time 2  Year 6 | Time 3  Year 9 | | Time 4  Year 13 | Time 5  Year 16 | Time 6 Year 19 | Time 7 Year 23 | | Time 8 Year 26 | Time 9 Year 29 |
| Cohort 1 | *B  '92-'93* | *C  '95-'96* | *D  '98-'99* | *E  '01-'02* | | *F  '05-'06* | *G  '08-'09* | *H  '11-'12* | *I  '15-'16* | | *J  '18-'19* | *K  '21-'22* |
| Cohort 2 | *2B '02-'03* | *F  '05-'06* | *G  '08-'09* | *H  '11-'12* | | *I  '15-'16* | *J  '18-'19* | *K  '21-'22* |  | |  |  |
| Cohort 3 | *3B '12-'13* | *I  '15-'16* | *J  '18-'19* | *K  '21-'22* | |  |  |  |  | |  |  |
| Valid data | 4177 | 3310 | 2576 | 2272 | | 1440 | 1155 | 975 | 364 | | 220 | 133 |
| Invalid | 33 | 38 | 21 | 23 | | 17 | 10 | 15 | 7 | | 3 | 2 |
| Missing | 753 | 1326 | 1681 | 1587 | | 996 | 947 | 818 | 402 | | 314 | 231 |
| Deceased | 0 | 289 | 685 | 1081 | | 1492 | 1833 | 2137 | 2181 | | 2417 | 2588 |
| Total included | 4963 | | | | | 3945 | | | 2954 | | | |

A total of 130 respondents were filtered out prior to filling in this table, as they had no LAPAQ data on any measurement wave. An additional 39 respondents were dropped, due to being bedridden, bound to wheelchair, and/or deceased on all waves. Imputations of these missing records would be invalid, as it would simulate fictive records as opposed to imputing missing observations.

Valid data were completely observed LAPAQ records.
Invalid records were refusals, and respondents who were bedridden or wheelchair-bound and can be considered ‘missing not at random’. These observations were analyzed as missing.
Missing records were shortened interviews (where the LAPAQ was skipped), incomplete records (where the LAPAQ was not completed for unknown reasons), or terminated interviews and can be considered ‘missing at random’. These records were later on imputed and added to the data for analysis.
Respondents are recorded as deceased if they were deceased before the approach of the corresponding wave.
Deceased was an exclusion criteria at baseline measures (T0), as these records would not add any PA data. Reported numbers on deceased correspond to respondents that died during the follow-up period of a certain analytical block, with exception of baseline.

### Appendix B – PA component scoring instrument

| LAPAQ activity | Intensity component score | Strength component score | Mechanical strain component score | Turning actions component score |
| --- | --- | --- | --- | --- |
| General: |  |  |  |  |
| Walking outdoors | 3.5 | 2 | 2 | 1 |
| Bicycling | 4.5 | 3 | 1 | 1 |
| Light household | 2.5 | 2 | 1 | 2 |
| Heavy household | 4.5 | 4 | 2 | 2 |
| Sports: |  |  |  |  |
| 1. Distance walking | 4.0 | 3 | 2 | 1 |
| 2. Distance cycling | 6.0 | 4 | 1 | 1 |
| 3. Gym/game | 4.0 | 2 | 2 | 2 |
| 4. Home trainer | 4.0 | 2 | 1 | 1 |
| 5. Swimming | 5.0 | 3 | 1 | 2 |
| 6. (Folk) Dancing | 5.0 | 2 | 3 | 3 |
| 7. Bowling/jeu de boules | 3.5 | 3 | 2 | 2 |
| 8. Tennis/badminton | 6.0 | 3 | 3 | 3 |
| 9. Jogging/running/speed walking | 6.0 | 3 | 3 | 1 |
| 10. Rowing | 5.5 | 3 | 1 | 1 |
| 11. Sailing | 3.0 | 2 | 1 | 1 |
| 12. Billiards | 2.5 | 3 | 1 | 1 |
| 13. Fishing | 3.0 | 1 | 1 | 1 |
| 14. Soccer/basketball/ hockey | 6.0 | 4 | 4 | 3 |
| 15. Volleyball/baseball | 5.0 | 4 | 4 | 3 |
| 16. Skiing | 6.0 | 4 | 2 | 3 |
| 17. All other activities | 4.0 | 2 | 2 | 2 |
| Scoring instrument originally published in:  Verweij, L.M., Van Schoor, N.M., Dekker, J. and Visser, M.. 2010. “Distinguishing four components underlying physical activity: a new approach to using physical activity questionnaire data in old age.” BMC Geriatrics 10(1):20. doi:10.1186/1471-2318-10-20. Abbreviations: LAPA: LASA Physical Activity Questionnaire; LASA: Longitudinal Aging Study of Amsterdam | | | | |

### Appendix C – Correlation of PA components

Through these correlation matrices it becomes clear that correlations between the tertiles of components is mainly driven by deceased status.

**Figure C:**

Correlation matrices of the PA components scores and trajectories

1)
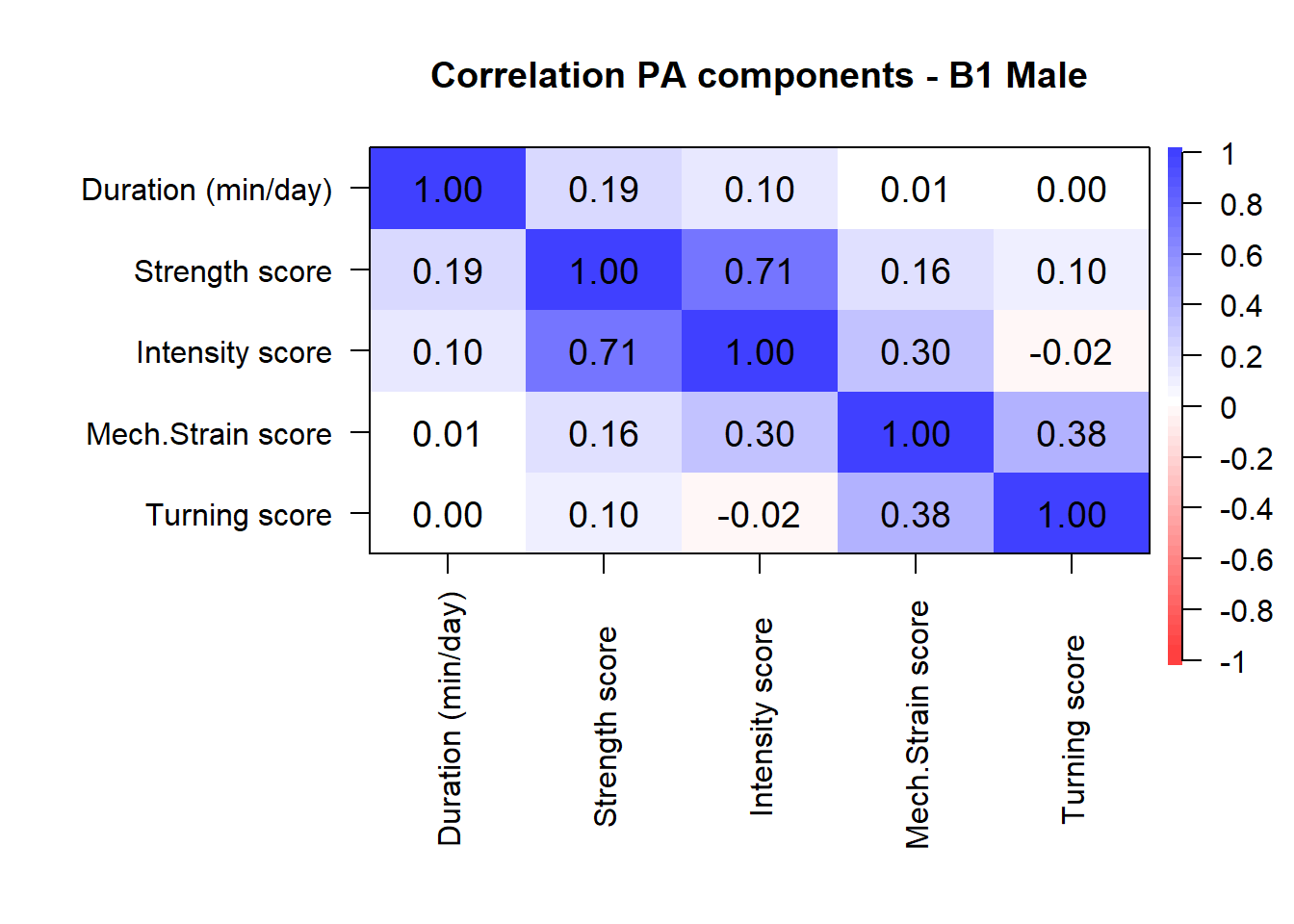
**Men – Correlation of PA component scores**

2)
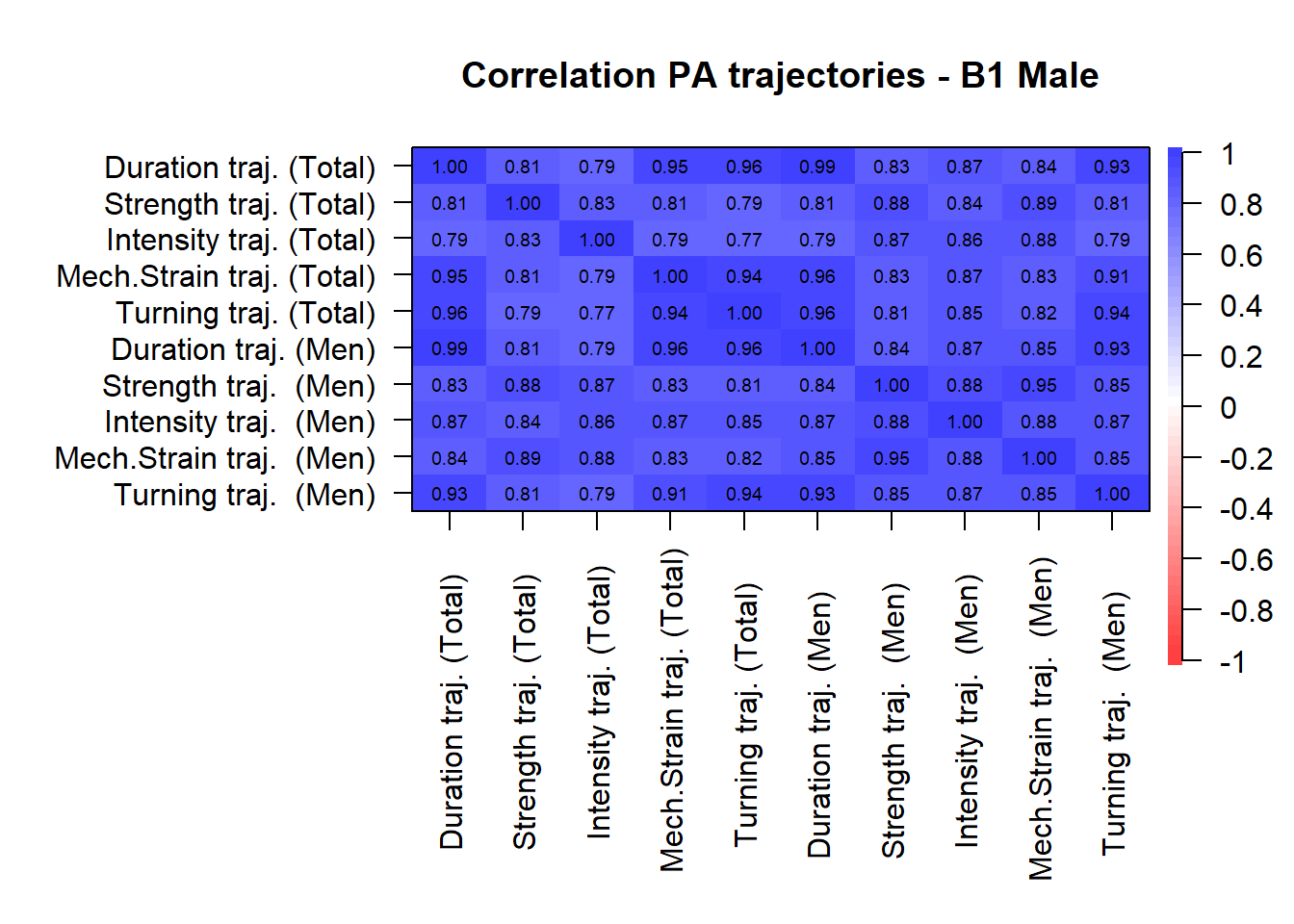
**Men – Correlation of PA component trajectories**

3)
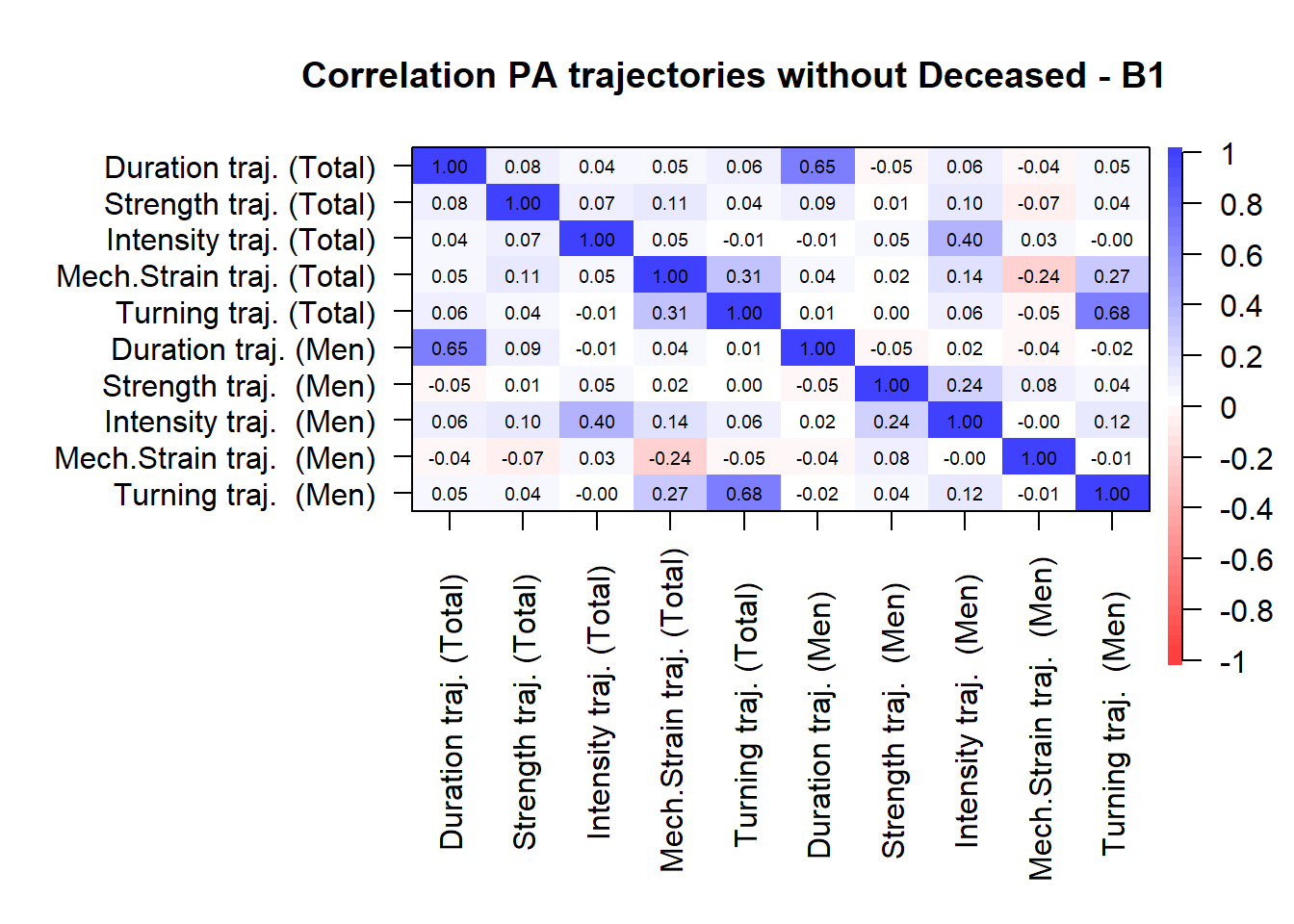
**Men – Correlation of PA component trajectories (with exclusion of deceased trajectories)**

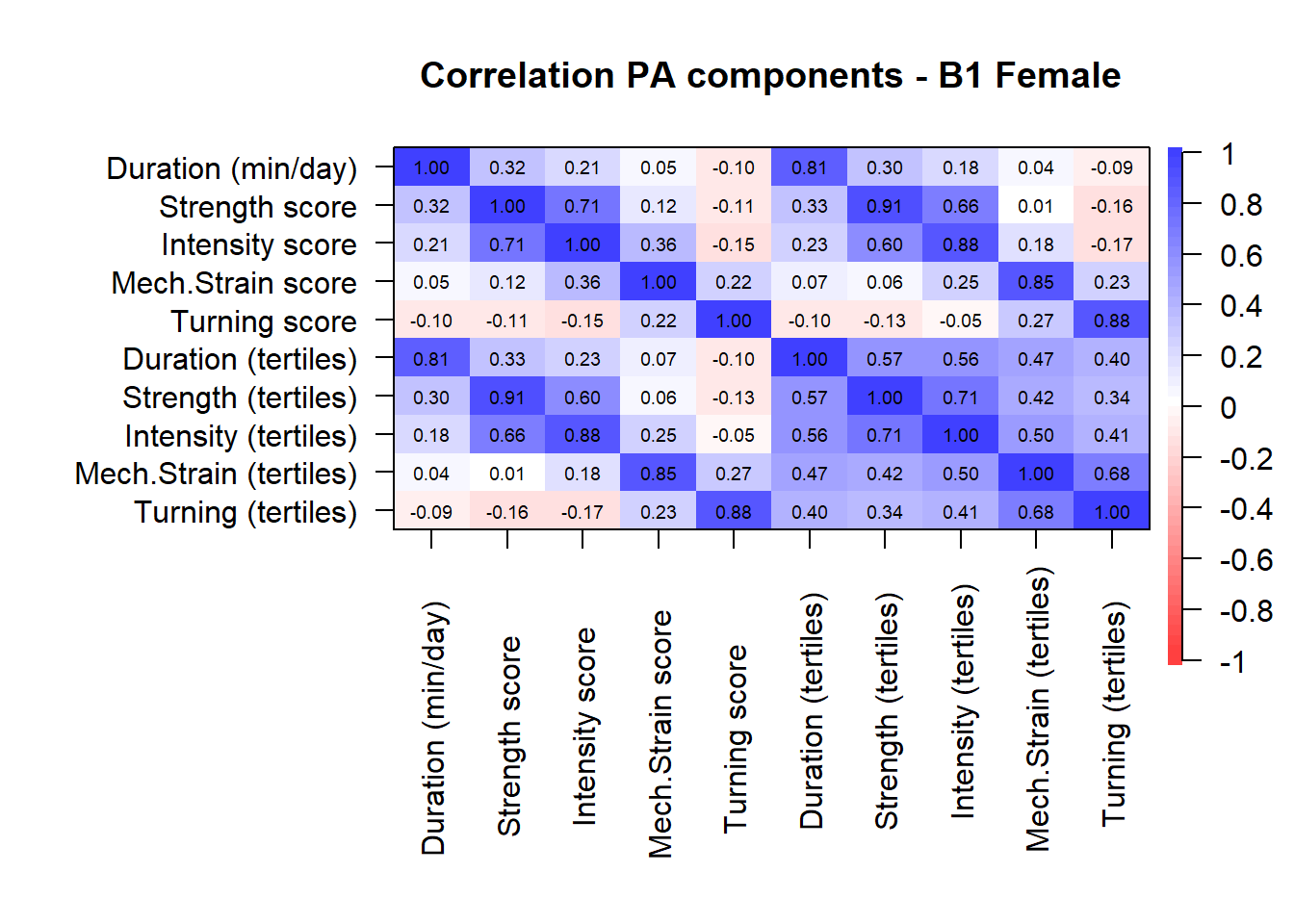

4)
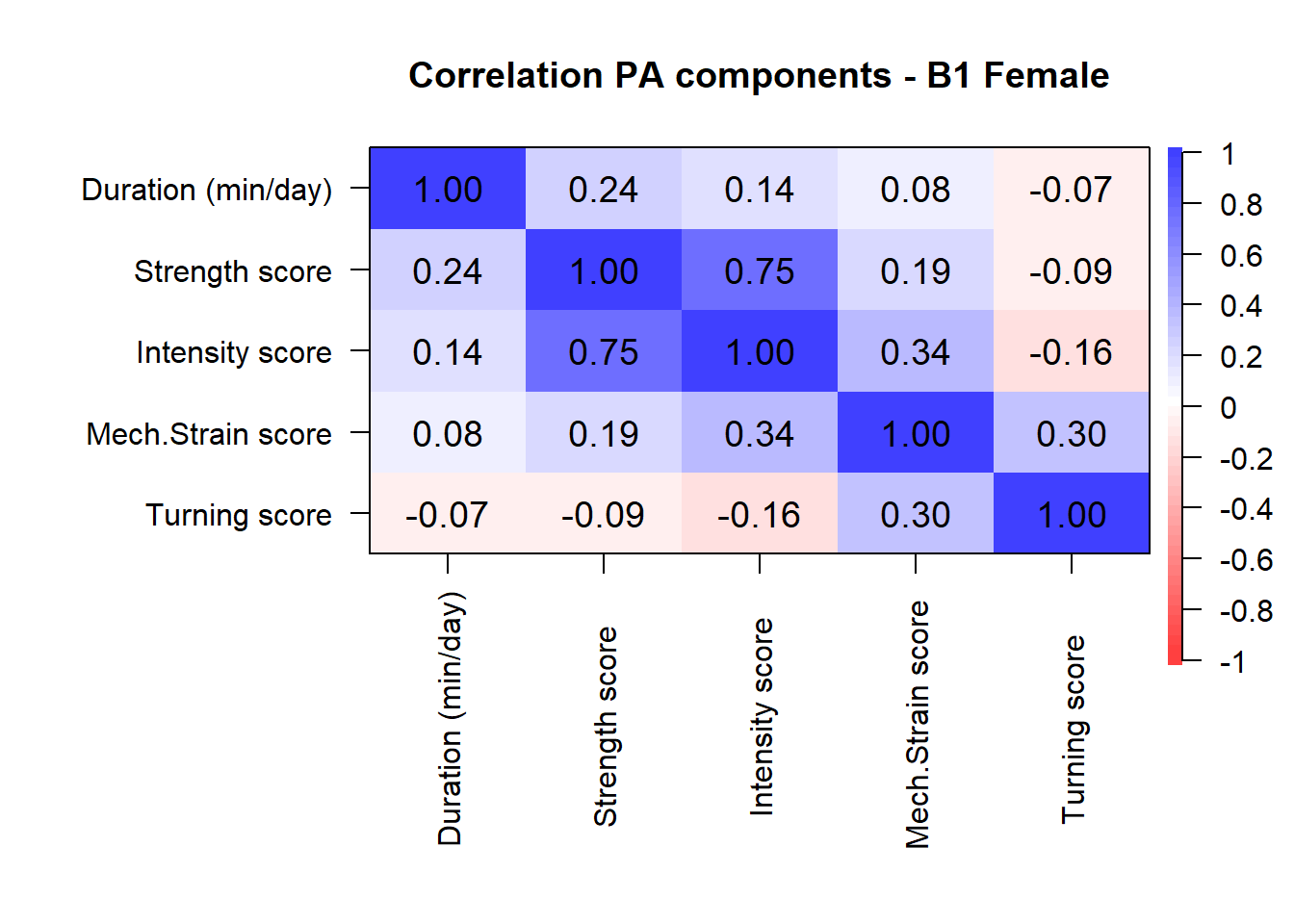
**Women – Correlation of PA component scores**

5)
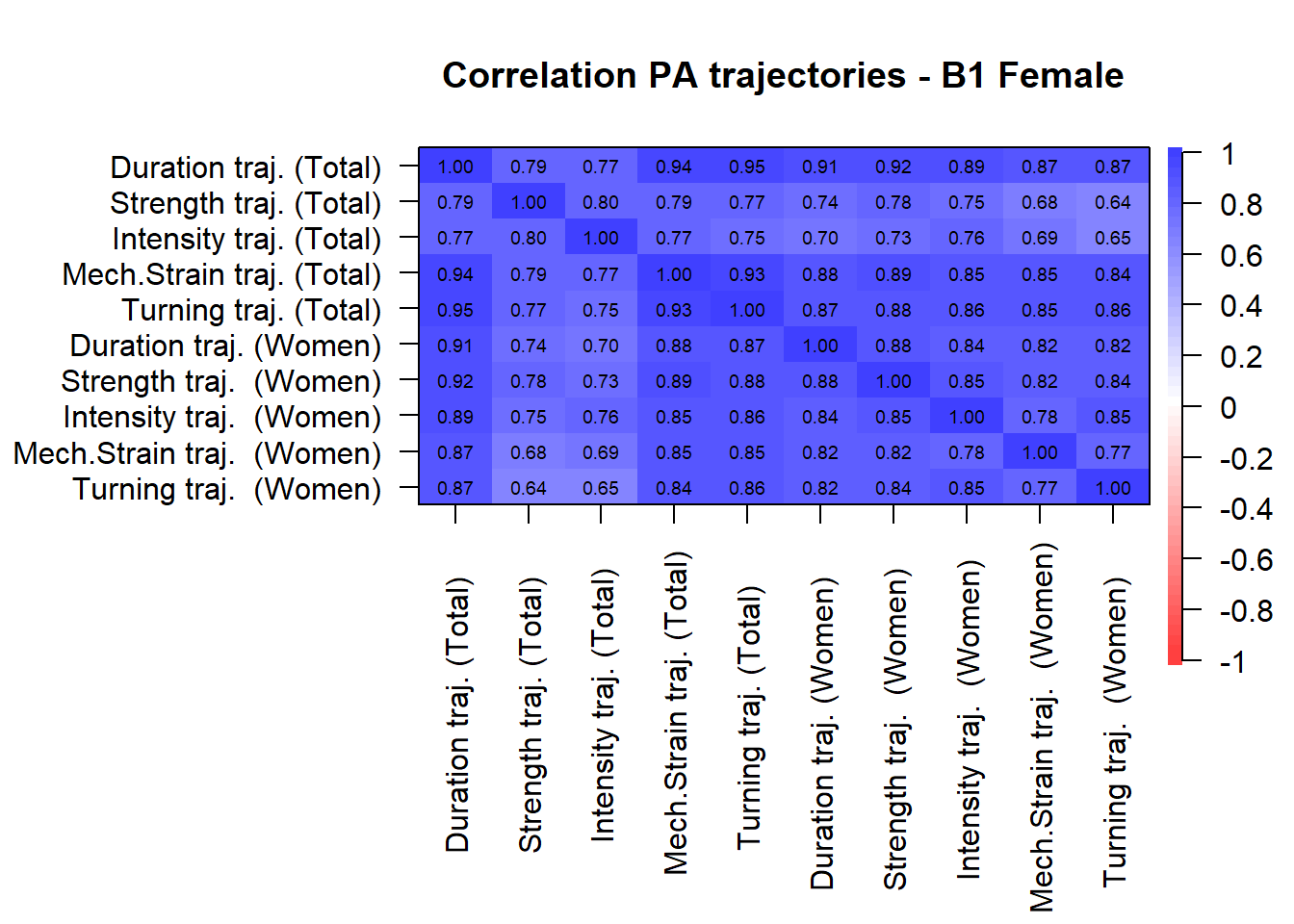
**Women – Correlation of PA component trajectories**

6)
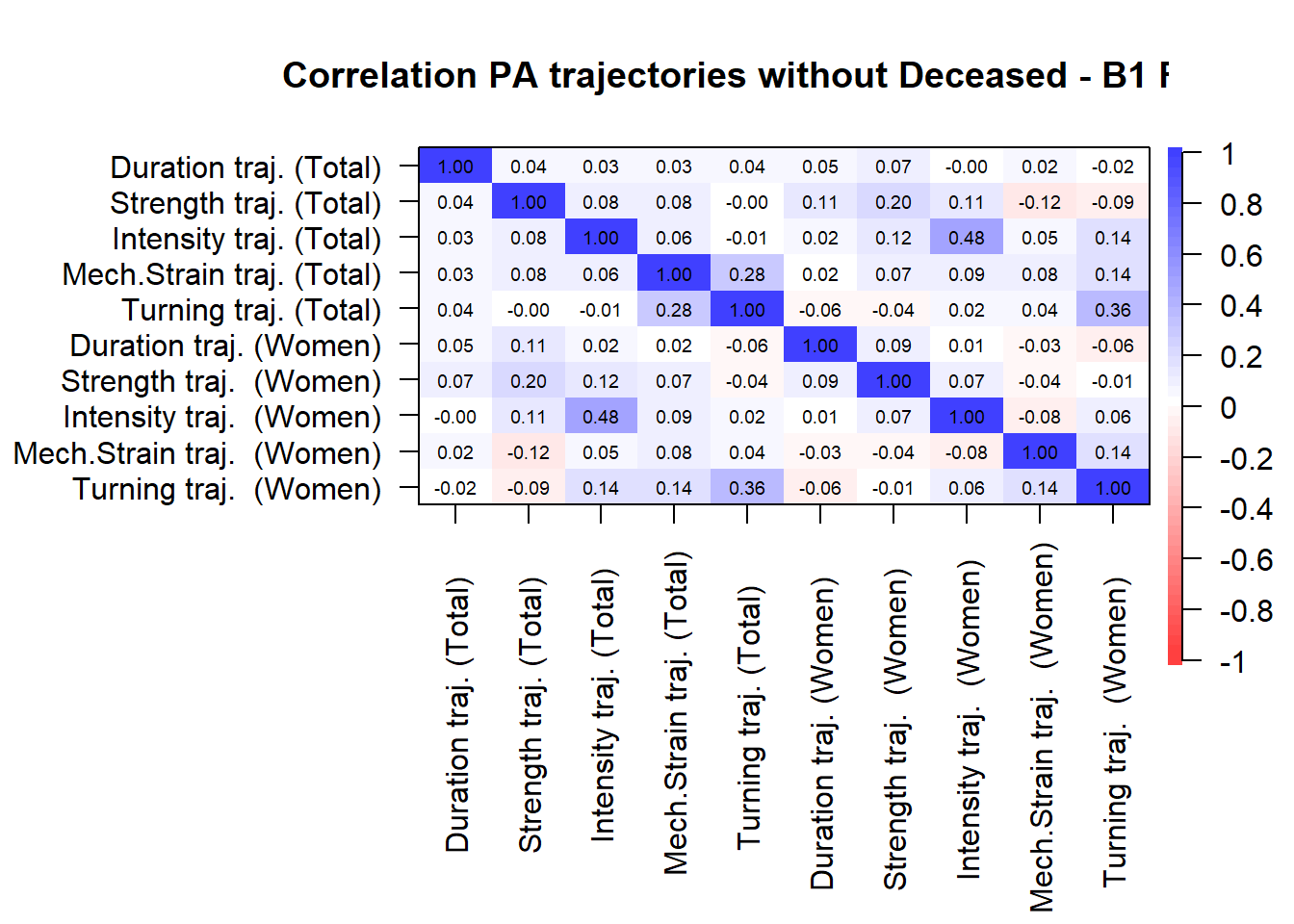
**Women – Correlation of PA component trajectories (with exclusion of deceased trajectories)**

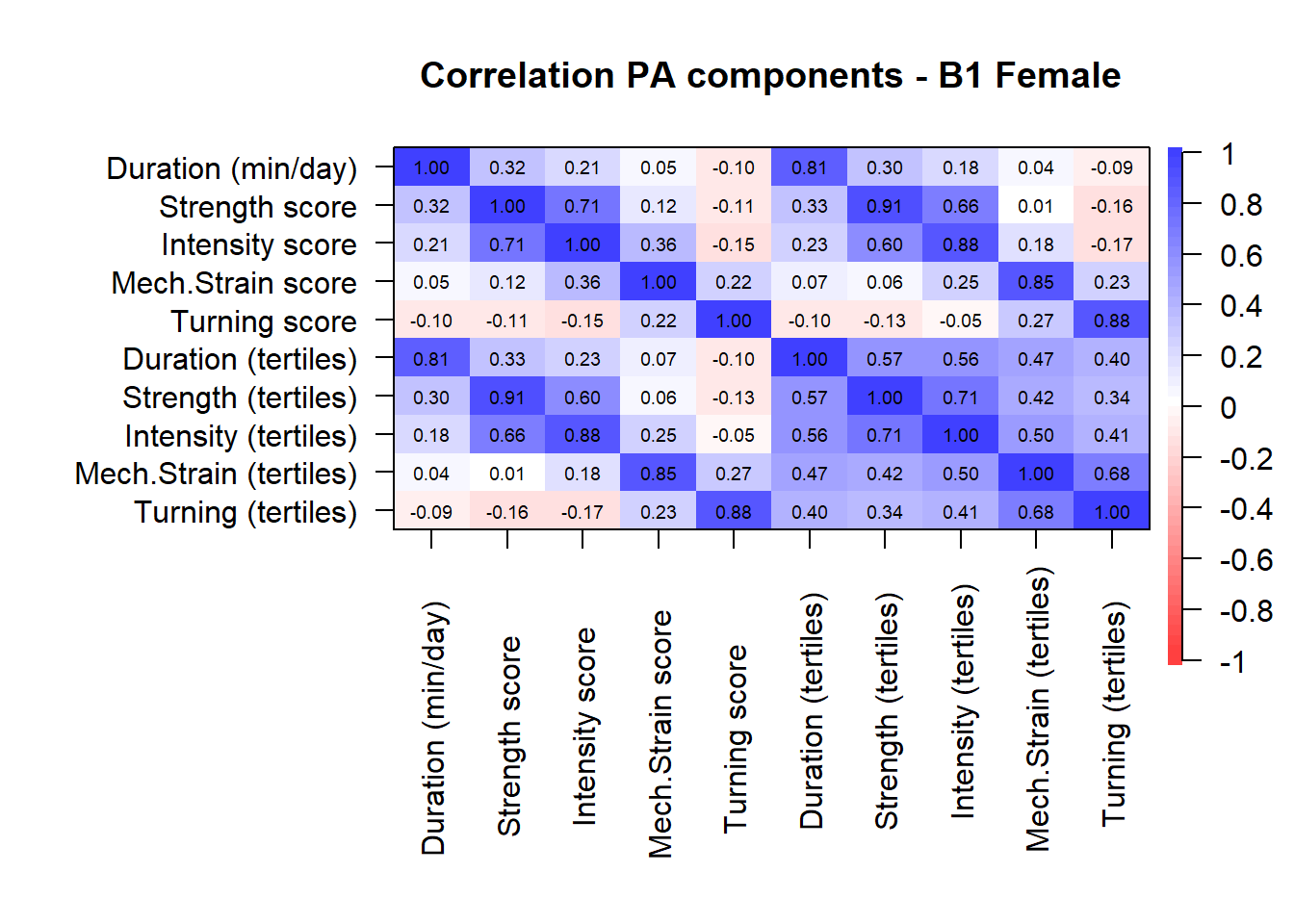

**Note:** The scales of the component scores differed between duration (range 0-960), strength (range 1-4), intensity (range 2.5 – 6.5), mechanical strain (range 1-3), turning actions (range 1-3).

### Appendix D – PA components over time

#### Figure D1 – Duration component

| A) 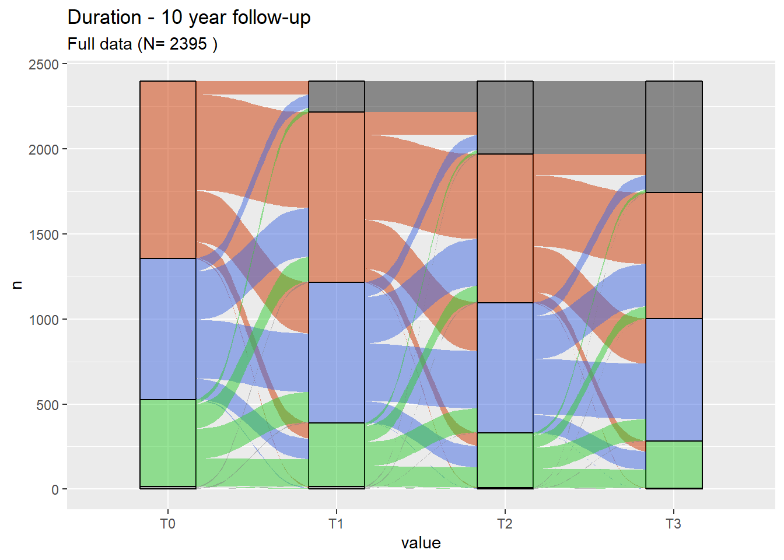Men | B) 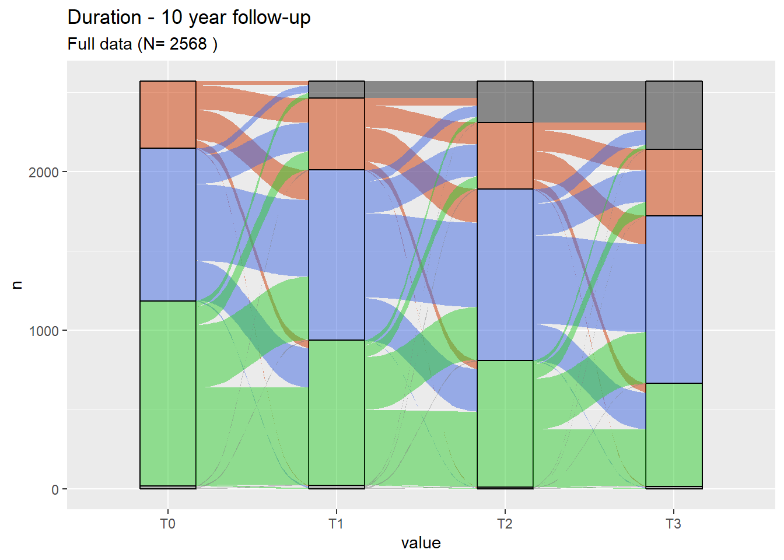Women |
| --- | --- |

#### Figure D2 – Strength component in the sample of men

| A) 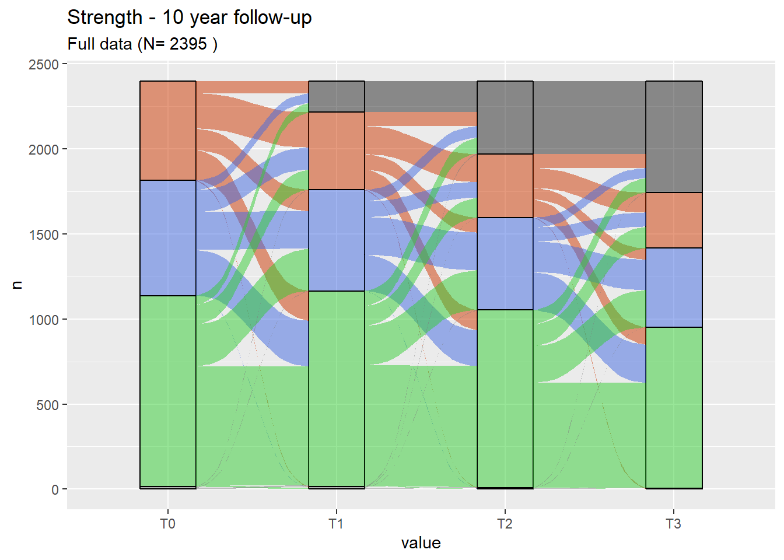Men | B) 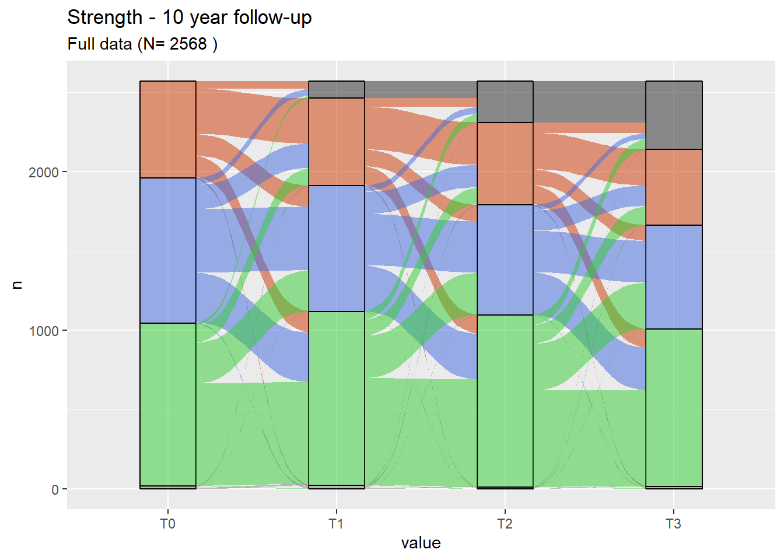Women |
| --- | --- |

#### Figure D3 – Intensity component in the ample of women

| A) 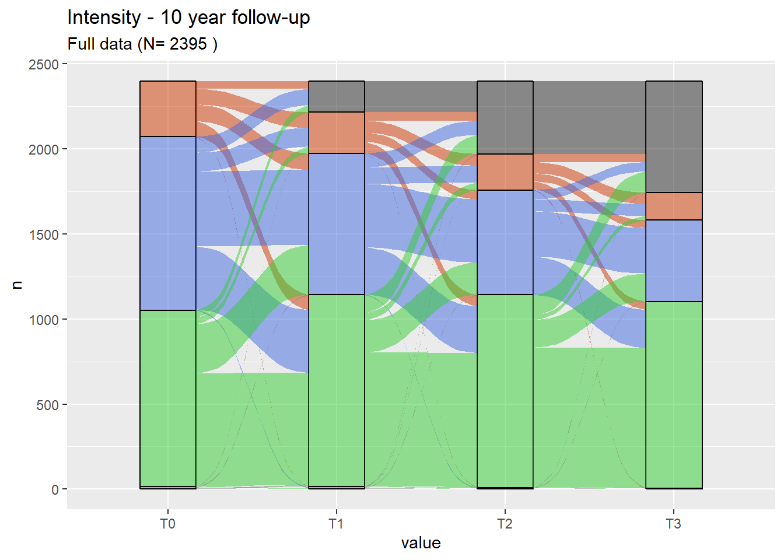Men | B) 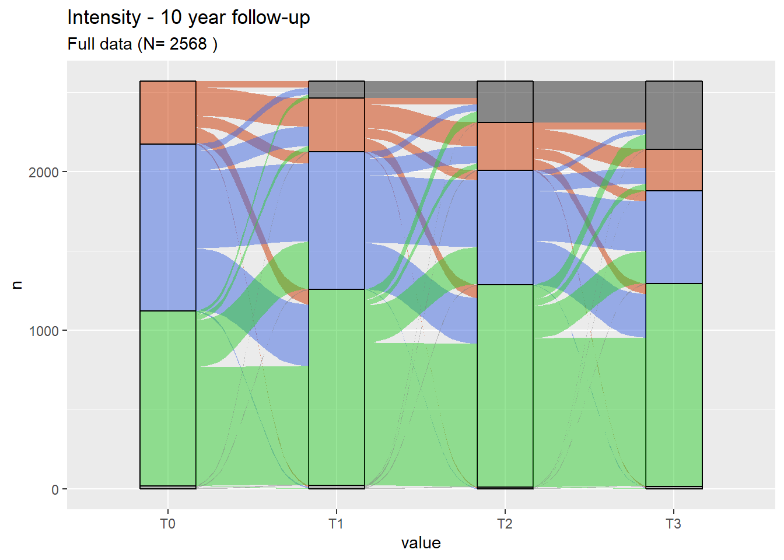Women |
| --- | --- |

#### Figure D4 – Mechanical Strain component

| A) 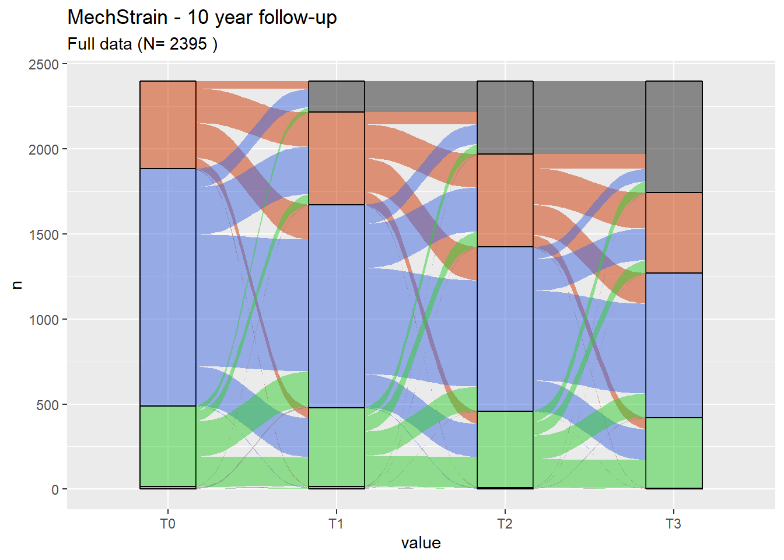Men | B) 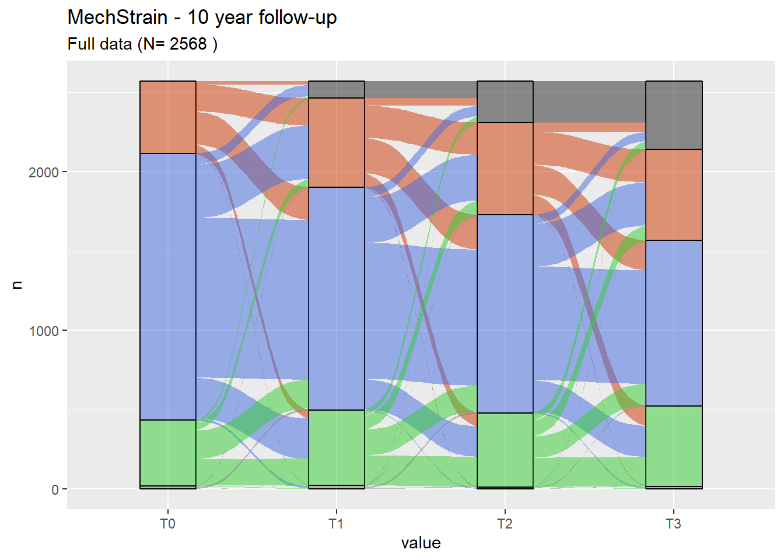Women |
| --- | --- |

#### Figure D5 – Turning Actions component

| A) 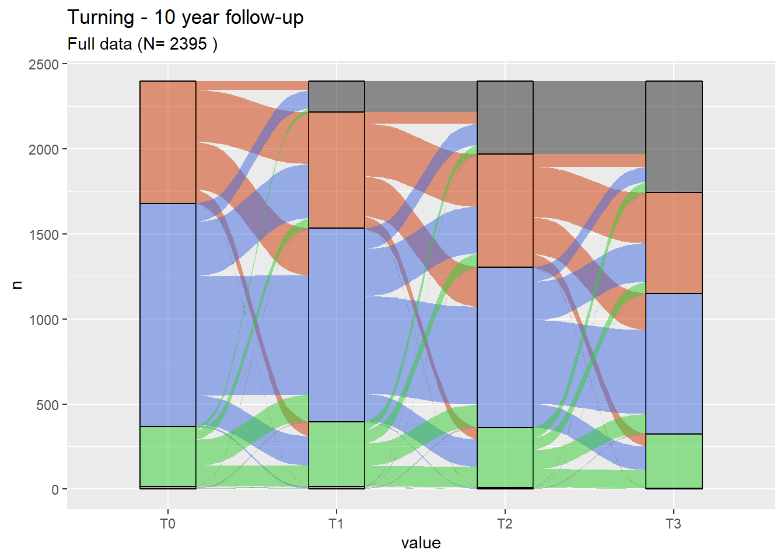Men | B) 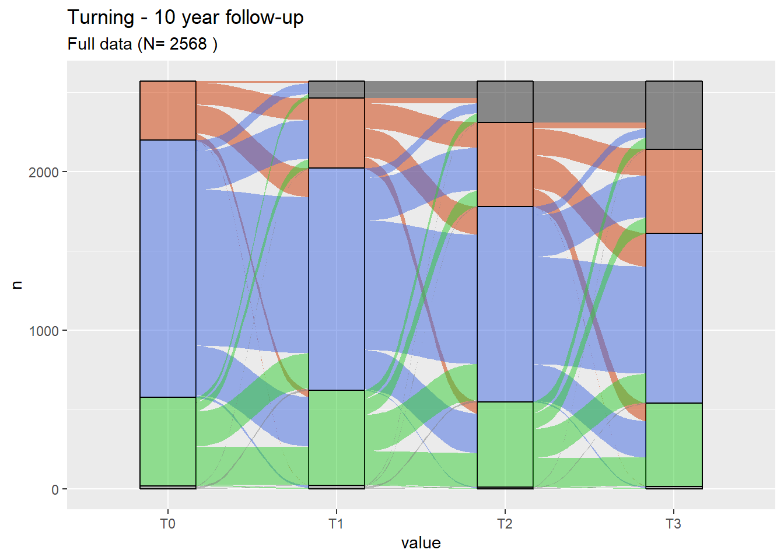Women |
| --- | --- |

### Appendix E – Overview of all component trajectory clusters

#### Figure E1 – Trajectories in the sample of men with 20 years data

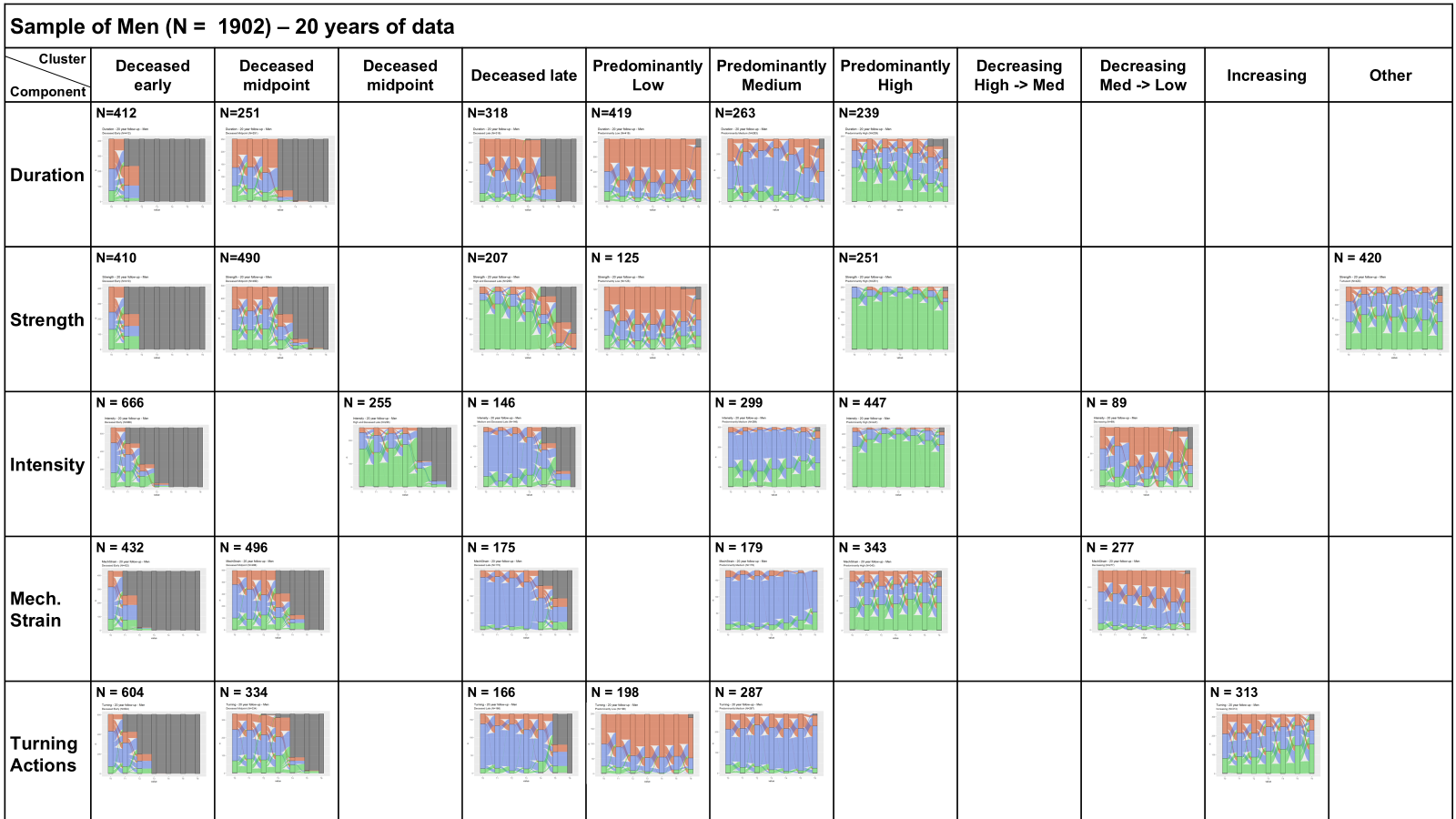

#### Figure E2 – Trajectories in the sample of women with 20 years data

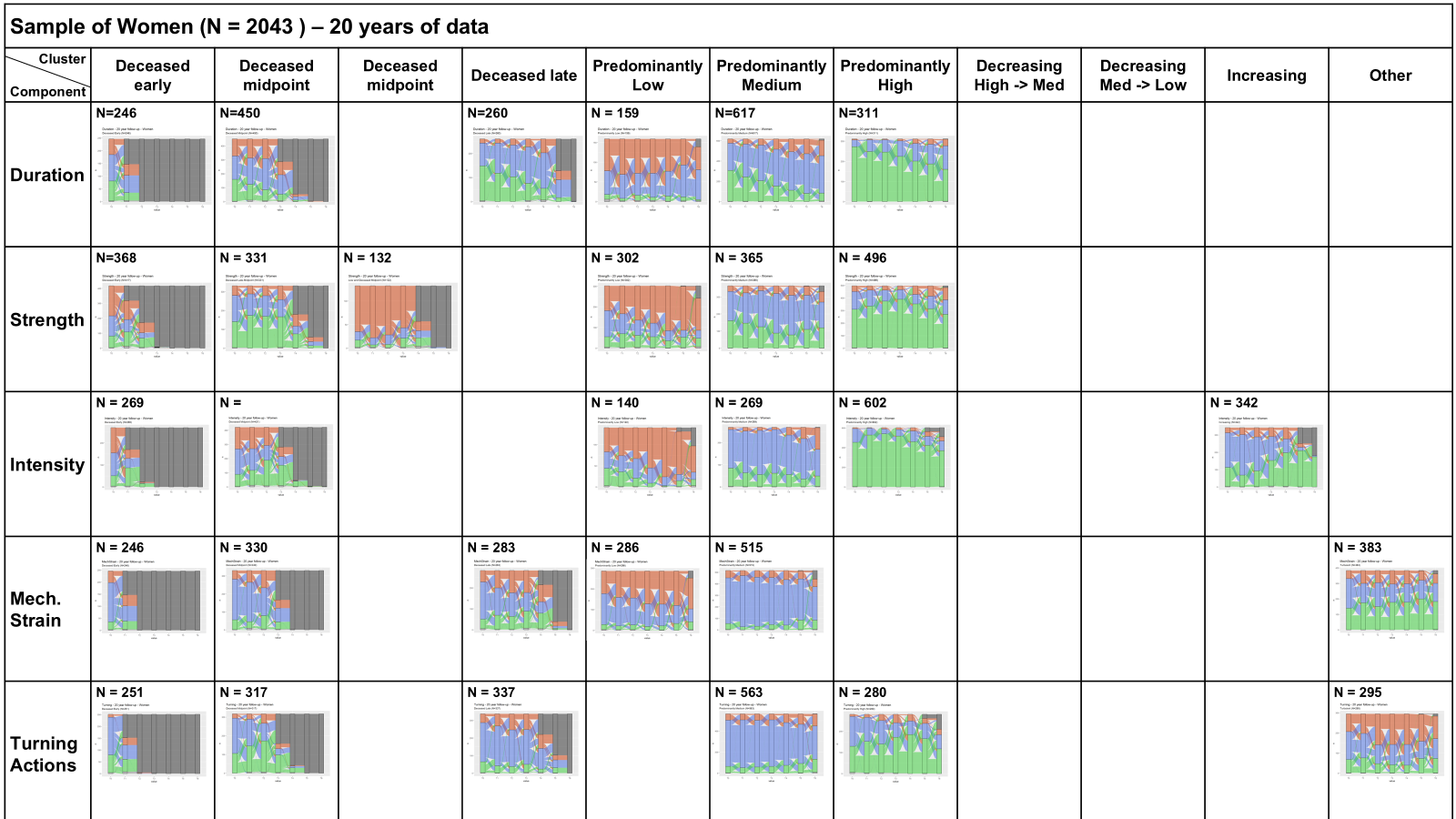

#### Figure E3 – Trajectories in the sample of men with 30 years data

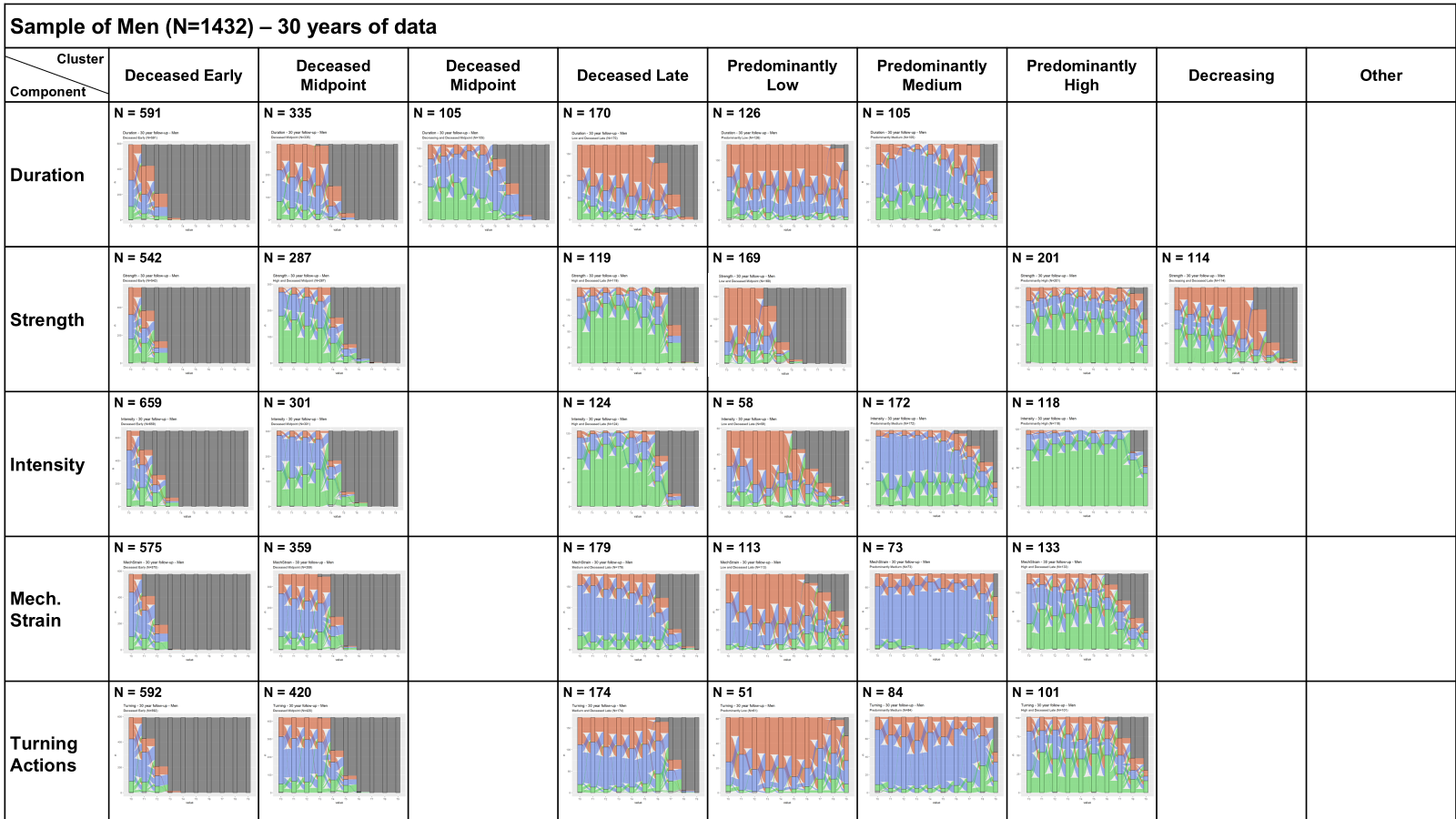

#### Figure E4 – Trajectories in the sample of women with 30 years data

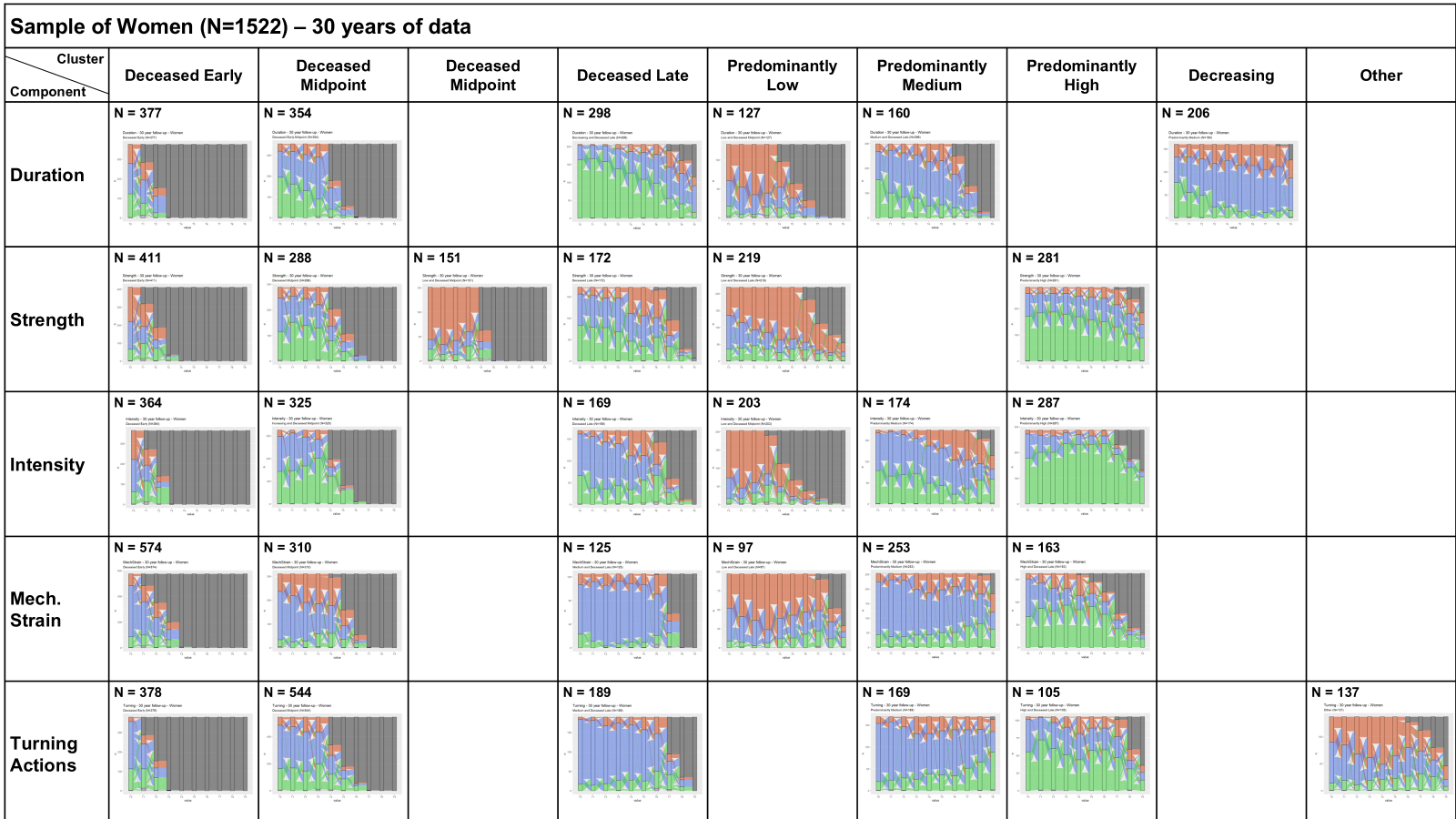

#### Figure E5 – Trajectories in the complete cases of men with 10 years data

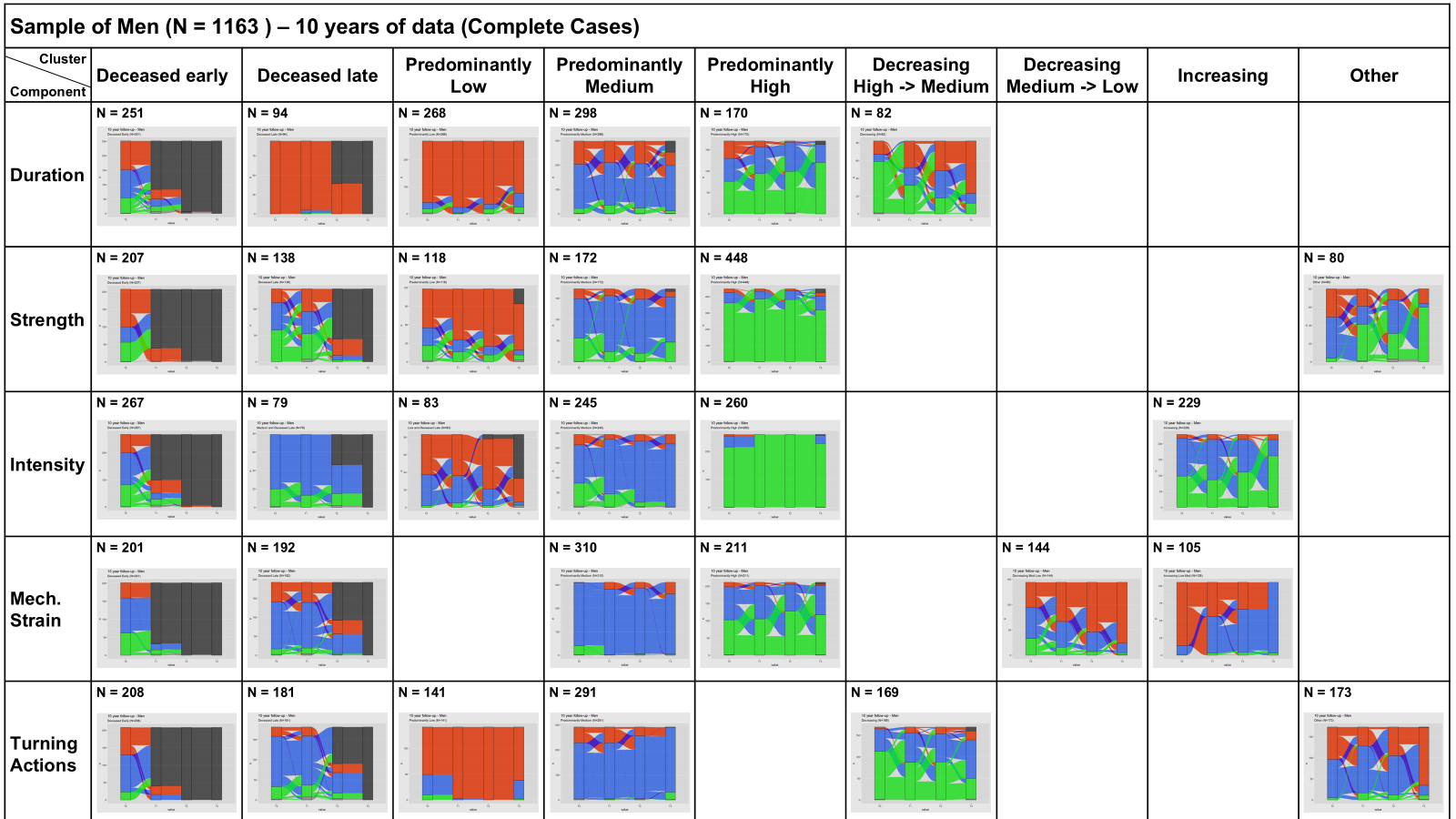

#### Figure E6 – Trajectories in the complete cases of women with 10 years data

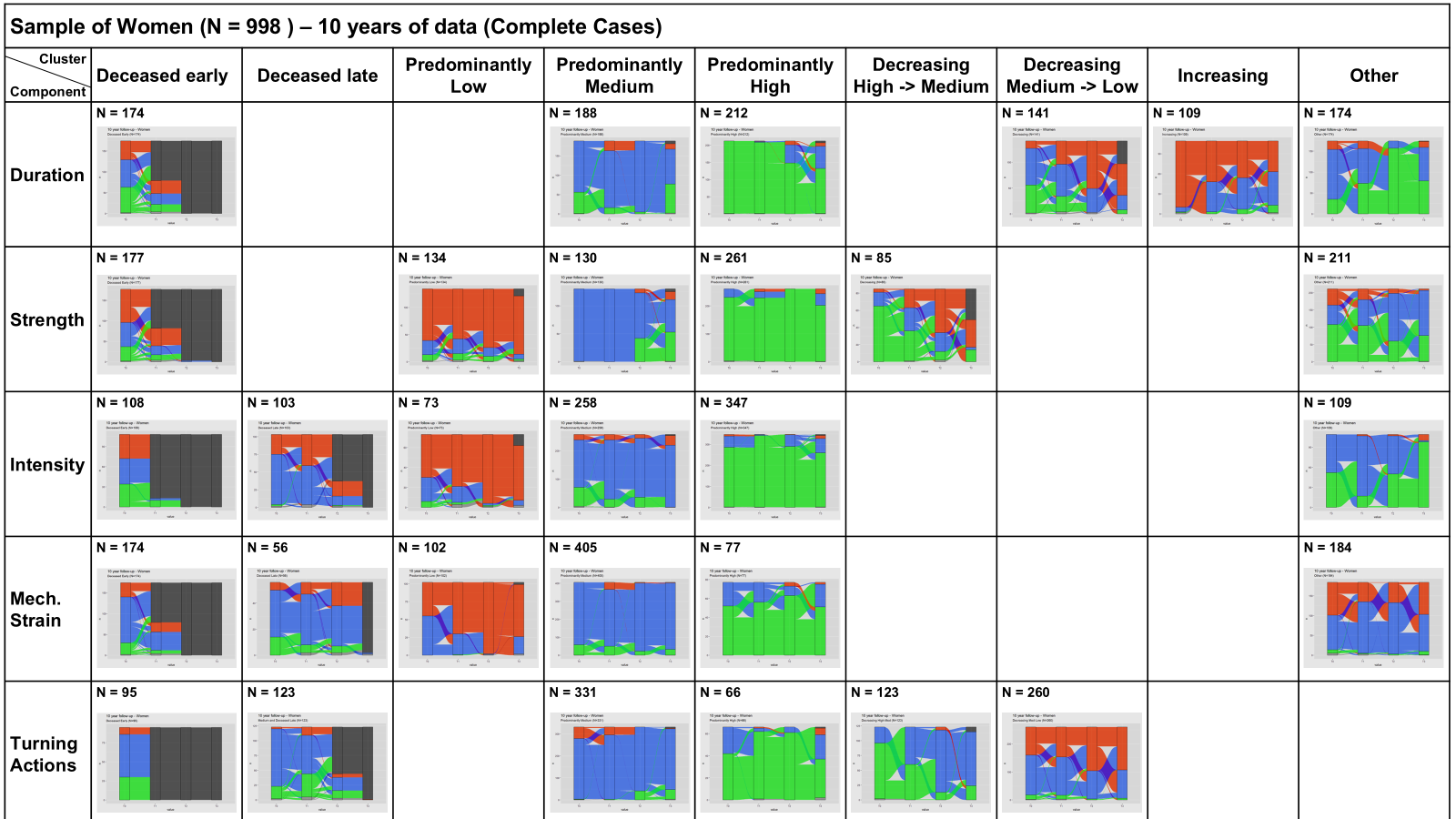

#### Figure E7 – Trajectories in the male survivors with 10 years data

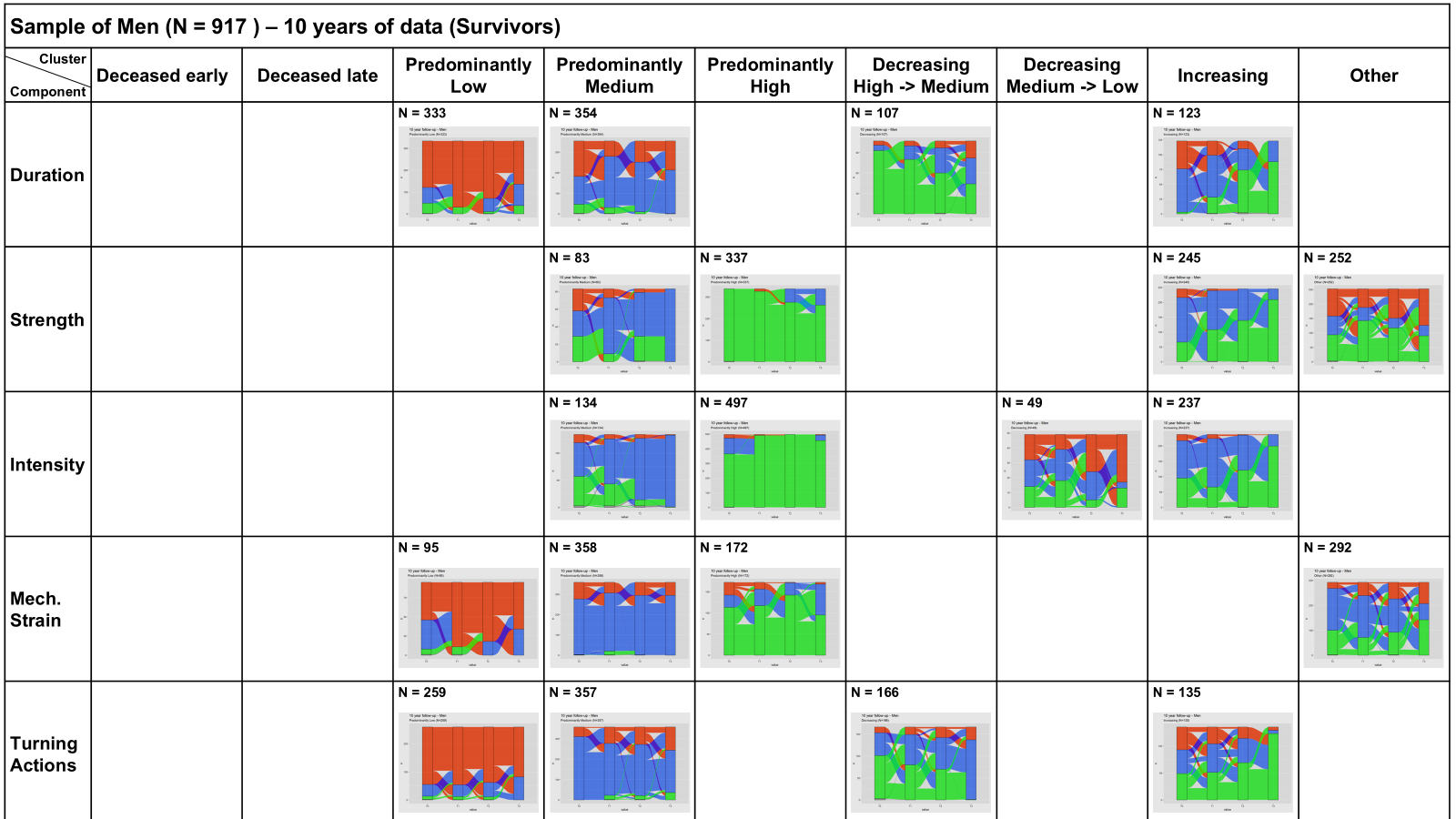

#### Figure E8 – Trajectories in the female survivors with 10 years data

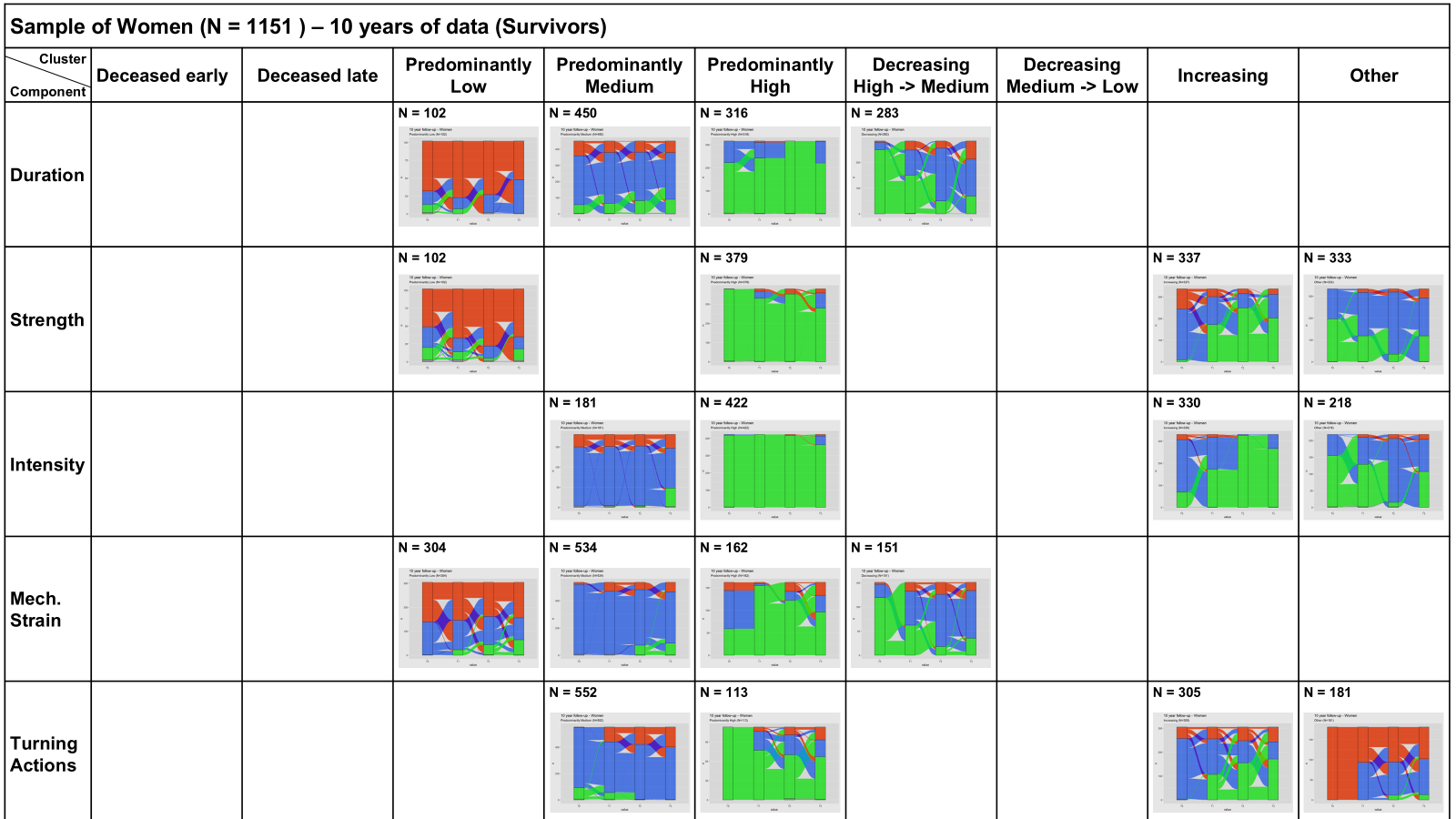

### Appendix F – Comparisons of all component trajectory groups

All p-values are calculated across the groups, and excluded the "Overall" column. All p-values are adjusted for multiple testing with a Bonferoni correction.

#### Table F1 – Intensity trajectories in the sample of Men

|  |  | **Intensity trajectory clusters** | | | | | |  | |
| --- | --- | --- | --- | --- | --- | --- | --- | --- | --- |
| Men | **Overall (N=2395)** | **Deceased Early (N=259)** | **Deceased Late (N=337)** | **Predominantly Low (N=164)** | **Predominantly Medium (N=600)** | **Predominantly High (N=524)** | **Increasing Med 🡪 High  (N=511)** | **Adjusted p-value** | |
| **Recruitment cohort** |  |  |  |  |  |  |  |  |  |
| Cohort 1 (1992) | 1432 (59.8%) | 239 (92.3%) | 291 (86.4%) | 137 (83.5%) | 375 (62.5%) | 176 (33.6%) | 214 (41.9%) | <0.001 | Chi-Square |
| Cohort 2 (2002) | 470 (19.6%) | 15 (5.8%) | 23 (6.8%) | 17 (10.4%) | 128 (21.3%) | 158 (30.2%) | 129 (25.2%) |  |  |
| Cohort 3 (2012) | 493 (20.6%) | 5 (1.9%) | 23 (6.8%) | 10 (6.1%) | 97 (16.2%) | 190 (36.3%) | 168 (32.9%) |  |  |
| **Years participating in the study** |  |  |  |  |  |  |  |  |  |
| Mean (sd) | 9.95 (7.84) | 0.791 (1.36) | 3.43 (2.24) | 11.6 (5.69) | 15.2 (6.41) | 11.9 (7.67) | 10.2 (7.59) | <0.001 | ANOVA |
| **Age at baseline** |  |  |  |  |  |  |  |  |  |
| Mean (sd) | 66.4 (8.68) | 75.3 (8.09) | 73.0 (8.42) | 71.0 (9.40) | 64.2 (6.94) | 61.8 (5.31) | 63.4 (6.93) | <0.001 | ANOVA |
| **Partner status** |  |  |  |  |  |  |  |  |  |
| No partner | 375 (15.7%) | 73 (28.2%) | 74 (22.0%) | 35 (21.3%) | 91 (15.2%) | 35 (6.7%) | 67 (13.1%) | <0.001 | Chi-Square |
| Partner (co-residence) | 1924 (80.3%) | 181 (69.9%) | 250 (74.2%) | 120 (73.2%) | 484 (80.7%) | 469 (89.5%) | 420 (82.2%) |  |  |
| Partner (outside household) | 95 (4.0%) | 5 (1.9%) | 13 (3.9%) | 9 (5.5%) | 25 (4.2%) | 20 (3.8%) | 23 (4.5%) |  |  |
| **Educational level** |  |  |  |  |  |  |  |  |  |
| Lower Education | 1110 (46.3%) | 159 (61.4%) | 204 (60.5%) | 78 (47.6%) | 271 (45.2%) | 163 (31.1%) | 235 (46.0%) | <0.001 | Chi-Square |
| Intermediate Education | 732 (30.6%) | 62 (23.9%) | 77 (22.8%) | 53 (32.3%) | 197 (32.8%) | 174 (33.2%) | 169 (33.1%) |  |  |
| Higher Education | 553 (23.1%) | 38 (14.7%) | 56 (16.6%) | 33 (20.1%) | 132 (22.0%) | 187 (35.7%) | 107 (20.9%) |  |  |
| **Number of Sports** |  |  |  |  |  |  |  |  |  |
| 0 | 1028 (42.9%) | 157 (60.6%) | 191 (56.7%) | 81 (49.4%) | 304 (50.7%) | 46 (8.8%) | 249 (48.7%) | <0.001 | Chi-Square |
| 1 | 700 (29.2%) | 69 (26.6%) | 82 (24.3%) | 51 (31.1%) | 150 (25.0%) | 196 (37.4%) | 152 (29.7%) |  |  |
| 2+ | 667 (27.8%) | 33 (12.7%) | 64 (19.0%) | 32 (19.5%) | 146 (24.3%) | 282 (53.8%) | 110 (21.5%) |  |  |
| **Total PA duration: mean min/day** |  |  |  |  |  |  |  |  |  |
| Mean (sd) | 134 (102) | 107 (86.3) | 132 (103) | 126 (105) | 143 (103) | 148 (106) | 129 (98.9) | <0.001 | ANOVA |
| **PA component: Duration** |  |  |  |  |  |  |  |  |  |
| Long duration | 512 (21.4%) | 32 (12.4%) | 66 (19.6%) | 30 (18.3%) | 135 (22.5%) | 140 (26.7%) | 109 (21.3%) | <0.001 | Chi-Square |
| Medium duration | 830 (34.7%) | 100 (38.6%) | 108 (32.0%) | 48 (29.3%) | 227 (37.8%) | 182 (34.7%) | 165 (32.3%) |  |  |
| Short duration | 1039 (43.4%) | 127 (49.0%) | 160 (47.5%) | 86 (52.4%) | 232 (38.7%) | 199 (38.0%) | 235 (46.0%) |  |  |
| **PA component: Muscle Strength** |  |  |  |  |  |  |  |  |  |
| High Strength | 1123 (46.9%) | 63 (24.3%) | 137 (40.7%) | 24 (14.6%) | 296 (49.3%) | 364 (69.5%) | 239 (46.8%) | <0.001 | Chi-Square |
| Medium Strength | 676 (28.2%) | 71 (27.4%) | 109 (32.3%) | 38 (23.2%) | 185 (30.8%) | 136 (26.0%) | 137 (26.8%) |  |  |
| Low Strength | 582 (24.3%) | 125 (48.3%) | 88 (26.1%) | 102 (62.2%) | 113 (18.8%) | 21 (4.0%) | 133 (26.0%) |  |  |
| **PA component: Intensity** |  |  |  |  |  |  |  |  |  |
| High Intensity | 1036 (43.3%) | 52 (20.1%) | 97 (28.8%) | 16 (9.8%) | 203 (33.8%) | 519 (99.0%) | 149 (29.2%) | 0.0085 | Fisher's exact |
| Medium Intensity | 1020 (42.6%) | 112 (43.2%) | 214 (63.5%) | 54 (32.9%) | 355 (59.2%) | 2 (0.4%) | 283 (55.4%) |  |  |
| Low Intensity | 325 (13.6%) | 95 (36.7%) | 23 (6.8%) | 94 (57.3%) | 36 (6.0%) | 0 (0%) | 77 (15.1%) |  |  |
| **PA component: Mechanical Strain** |  |  |  |  |  |  |  |  |  |
| High Mech.Strain | 474 (19.8%) | 40 (15.4%) | 67 (19.9%) | 25 (15.2%) | 97 (16.2%) | 159 (30.3%) | 86 (16.8%) | <0.001 | Chi-Square |
| Medium Mech.Strain | 1393 (58.2%) | 155 (59.8%) | 177 (52.5%) | 108 (65.9%) | 372 (62.0%) | 249 (47.5%) | 332 (65.0%) |  |  |
| Low Mech.Strain | 514 (21.5%) | 64 (24.7%) | 90 (26.7%) | 31 (18.9%) | 125 (20.8%) | 113 (21.6%) | 91 (17.8%) |  |  |
| **PA component: Turning Actions** |  |  |  |  |  |  |  |  |  |
| High Turning | 355 (14.8%) | 41 (15.8%) | 44 (13.1%) | 28 (17.1%) | 68 (11.3%) | 116 (22.1%) | 58 (11.4%) | <0.001 | Chi-Square |
| Medium Turning | 1309 (54.7%) | 151 (58.3%) | 192 (57.0%) | 106 (64.6%) | 359 (59.8%) | 195 (37.2%) | 306 (59.9%) |  |  |
| Low Turning | 717 (29.9%) | 67 (25.9%) | 98 (29.1%) | 30 (18.3%) | 167 (27.8%) | 210 (40.1%) | 145 (28.4%) |  |  |
| **Functional limitation scale[0-9]** |  |  |  |  |  |  |  |  |  |
| Median [Q1, Q3] | 0 [0, 1.00] | 1.00 [0, 3.00] | 0 [0, 2.00] | 0 [0, 2.00] | 0 [0, 0] | 0 [0, 0] | 0 [0, 1.00] | <0.001 | Kruskal-Wallis |
| **MMSE-score based on maximum spel/num** |  |  |  |  |  |  |  |  |  |
| Mean (sd) | 27.4 (2.53) | 26.0 (3.81) | 26.4 (2.98) | 27.5 (2.02) | 27.9 (2.02) | 28.1 (1.62) | 27.6 (2.35) | <0.001 | ANOVA |
| **Number of Chronic comorbidities** |  |  |  |  |  |  |  |  |  |
| Median [Q1, Q3] | 1.00 [0, 2.00] | 2.00 [1.00, 3.00] | 1.00 [1.00, 2.00] | 1.50 [1.00, 2.00] | 1.00 [0, 2.00] | 1.00 [0, 2.00] | 1.00 [0, 2.00] | <0.001 | Kruskal-Wallis |
| **Health problems limit normal activities** |  |  |  |  |  |  |  |  |  |
| Yes, severely | 255 (10.6%) | 64 (24.7%) | 46 (13.6%) | 26 (15.9%) | 51 (8.5%) | 21 (4.0%) | 47 (9.2%) | <0.001 | Chi-Square |
| Yes, slightly | 435 (18.2%) | 59 (22.8%) | 59 (17.5%) | 32 (19.5%) | 107 (17.8%) | 84 (16.0%) | 94 (18.4%) |  |  |
| No | 1699 (70.9%) | 135 (52.1%) | 230 (68.2%) | 106 (64.6%) | 441 (73.5%) | 419 (80.0%) | 368 (72.0%) |  |  |
| **Self-perceived health** |  |  |  |  |  |  |  |  |  |
| Excellent | 365 (15.2%) | 17 (6.6%) | 47 (13.9%) | 16 (9.8%) | 93 (15.5%) | 107 (20.4%) | 85 (16.6%) | 0.0085 | Fisher's exact |
| Good | 1265 (52.8%) | 113 (43.6%) | 179 (53.1%) | 78 (47.6%) | 310 (51.7%) | 319 (60.9%) | 266 (52.1%) |  |  |
| Fair | 502 (21.0%) | 76 (29.3%) | 72 (21.4%) | 44 (26.8%) | 140 (23.3%) | 70 (13.4%) | 100 (19.6%) |  |  |
| Sometimes good/bad | 196 (8.2%) | 37 (14.3%) | 22 (6.5%) | 17 (10.4%) | 47 (7.8%) | 25 (4.8%) | 48 (9.4%) |  |  |
| Poor | 61 (2.5%) | 15 (5.8%) | 15 (4.5%) | 8 (4.9%) | 9 (1.5%) | 3 (0.6%) | 11 (2.2%) |  |  |

#### Table F2 – Intensity trajectories in the sample of Women

|  |  | **Intensity trajectory clusters** | | | | | |  | |
| --- | --- | --- | --- | --- | --- | --- | --- | --- | --- |
| Women | **Overall (N=2568)** | **Deceased Early (N=209)** | **Deceased Late (N=141)** | **Predominantly Low (N=217)** | **Predominantly Medium (N=548)** | **Predominantly High (N=1117)** | **Increasing Med 🡪 High (N=336)** | **Adjusted p-value** | |
| **Recruitment cohort** |  |  |  |  |  |  |  |  |  |
| Cohort 1 (1992) | 1522 (59.3%) | 177 (84.7%) | 134 (95.0%) | 181 (83.4%) | 334 (60.9%) | 482 (43.2%) | 214 (63.7%) | 0.0085 | Fisher's exact |
| Cohort 2 (2002) | 521 (20.3%) | 21 (10.0%) | 3 (2.1%) | 25 (11.5%) | 125 (22.8%) | 292 (26.1%) | 55 (16.4%) |  |  |
| Cohort 3 (2012) | 525 (20.4%) | 11 (5.3%) | 4 (2.8%) | 11 (5.1%) | 89 (16.2%) | 343 (30.7%) | 67 (19.9%) |  |  |
| **Years participating in the study** |  |  |  |  |  |  |  |  |  |
| Mean (sd) | 11.6 (8.23) | 1.18 (1.54) | 3.88 (2.21) | 13.5 (5.20) | 16.2 (7.09) | 11.5 (8.27) | 12.5 (7.51) | <0.001 | ANOVA |
| **Age at baseline** |  |  |  |  |  |  |  |  |  |
| Mean (sd) | 66.1 (8.55) | 73.5 (8.83) | 78.6 (6.16) | 71.9 (8.74) | 63.8 (6.73) | 63.1 (6.64) | 66.3 (8.15) | <0.001 | ANOVA |
| **Partner status** |  |  |  |  |  |  |  |  |  |
| No partner | 940 (36.6%) | 121 (57.9%) | 100 (70.9%) | 117 (53.9%) | 184 (33.6%) | 293 (26.2%) | 125 (37.2%) | 0.0085 | Fisher's exact |
| Partner (co-residence) | 1559 (60.7%) | 87 (41.6%) | 38 (27.0%) | 94 (43.3%) | 341 (62.2%) | 797 (71.4%) | 202 (60.1%) |  |  |
| Partner (outside household) | 69 (2.7%) | 1 (0.5%) | 3 (2.1%) | 6 (2.8%) | 23 (4.2%) | 27 (2.4%) | 9 (2.7%) |  |  |
| **Educational level** |  |  |  |  |  |  |  |  |  |
| Lower Education | 1441 (56.1%) | 153 (73.2%) | 106 (75.2%) | 137 (63.1%) | 305 (55.7%) | 536 (48.0%) | 204 (60.7%) | <0.001 | Chi-Square |
| Intermediate Education | 796 (31.0%) | 40 (19.1%) | 24 (17.0%) | 47 (21.7%) | 176 (32.1%) | 408 (36.5%) | 101 (30.1%) |  |  |
| Higher Education | 328 (12.8%) | 16 (7.7%) | 11 (7.8%) | 32 (14.7%) | 67 (12.2%) | 171 (15.3%) | 31 (9.2%) |  |  |
| **Number of Sports** |  |  |  |  |  |  |  |  |  |
| 0 | 1015 (39.5%) | 114 (54.5%) | 81 (57.4%) | 131 (60.4%) | 235 (42.9%) | 249 (22.3%) | 205 (61.0%) | <0.001 | Chi-Square |
| 1 | 811 (31.6%) | 61 (29.2%) | 55 (39.0%) | 66 (30.4%) | 169 (30.8%) | 371 (33.2%) | 89 (26.5%) |  |  |
| 2+ | 742 (28.9%) | 34 (16.3%) | 5 (3.5%) | 20 (9.2%) | 144 (26.3%) | 497 (44.5%) | 42 (12.5%) |  |  |
| **Total PA duration: mean min/day** |  |  |  |  |  |  |  |  |  |
| Mean (sd) | 199 (111) | 180 (104) | 130 (84.5) | 147 (91.1) | 214 (113) | 210 (107) | 211 (125) | <0.001 | ANOVA |
| **PA component: Duration** |  |  |  |  |  |  |  |  |  |
| Long duration | 1165 (45.4%) | 77 (36.8%) | 32 (22.7%) | 58 (26.7%) | 275 (50.2%) | 560 (50.1%) | 163 (48.5%) | <0.001 | Chi-Square |
| Medium duration | 962 (37.5%) | 90 (43.1%) | 50 (35.5%) | 78 (35.9%) | 206 (37.6%) | 424 (38.0%) | 114 (33.9%) |  |  |
| Short duration | 422 (16.4%) | 39 (18.7%) | 58 (41.1%) | 75 (34.6%) | 63 (11.5%) | 130 (11.6%) | 57 (17.0%) |  |  |
| **PA component: Muscle Strength** |  |  |  |  |  |  |  |  |  |
| High Strength | 1025 (39.9%) | 46 (22.0%) | 8 (5.7%) | 22 (10.1%) | 273 (49.8%) | 559 (50.0%) | 117 (34.8%) | <0.001 | Chi-Square |
| Medium Strength | 915 (35.6%) | 84 (40.2%) | 17 (12.1%) | 36 (16.6%) | 206 (37.6%) | 411 (36.8%) | 161 (47.9%) |  |  |
| Low Strength | 609 (23.7%) | 76 (36.4%) | 115 (81.6%) | 153 (70.5%) | 65 (11.9%) | 144 (12.9%) | 56 (16.7%) |  |  |
| **PA component: Intensity** |  |  |  |  |  |  |  |  |  |
| High Intensity | 1102 (42.9%) | 47 (22.5%) | 1 (0.7%) | 21 (9.7%) | 232 (42.3%) | 801 (71.7%) | 0 (0%) | 0.0085 | Fisher's exact |
| Medium Intensity | 1053 (41.0%) | 111 (53.1%) | 25 (17.7%) | 50 (23.0%) | 297 (54.2%) | 236 (21.1%) | 334 (99.4%) |  |  |
| Low Intensity | 394 (15.3%) | 48 (23.0%) | 114 (80.9%) | 140 (64.5%) | 15 (2.7%) | 77 (6.9%) | 0 (0%) |  |  |
| **PA component: Mechanical Strain** |  |  |  |  |  |  |  |  |  |
| High Mech.Strain | 415 (16.2%) | 34 (16.3%) | 9 (6.4%) | 18 (8.3%) | 90 (16.4%) | 169 (15.1%) | 95 (28.3%) | <0.001 | Chi-Square |
| Medium Mech.Strain | 1681 (65.5%) | 137 (65.6%) | 102 (72.3%) | 148 (68.2%) | 367 (67.0%) | 716 (64.1%) | 211 (62.8%) |  |  |
| Low Mech.Strain | 453 (17.6%) | 35 (16.7%) | 29 (20.6%) | 45 (20.7%) | 87 (15.9%) | 229 (20.5%) | 28 (8.3%) |  |  |
| **PA component: Turning Actions** |  |  |  |  |  |  |  |  |  |
| High Turning | 558 (21.7%) | 55 (26.3%) | 43 (30.5%) | 61 (28.1%) | 99 (18.1%) | 219 (19.6%) | 81 (24.1%) | 0.0085 | Fisher's exact |
| Medium Turning | 1621 (63.1%) | 137 (65.6%) | 93 (66.0%) | 134 (61.8%) | 362 (66.1%) | 663 (59.4%) | 232 (69.0%) |  |  |
| Low Turning | 370 (14.4%) | 14 (6.7%) | 4 (2.8%) | 16 (7.4%) | 83 (15.1%) | 232 (20.8%) | 21 (6.3%) |  |  |
| **Functional limitation scale[0-9]** |  |  |  |  |  |  |  |  |  |
| Median [Q1, Q3] | 0 [0, 2.00] | 1.00 [0, 5.00] | 4.00 [2.00, 7.00] | 2.00 [0.750, 5.00] | 0 [0, 1.00] | 0 [0, 1.00] | 0 [0, 2.00] | <0.001 | Kruskal-Wallis |
| **MMSE-score based on maximum spel/num** |  |  |  |  |  |  |  |  |  |
| Mean (sd) | 27.4 (2.62) | 26.0 (3.57) | 25.5 (3.61) | 27.1 (2.45) | 28.0 (1.92) | 27.7 (2.52) | 27.3 (2.15) | <0.001 | ANOVA |
| **Number of Chronic comorbidities** |  |  |  |  |  |  |  |  |  |
| Median [Q1, Q3] | 2.00 [1.00, 3.00] | 2.00 [1.00, 3.00] | 2.00 [1.00, 3.00] | 2.00 [1.00, 3.00] | 1.00 [1.00, 3.00] | 1.00 [0, 2.00] | 2.00 [1.00, 3.00] | <0.001 | Kruskal-Wallis |
| **Health problems limit normal activities** |  |  |  |  |  |  |  |  |  |
| Yes, severely | 275 (10.7%) | 36 (17.2%) | 34 (24.1%) | 54 (24.9%) | 40 (7.3%) | 79 (7.1%) | 32 (9.5%) | <0.001 | Chi-Square |
| Yes, slightly | 581 (22.6%) | 65 (31.1%) | 42 (29.8%) | 65 (30.0%) | 119 (21.7%) | 216 (19.3%) | 74 (22.0%) |  |  |
| No | 1707 (66.5%) | 107 (51.2%) | 65 (46.1%) | 97 (44.7%) | 388 (70.8%) | 820 (73.4%) | 230 (68.5%) |  |  |
| **Self-perceived health** |  |  |  |  |  |  |  |  |  |
| Excellent | 347 (13.5%) | 10 (4.8%) | 12 (8.5%) | 14 (6.5%) | 86 (15.7%) | 185 (16.6%) | 40 (11.9%) | 0.0085 | Fisher's exact |
| Good | 1247 (48.6%) | 91 (43.5%) | 53 (37.6%) | 73 (33.6%) | 268 (48.9%) | 596 (53.4%) | 166 (49.4%) |  |  |
| Fair | 601 (23.4%) | 70 (33.5%) | 43 (30.5%) | 68 (31.3%) | 118 (21.5%) | 216 (19.3%) | 86 (25.6%) |  |  |
| Sometimes good/bad | 309 (12.0%) | 26 (12.4%) | 25 (17.7%) | 45 (20.7%) | 65 (11.9%) | 107 (9.6%) | 41 (12.2%) |  |  |
| Poor | 56 (2.2%) | 12 (5.7%) | 6 (4.3%) | 16 (7.4%) | 9 (1.6%) | 10 (0.9%) | 3 (0.9%) |  |  |

#### Table F3 – Strength trajectories in the sample of Men

|  |  | **Strength trajectory clusters** | | | | | |  | | |
| --- | --- | --- | --- | --- | --- | --- | --- | --- | --- | --- |
| Men | **Overall (N=2395)** | **Deceased Early (N=182)** | **Deceased Late (N=450)** | **Predominantly Low (N=214)** | **Predominantly Medium (N=348)** | **Predominantly High (N=974)** | **Increasing Low 🡪 High (N=227)** | **Adjusted p-value** |  | |
| Cohort 1 (1992) | 1432 (59.8%) | 166 (91.2%) | 398 (88.4%) | 148 (69.2%) | 170 (48.9%) | 443 (45.5%) | 107 (47.1%) | 0.0085 | | Fisher's exact |
| Cohort 2 (2002) | 470 (19.6%) | 12 (6.6%) | 29 (6.4%) | 36 (16.8%) | 76 (21.8%) | 254 (26.1%) | 63 (27.8%) |  | |  |
| Cohort 3 (2012) | 493 (20.6%) | 4 (2.2%) | 23 (5.1%) | 30 (14.0%) | 102 (29.3%) | 277 (28.4%) | 57 (25.1%) |  | |  |
| **Years participating in the study** |  |  |  |  |  |  |  |  | |  |
| Mean (sd) | 9.95 (7.84) | 0 (0) | 3.47 (2.14) | 13.0 (5.71) | 12.0 (6.96) | 13.0 (7.67) | 11.8 (7.86) | <0.001 | | ANOVA |
| **Age at baseline** |  |  |  |  |  |  |  |  | |  |
| Mean (sd) | 66.4 (8.68) | 74.9 (8.43) | 74.0 (8.20) | 68.1 (9.05) | 63.6 (6.74) | 62.7 (5.99) | 63.3 (6.92) | <0.001 | | ANOVA |
| **Partner status** |  |  |  |  |  |  |  |  | |  |
| No partner | 375 (15.7%) | 51 (28.0%) | 111 (24.7%) | 45 (21.0%) | 47 (13.5%) | 87 (8.9%) | 34 (15.0%) | 0.0085 | | Fisher's exact |
| Partner (co-residence) | 1924 (80.3%) | 127 (69.8%) | 325 (72.2%) | 154 (72.0%) | 279 (80.2%) | 855 (87.8%) | 184 (81.1%) |  | |  |
| Partner (outside household) | 95 (4.0%) | 4 (2.2%) | 14 (3.1%) | 15 (7.0%) | 22 (6.3%) | 31 (3.2%) | 9 (4.0%) |  | |  |
| **Educational level** |  |  |  |  |  |  |  |  | |  |
| Lower Education | 1110 (46.3%) | 118 (64.8%) | 258 (57.3%) | 103 (48.1%) | 126 (36.2%) | 398 (40.9%) | 107 (47.1%) | <0.001 | | Chi-Square |
| Intermediate Education | 732 (30.6%) | 39 (21.4%) | 114 (25.3%) | 61 (28.5%) | 119 (34.2%) | 322 (33.1%) | 77 (33.9%) |  | |  |
| Higher Education | 553 (23.1%) | 25 (13.7%) | 78 (17.3%) | 50 (23.4%) | 103 (29.6%) | 254 (26.1%) | 43 (18.9%) |  | |  |
| **Number of Sports** |  |  |  |  |  |  |  |  | |  |
| 0 | 1028 (42.9%) | 113 (62.1%) | 253 (56.2%) | 92 (43.0%) | 134 (38.5%) | 325 (33.4%) | 111 (48.9%) | <0.001 | | Chi-Square |
| 1 | 700 (29.2%) | 44 (24.2%) | 122 (27.1%) | 68 (31.8%) | 106 (30.5%) | 291 (29.9%) | 69 (30.4%) |  | |  |
| 2+ | 667 (27.8%) | 25 (13.7%) | 75 (16.7%) | 54 (25.2%) | 108 (31.0%) | 358 (36.8%) | 47 (20.7%) |  | |  |
| **Total PA duration: mean min/day** |  |  |  |  |  |  |  |  | |  |
| Mean (sd) | 134 (102) | 118 (92.4) | 120 (97.3) | 134 (104) | 144 (98.1) | 150 (108) | 96.9 (75.2) | <0.001 | | ANOVA |
| **PA component: Duration** |  |  |  |  |  |  |  |  | |  |
| Long duration | 512 (21.4%) | 27 (14.8%) | 74 (16.4%) | 43 (20.1%) | 85 (24.4%) | 260 (26.7%) | 23 (10.1%) | <0.001 | | Chi-Square |
| Medium duration | 830 (34.7%) | 77 (42.3%) | 142 (31.6%) | 68 (31.8%) | 133 (38.2%) | 344 (35.3%) | 66 (29.1%) |  | |  |
| Short duration | 1039 (43.4%) | 78 (42.9%) | 232 (51.6%) | 101 (47.2%) | 127 (36.5%) | 363 (37.3%) | 138 (60.8%) |  | |  |
| **PA component: Muscle Strength** |  |  |  |  |  |  |  |  | |  |
| High Strength | 1123 (46.9%) | 55 (30.2%) | 135 (30.0%) | 50 (23.4%) | 33 (9.5%) | 850 (87.3%) | 0 (0%) | 0.0085 | | Fisher's exact |
| Medium Strength | 676 (28.2%) | 56 (30.8%) | 133 (29.6%) | 69 (32.2%) | 301 (86.5%) | 117 (12.0%) | 0 (0%) |  | |  |
| Low Strength | 582 (24.3%) | 71 (39.0%) | 180 (40.0%) | 93 (43.5%) | 11 (3.2%) | 0 (0%) | 227 (100%) |  | |  |
| **PA component: Intensity** |  |  |  |  |  |  |  |  | |  |
| High Intensity | 1036 (43.3%) | 43 (23.6%) | 114 (25.3%) | 65 (30.4%) | 164 (47.1%) | 630 (64.7%) | 20 (8.8%) | <0.001 | | Chi-Square |
| Medium Intensity | 1020 (42.6%) | 94 (51.6%) | 226 (50.2%) | 100 (46.7%) | 165 (47.4%) | 323 (33.2%) | 112 (49.3%) |  | |  |
| Low Intensity | 325 (13.6%) | 45 (24.7%) | 108 (24.0%) | 47 (22.0%) | 16 (4.6%) | 14 (1.4%) | 95 (41.9%) |  | |  |
| **PA component: Mechanical Strain** |  |  |  |  |  |  |  |  | |  |
| High Mech.Strain | 474 (19.8%) | 32 (17.6%) | 81 (18.0%) | 53 (24.8%) | 75 (21.6%) | 195 (20.0%) | 38 (16.7%) | <0.001 | | Chi-Square |
| Medium Mech.Strain | 1393 (58.2%) | 108 (59.3%) | 249 (55.3%) | 106 (49.5%) | 229 (65.8%) | 586 (60.2%) | 115 (50.7%) |  | |  |
| Low Mech.Strain | 514 (21.5%) | 42 (23.1%) | 118 (26.2%) | 53 (24.8%) | 41 (11.8%) | 186 (19.1%) | 74 (32.6%) |  | |  |
| **PA component: Turning Actions** |  |  |  |  |  |  |  |  | |  |
| High Turning | 355 (14.8%) | 23 (12.6%) | 71 (15.8%) | 39 (18.2%) | 51 (14.7%) | 149 (15.3%) | 22 (9.7%) | <0.001 | | Chi-Square |
| Medium Turning | 1309 (54.7%) | 108 (59.3%) | 257 (57.1%) | 118 (55.1%) | 236 (67.8%) | 486 (49.9%) | 104 (45.8%) |  | |  |
| Low Turning | 717 (29.9%) | 51 (28.0%) | 120 (26.7%) | 55 (25.7%) | 58 (16.7%) | 332 (34.1%) | 101 (44.5%) |  | |  |
| **Functional limitation scale[0-9]** |  |  |  |  |  |  |  |  | |  |
| Median [Q1, Q3] | 0 [0, 1.00] | 1.00 [0, 3.00] | 0 [0, 2.00] | 0 [0, 1.00] | 0 [0, 0] | 0 [0, 0] | 0 [0, 1.00] | <0.001 | | Kruskal-Wallis |
| **MMSE-score based on maximum spel/num** |  |  |  |  |  |  |  |  | |  |
| Mean (sd) | 27.4 (2.53) | 25.8 (4.08) | 26.5 (2.95) | 27.6 (2.04) | 28.1 (2.02) | 27.9 (1.93) | 27.5 (2.33) | <0.001 | | ANOVA |
| **Number of Chronic comorbidities** |  |  |  |  |  |  |  |  | |  |
| Median [Q1, Q3] | 1.00 [0, 2.00] | 2.00 [1.00, 3.00] | 2.00 [1.00, 3.00] | 1.00 [1.00, 2.00] | 1.00 [0, 2.00] | 1.00 [0, 2.00] | 1.00 [0, 2.00] | <0.001 | | Kruskal-Wallis |
| **Health problems limit normal activities** |  |  |  |  |  |  |  |  | |  |
| Yes, severely | 255 (10.6%) | 43 (23.6%) | 76 (16.9%) | 32 (15.0%) | 26 (7.5%) | 46 (4.7%) | 32 (14.1%) | <0.001 | | Chi-Square |
| Yes, slightly | 435 (18.2%) | 37 (20.3%) | 87 (19.3%) | 39 (18.2%) | 62 (17.8%) | 165 (16.9%) | 45 (19.8%) |  | |  |
| No | 1699 (70.9%) | 101 (55.5%) | 286 (63.6%) | 141 (65.9%) | 260 (74.7%) | 762 (78.2%) | 149 (65.6%) |  | |  |
| **Self-perceived health** |  |  |  |  |  |  |  |  | |  |
| Excellent | 365 (15.2%) | 13 (7.1%) | 52 (11.6%) | 31 (14.5%) | 57 (16.4%) | 178 (18.3%) | 34 (15.0%) | <0.001 | | Chi-Square |
| Good | 1265 (52.8%) | 81 (44.5%) | 227 (50.4%) | 101 (47.2%) | 185 (53.2%) | 559 (57.4%) | 112 (49.3%) |  | |  |
| Fair | 502 (21.0%) | 51 (28.0%) | 107 (23.8%) | 59 (27.6%) | 69 (19.8%) | 170 (17.5%) | 46 (20.3%) |  | |  |
| Sometimes good/bad | 196 (8.2%) | 25 (13.7%) | 41 (9.1%) | 16 (7.5%) | 27 (7.8%) | 59 (6.1%) | 28 (12.3%) |  | |  |
| Poor | 61 (2.5%) | 11 (6.0%) | 22 (4.9%) | 5 (2.3%) | 9 (2.6%) | 7 (0.7%) | 7 (3.1%) |  | |  |

#### Table F4 – Strength trajectories in the sample of Women

|  |  | **Strength trajectory clusters** | | | | | |  | |
| --- | --- | --- | --- | --- | --- | --- | --- | --- | --- |
| Women | **Overall (N=2568)** | **Deceased Early (N=235)** | **Deceased Late (N=160)** | **Predominantly Low (N=488)** | **Predominantly Medium (N=482)** | **Predominantly High (N=766)** | **Increasing Med 🡪 High (N=437)** | **Adjusted p-value** | |
| **Recruitment cohort** |  |  |  |  |  |  |  |  |  |
| Cohort 1 (1992) | 1522 (59.3%) | 209 (88.9%) | 137 (85.6%) | 366 (75.0%) | 253 (52.5%) | 338 (44.1%) | 219 (50.1%) | <0.001 | Chi-Square |
| Cohort 2 (2002) | 521 (20.3%) | 15 (6.4%) | 18 (11.3%) | 70 (14.3%) | 108 (22.4%) | 195 (25.5%) | 115 (26.3%) |  |  |
| Cohort 3 (2012) | 525 (20.4%) | 11 (4.7%) | 5 (3.1%) | 52 (10.7%) | 121 (25.1%) | 233 (30.4%) | 103 (23.6%) |  |  |
| **Years participating in the study** |  |  |  |  |  |  |  |  |  |
| Mean (sd) | 11.6 (8.23) | 1.23 (1.55) | 4.68 (2.42) | 13.2 (6.73) | 13.4 (7.73) | 13.4 (8.10) | 12.7 (8.66) | <0.001 | ANOVA |
| **Age at baseline** |  |  |  |  |  |  |  |  |  |
| Mean (sd) | 66.1 (8.55) | 75.2 (8.61) | 74.8 (8.07) | 69.7 (8.80) | 64.3 (7.31) | 62.1 (5.41) | 63.1 (6.57) | <0.001 | ANOVA |
| **Partner status** |  |  |  |  |  |  |  |  |  |
| No partner | 940 (36.6%) | 149 (63.4%) | 97 (60.6%) | 213 (43.6%) | 156 (32.4%) | 198 (25.8%) | 127 (29.1%) | 0.0085 | Fisher's exact |
| Partner (co-residence) | 1559 (60.7%) | 85 (36.2%) | 60 (37.5%) | 260 (53.3%) | 312 (64.7%) | 545 (71.1%) | 297 (68.0%) |  |  |
| Partner (outside household) | 69 (2.7%) | 1 (0.4%) | 3 (1.9%) | 15 (3.1%) | 14 (2.9%) | 23 (3.0%) | 13 (3.0%) |  |  |
| **Educational level** |  |  |  |  |  |  |  |  |  |
| Lower Education | 1441 (56.1%) | 173 (73.6%) | 116 (72.5%) | 276 (56.6%) | 214 (44.4%) | 415 (54.2%) | 247 (56.5%) | <0.001 | Chi-Square |
| Intermediate Education | 796 (31.0%) | 43 (18.3%) | 33 (20.6%) | 145 (29.7%) | 193 (40.0%) | 256 (33.4%) | 126 (28.8%) |  |  |
| Higher Education | 328 (12.8%) | 19 (8.1%) | 11 (6.9%) | 66 (13.5%) | 74 (15.4%) | 94 (12.3%) | 64 (14.6%) |  |  |
| **Number of Sports** |  |  |  |  |  |  |  |  |  |
| 0 | 1015 (39.5%) | 135 (57.4%) | 81 (50.6%) | 216 (44.3%) | 155 (32.2%) | 293 (38.3%) | 135 (30.9%) | <0.001 | Chi-Square |
| 1 | 811 (31.6%) | 70 (29.8%) | 56 (35.0%) | 150 (30.7%) | 156 (32.4%) | 236 (30.8%) | 143 (32.7%) |  |  |
| 2+ | 742 (28.9%) | 30 (12.8%) | 23 (14.4%) | 122 (25.0%) | 171 (35.5%) | 237 (30.9%) | 159 (36.4%) |  |  |
| **Total PA duration: mean min/day** |  |  |  |  |  |  |  |  |  |
| Mean (sd) | 199 (111) | 160 (101) | 174 (109) | 163 (97.4) | 215 (113) | 225 (114) | 207 (108) | <0.001 | ANOVA |
| **PA component: Duration** |  |  |  |  |  |  |  |  |  |
| Long duration | 1165 (45.4%) | 73 (31.1%) | 58 (36.3%) | 173 (35.5%) | 232 (48.1%) | 418 (54.6%) | 211 (48.3%) | <0.001 | Chi-Square |
| Medium duration | 962 (37.5%) | 96 (40.9%) | 59 (36.9%) | 175 (35.9%) | 199 (41.3%) | 264 (34.5%) | 169 (38.7%) |  |  |
| Short duration | 422 (16.4%) | 63 (26.8%) | 40 (25.0%) | 136 (27.9%) | 50 (10.4%) | 76 (9.9%) | 57 (13.0%) |  |  |
| **PA component: Muscle Strength** |  |  |  |  |  |  |  |  |  |
| High Strength | 1025 (39.9%) | 23 (9.8%) | 59 (36.9%) | 76 (15.6%) | 159 (33.0%) | 699 (91.3%) | 9 (2.1%) | 0.0085 | Fisher's exact |
| Medium Strength | 915 (35.6%) | 85 (36.2%) | 26 (16.3%) | 108 (22.1%) | 322 (66.8%) | 20 (2.6%) | 354 (81.0%) |  |  |
| Low Strength | 609 (23.7%) | 124 (52.8%) | 72 (45.0%) | 300 (61.5%) | 0 (0%) | 39 (5.1%) | 74 (16.9%) |  |  |
| **PA component: Intensity** |  |  |  |  |  |  |  |  |  |
| High Intensity | 1102 (42.9%) | 36 (15.3%) | 29 (18.1%) | 105 (21.5%) | 245 (50.8%) | 462 (60.3%) | 225 (51.5%) | 0.0085 | Fisher's exact |
| Medium Intensity | 1053 (41.0%) | 100 (42.6%) | 64 (40.0%) | 201 (41.2%) | 235 (48.8%) | 275 (35.9%) | 178 (40.7%) |  |  |
| Low Intensity | 394 (15.3%) | 96 (40.9%) | 64 (40.0%) | 178 (36.5%) | 1 (0.2%) | 21 (2.7%) | 34 (7.8%) |  |  |
| **PA component: Mechanical Strain** |  |  |  |  |  |  |  |  |  |
| High Mech.Strain | 415 (16.2%) | 33 (14.0%) | 19 (11.9%) | 77 (15.8%) | 113 (23.4%) | 85 (11.1%) | 88 (20.1%) | <0.001 | Chi-Square |
| Medium Mech.Strain | 1681 (65.5%) | 157 (66.8%) | 108 (67.5%) | 297 (60.9%) | 322 (66.8%) | 497 (64.9%) | 300 (68.6%) |  |  |
| Low Mech.Strain | 453 (17.6%) | 42 (17.9%) | 30 (18.8%) | 110 (22.5%) | 46 (9.5%) | 176 (23.0%) | 49 (11.2%) |  |  |
| **PA component: Turning Actions** |  |  |  |  |  |  |  |  |  |
| High Turning | 558 (21.7%) | 67 (28.5%) | 43 (26.9%) | 133 (27.3%) | 124 (25.7%) | 107 (14.0%) | 84 (19.2%) | <0.001 | Chi-Square |
| Medium Turning | 1621 (63.1%) | 154 (65.5%) | 97 (60.6%) | 290 (59.4%) | 318 (66.0%) | 434 (56.7%) | 328 (75.1%) |  |  |
| Low Turning | 370 (14.4%) | 11 (4.7%) | 17 (10.6%) | 61 (12.5%) | 39 (8.1%) | 217 (28.3%) | 25 (5.7%) |  |  |
| **Functional limitation scale[0-9]** |  |  |  |  |  |  |  |  |  |
| Median [Q1, Q3] | 0 [0, 2.00] | 3.00 [0, 7.00] | 2.50 [0, 5.00] | 1.00 [0, 4.00] | 0 [0, 1.00] | 0 [0, 1.00] | 0 [0, 1.00] | <0.001 | Kruskal-Wallis |
| **MMSE-score based on maximum spel/num** |  |  |  |  |  |  |  |  |  |
| Mean (sd) | 27.4 (2.62) | 25.8 (3.60) | 26.1 (3.36) | 27.3 (2.62) | 27.8 (2.17) | 27.9 (2.09) | 27.7 (2.48) | <0.001 | ANOVA |
| **Number of Chronic comorbidities** |  |  |  |  |  |  |  |  |  |
| Median [Q1, Q3] | 2.00 [1.00, 3.00] | 2.00 [1.00, 3.00] | 2.00 [1.00, 3.00] | 2.00 [1.00, 3.00] | 1.00 [1.00, 2.00] | 1.00 [0, 2.00] | 1.00 [1.00, 2.00] | <0.001 | Kruskal-Wallis |
| **Health problems limit normal activities** |  |  |  |  |  |  |  |  |  |
| Yes, severely | 275 (10.7%) | 48 (20.4%) | 25 (15.6%) | 97 (19.9%) | 17 (3.5%) | 51 (6.7%) | 37 (8.5%) | <0.001 | Chi-Square |
| Yes, slightly | 581 (22.6%) | 73 (31.1%) | 45 (28.1%) | 139 (28.5%) | 97 (20.1%) | 129 (16.8%) | 98 (22.4%) |  |  |
| No | 1707 (66.5%) | 113 (48.1%) | 90 (56.3%) | 251 (51.4%) | 367 (76.1%) | 586 (76.5%) | 300 (68.6%) |  |  |
| **Self-perceived health** |  |  |  |  |  |  |  |  |  |
| Excellent | 347 (13.5%) | 10 (4.3%) | 24 (15.0%) | 50 (10.2%) | 75 (15.6%) | 117 (15.3%) | 71 (16.2%) | 0.0085 | Fisher's exact |
| Good | 1247 (48.6%) | 98 (41.7%) | 63 (39.4%) | 181 (37.1%) | 248 (51.5%) | 445 (58.1%) | 212 (48.5%) |  |  |
| Fair | 601 (23.4%) | 83 (35.3%) | 39 (24.4%) | 150 (30.7%) | 107 (22.2%) | 121 (15.8%) | 101 (23.1%) |  |  |
| Sometimes good/bad | 309 (12.0%) | 30 (12.8%) | 27 (16.9%) | 85 (17.4%) | 49 (10.2%) | 76 (9.9%) | 42 (9.6%) |  |  |
| Poor | 56 (2.2%) | 14 (6.0%) | 5 (3.1%) | 20 (4.1%) | 2 (0.4%) | 5 (0.7%) | 10 (2.3%) |  |  |

#### Table F5 – Mechanical Strain trajectories in the sample of Men

|  |  | **Mechanical Strain** **trajectory clusters** | | | | | |  | |
| --- | --- | --- | --- | --- | --- | --- | --- | --- | --- |
| Men | **Overall (N=2395)** | **Deceased Early (N=182)** | **Deceased Late (N=469)** | **Predominantly Medium (N=578)** | **Predominantly High (N=556)** | **Decreasing Med 🡪 Low (N=332)** | **Increasing Low 🡪 Med (N=278)** | **Adjusted  p-value** | |
| **Recruitment cohort** |  |  |  |  |  |  |  |  |  |
| Cohort 1 (1992) | 1432 (59.8%) | 166 (91.2%) | 414 (88.3%) | 294 (50.9%) | 234 (42.1%) | 185 (55.7%) | 139 (50.0%) | 0.0085 | Fisher's exact |
| Cohort 2 (2002) | 470 (19.6%) | 12 (6.6%) | 29 (6.2%) | 133 (23.0%) | 146 (26.3%) | 71 (21.4%) | 79 (28.4%) |  |  |
| Cohort 3 (2012) | 493 (20.6%) | 4 (2.2%) | 26 (5.5%) | 151 (26.1%) | 176 (31.7%) | 76 (22.9%) | 60 (21.6%) |  |  |
| **Years participating in the study** |  |  |  |  |  |  |  |  |  |
| Mean (sd) | 9.95 (7.84) | 0 (0) | 3.53 (2.15) | 14.4 (6.76) | 11.4 (7.51) | 11.2 (7.41) | 13.5 (7.32) | <0.001 | ANOVA |
| **Age at baseline** |  |  |  |  |  |  |  |  |  |
| Mean (sd) | 66.4 (8.68) | 74.9 (8.43) | 73.8 (8.27) | 63.3 (6.66) | 63.1 (6.75) | 64.5 (7.33) | 63.8 (6.93) | <0.001 | ANOVA |
| **Partner status** |  |  |  |  |  |  |  |  |  |
| No partner | 375 (15.7%) | 51 (28.0%) | 111 (23.7%) | 65 (11.2%) | 69 (12.4%) | 45 (13.6%) | 34 (12.2%) | 0.0085 | Fisher's exact |
| Partner (co-residence) | 1924 (80.3%) | 127 (69.8%) | 341 (72.7%) | 495 (85.6%) | 458 (82.4%) | 272 (81.9%) | 231 (83.1%) |  |  |
| Partner (outside household) | 95 (4.0%) | 4 (2.2%) | 17 (3.6%) | 18 (3.1%) | 28 (5.0%) | 15 (4.5%) | 13 (4.7%) |  |  |
| **Educational level** |  |  |  |  |  |  |  |  |  |
| Lower Education | 1110 (46.3%) | 118 (64.8%) | 268 (57.1%) | 237 (41.0%) | 199 (35.8%) | 151 (45.5%) | 137 (49.3%) | <0.001 | Chi-Square |
| Intermediate Education | 732 (30.6%) | 39 (21.4%) | 121 (25.8%) | 183 (31.7%) | 206 (37.1%) | 109 (32.8%) | 74 (26.6%) |  |  |
| Higher Education | 553 (23.1%) | 25 (13.7%) | 80 (17.1%) | 158 (27.3%) | 151 (27.2%) | 72 (21.7%) | 67 (24.1%) |  |  |
| **Number of Sports** |  |  |  |  |  |  |  |  |  |
| 0 | 1028 (42.9%) | 113 (62.1%) | 265 (56.5%) | 289 (50.0%) | 161 (29.0%) | 112 (33.7%) | 88 (31.7%) | <0.001 | Chi-Square |
| 1 | 700 (29.2%) | 44 (24.2%) | 126 (26.9%) | 119 (20.6%) | 189 (34.0%) | 114 (34.3%) | 108 (38.8%) |  |  |
| 2+ | 667 (27.8%) | 25 (13.7%) | 78 (16.6%) | 170 (29.4%) | 206 (37.1%) | 106 (31.9%) | 82 (29.5%) |  |  |
| **Total PA duration: mean min/day** |  |  |  |  |  |  |  |  |  |
| Mean (sd) | 134 (102) | 118 (92.4) | 122 (97.6) | 145 (107) | 138 (101) | 135 (101) | 135 (103) | 0.0356 | ANOVA |
| **PA component: Duration** |  |  |  |  |  |  |  |  |  |
| Long duration | 512 (21.4%) | 27 (14.8%) | 81 (17.3%) | 147 (25.4%) | 129 (23.2%) | 73 (22.0%) | 55 (19.8%) | 0.0741 | Chi-Square |
| Medium duration | 830 (34.7%) | 77 (42.3%) | 148 (31.6%) | 194 (33.6%) | 197 (35.4%) | 116 (34.9%) | 98 (35.3%) |  |  |
| Short duration | 1039 (43.4%) | 78 (42.9%) | 237 (50.5%) | 232 (40.1%) | 227 (40.8%) | 142 (42.8%) | 123 (44.2%) |  |  |
| **PA component: Muscle Strength** |  |  |  |  |  |  |  |  |  |
| High Strength | 1123 (46.9%) | 55 (30.2%) | 156 (33.3%) | 322 (55.7%) | 275 (49.5%) | 162 (48.8%) | 153 (55.0%) | <0.001 | Chi-Square |
| Medium Strength | 676 (28.2%) | 56 (30.8%) | 132 (28.1%) | 174 (30.1%) | 182 (32.7%) | 91 (27.4%) | 41 (14.7%) |  |  |
| Low Strength | 582 (24.3%) | 71 (39.0%) | 178 (38.0%) | 77 (13.3%) | 96 (17.3%) | 78 (23.5%) | 82 (29.5%) |  |  |
| **PA component: Intensity** |  |  |  |  |  |  |  |  |  |
| High Intensity | 1036 (43.3%) | 43 (23.6%) | 123 (26.2%) | 246 (42.6%) | 304 (54.7%) | 160 (48.2%) | 160 (57.6%) | <0.001 | Chi-Square |
| Medium Intensity | 1020 (42.6%) | 94 (51.6%) | 236 (50.3%) | 264 (45.7%) | 199 (35.8%) | 133 (40.1%) | 94 (33.8%) |  |  |
| Low Intensity | 325 (13.6%) | 45 (24.7%) | 107 (22.8%) | 63 (10.9%) | 50 (9.0%) | 38 (11.4%) | 22 (7.9%) |  |  |
| **PA component: Mechanical Strain** |  |  |  |  |  |  |  |  |  |
| High Mech.Strain | 474 (19.8%) | 32 (17.6%) | 82 (17.5%) | 1 (0.2%) | 271 (48.7%) | 88 (26.5%) | 0 (0%) | 0.0085 | Fisher's exact |
| Medium Mech.Strain | 1393 (58.2%) | 108 (59.3%) | 260 (55.4%) | 572 (99.0%) | 268 (48.2%) | 151 (45.5%) | 34 (12.2%) |  |  |
| Low Mech.Strain | 514 (21.5%) | 42 (23.1%) | 124 (26.4%) | 0 (0%) | 14 (2.5%) | 92 (27.7%) | 242 (87.1%) |  |  |
| **PA component: Turning Actions** |  |  |  |  |  |  |  |  |  |
| High Turning | 355 (14.8%) | 23 (12.6%) | 73 (15.6%) | 31 (5.4%) | 158 (28.4%) | 63 (19.0%) | 7 (2.5%) | <0.001 | Chi-Square |
| Medium Turning | 1309 (54.7%) | 108 (59.3%) | 267 (56.9%) | 402 (69.6%) | 280 (50.4%) | 156 (47.0%) | 96 (34.5%) |  |  |
| Low Turning | 717 (29.9%) | 51 (28.0%) | 126 (26.9%) | 140 (24.2%) | 115 (20.7%) | 112 (33.7%) | 173 (62.2%) |  |  |
| **Functional limitation scale[0-9]** |  |  |  |  |  |  |  |  |  |
| Median [Q1, Q3] | 0 [0, 1.00] | 1.00 [0, 3.00] | 0 [0, 2.00] | 0 [0, 0] | 0 [0, 0] | 0 [0, 1.00] | 0 [0, 1.00] | <0.001 | Kruskal-Wallis |
| **MMSE-score based on maximum spel/num** |  |  |  |  |  |  |  |  |  |
| Mean (sd) | 27.4 (2.53) | 25.8 (4.08) | 26.5 (2.94) | 28.0 (1.91) | 27.9 (2.00) | 27.7 (1.96) | 27.7 (2.27) | <0.001 | ANOVA |
| **Number of Chronic comorbidities** |  |  |  |  |  |  |  |  |  |
| Median [Q1, Q3] | 1.00 [0, 2.00] | 2.00 [1.00, 3.00] | 1.00 [1.00, 3.00] | 1.00 [0, 2.00] | 1.00 [0, 2.00] | 1.00 [1.00, 2.00] | 1.00 [0, 2.00] | <0.001 | Kruskal-Wallis |
| **Health problems limit normal activities** |  |  |  |  |  |  |  |  |  |
| Yes, severely | 255 (10.6%) | 43 (23.6%) | 76 (16.2%) | 33 (5.7%) | 39 (7.0%) | 45 (13.6%) | 19 (6.8%) | <0.001 | Chi-Square |
| Yes, slightly | 435 (18.2%) | 37 (20.3%) | 89 (19.0%) | 114 (19.7%) | 85 (15.3%) | 56 (16.9%) | 54 (19.4%) |  |  |
| No | 1699 (70.9%) | 101 (55.5%) | 302 (64.4%) | 429 (74.2%) | 431 (77.5%) | 231 (69.6%) | 205 (73.7%) |  |  |
| **Self-perceived health** |  |  |  |  |  |  |  |  |  |
| Excellent | 365 (15.2%) | 13 (7.1%) | 56 (11.9%) | 91 (15.7%) | 111 (20.0%) | 42 (12.7%) | 52 (18.7%) | 0.0085 | Fisher's exact |
| Good | 1265 (52.8%) | 81 (44.5%) | 238 (50.7%) | 327 (56.6%) | 305 (54.9%) | 166 (50.0%) | 148 (53.2%) |  |  |
| Fair | 502 (21.0%) | 51 (28.0%) | 111 (23.7%) | 118 (20.4%) | 88 (15.8%) | 77 (23.2%) | 57 (20.5%) |  |  |
| Sometimes good/bad | 196 (8.2%) | 25 (13.7%) | 40 (8.5%) | 36 (6.2%) | 40 (7.2%) | 37 (11.1%) | 18 (6.5%) |  |  |
| Poor | 61 (2.5%) | 11 (6.0%) | 22 (4.7%) | 6 (1.0%) | 9 (1.6%) | 10 (3.0%) | 3 (1.1%) |  |  |

#### Table F6 – Mechanical Strain trajectories in the sample of Women

|  |  | **Mechanical Strain** **trajectory clusters** | | | | | |  | |
| --- | --- | --- | --- | --- | --- | --- | --- | --- | --- |
| Women | **Overall (N=2568)** | **Deceased Early (N=281)** | **Deceased Late (N=148)** | **Predominantly Medium (N=901)** | **Predominantly High (N=258)** | **Increasing Med-High (N=467)** | **Other (N=513)** | **Adjusted  p-value** | |
| **Recruitment cohort** |  |  |  |  |  |  |  |  |  |
| Cohort 1 (1992) | 1522 (59.3%) | 245 (87.2%) | 130 (87.8%) | 528 (58.6%) | 112 (43.4%) | 226 (48.4%) | 281 (54.8%) | <0.001 | Chi-Square |
| Cohort 2 (2002) | 521 (20.3%) | 24 (8.5%) | 12 (8.1%) | 174 (19.3%) | 75 (29.1%) | 98 (21.0%) | 138 (26.9%) |  |  |
| Cohort 3 (2012) | 525 (20.4%) | 12 (4.3%) | 6 (4.1%) | 199 (22.1%) | 71 (27.5%) | 143 (30.6%) | 94 (18.3%) |  |  |
| **Years participating in the study** |  |  |  |  |  |  |  |  |  |
| Mean (sd) | 11.6 (8.23) | 1.66 (1.91) | 5.19 (2.70) | 14.9 (7.15) | 11.6 (8.56) | 9.75 (7.78) | 14.7 (7.47) | <0.001 | ANOVA |
| **Age at baseline** |  |  |  |  |  |  |  |  |  |
| Mean (sd) | 66.1 (8.55) | 74.8 (8.63) | 74.6 (8.23) | 64.8 (7.73) | 63.9 (6.90) | 64.3 (7.56) | 64.1 (7.13) | <0.001 | ANOVA |
| **Partner status** |  |  |  |  |  |  |  |  |  |
| No partner | 940 (36.6%) | 173 (61.6%) | 87 (58.8%) | 309 (34.3%) | 78 (30.2%) | 137 (29.3%) | 156 (30.4%) | 0.0085 | Fisher's exact |
| Partner (co-residence) | 1559 (60.7%) | 107 (38.1%) | 57 (38.5%) | 566 (62.8%) | 174 (67.4%) | 319 (68.3%) | 336 (65.5%) |  |  |
| Partner (outside household) | 69 (2.7%) | 1 (0.4%) | 4 (2.7%) | 26 (2.9%) | 6 (2.3%) | 11 (2.4%) | 21 (4.1%) |  |  |
| **Educational level** |  |  |  |  |  |  |  |  |  |
| Lower Education | 1441 (56.1%) | 211 (75.1%) | 104 (70.3%) | 491 (54.5%) | 116 (45.0%) | 229 (49.0%) | 290 (56.5%) | <0.001 | Chi-Square |
| Intermediate Education | 796 (31.0%) | 50 (17.8%) | 33 (22.3%) | 293 (32.5%) | 100 (38.8%) | 159 (34.0%) | 161 (31.4%) |  |  |
| Higher Education | 328 (12.8%) | 20 (7.1%) | 11 (7.4%) | 116 (12.9%) | 41 (15.9%) | 78 (16.7%) | 62 (12.1%) |  |  |
| **Number of Sports** |  |  |  |  |  |  |  |  |  |
| 0 | 1015 (39.5%) | 162 (57.7%) | 69 (46.6%) | 340 (37.7%) | 47 (18.2%) | 186 (39.8%) | 211 (41.1%) | <0.001 | Chi-Square |
| 1 | 811 (31.6%) | 81 (28.8%) | 57 (38.5%) | 294 (32.6%) | 87 (33.7%) | 134 (28.7%) | 158 (30.8%) |  |  |
| 2+ | 742 (28.9%) | 38 (13.5%) | 22 (14.9%) | 267 (29.6%) | 124 (48.1%) | 147 (31.5%) | 144 (28.1%) |  |  |
| **Total PA duration: mean min/day** |  |  |  |  |  |  |  |  |  |
| Mean (sd) | 199 (111) | 164 (102) | 183 (117) | 211 (109) | 210 (119) | 201 (115) | 195 (106) | <0.001 | ANOVA |
| **PA component: Duration** |  |  |  |  |  |  |  |  |  |
| Long duration | 1165 (45.4%) | 91 (32.4%) | 54 (36.5%) | 460 (51.1%) | 125 (48.4%) | 211 (45.2%) | 224 (43.7%) | <0.001 | Chi-Square |
| Medium duration | 962 (37.5%) | 113 (40.2%) | 58 (39.2%) | 322 (35.7%) | 98 (38.0%) | 182 (39.0%) | 189 (36.8%) |  |  |
| Short duration | 422 (16.4%) | 73 (26.0%) | 32 (21.6%) | 119 (13.2%) | 34 (13.2%) | 74 (15.8%) | 90 (17.5%) |  |  |
| **PA component: Muscle Strength** |  |  |  |  |  |  |  |  |  |
| High Strength | 1025 (39.9%) | 51 (18.1%) | 30 (20.3%) | 388 (43.1%) | 82 (31.8%) | 199 (42.6%) | 275 (53.6%) | <0.001 | Chi-Square |
| Medium Strength | 915 (35.6%) | 92 (32.7%) | 50 (33.8%) | 375 (41.6%) | 126 (48.8%) | 180 (38.5%) | 92 (17.9%) |  |  |
| Low Strength | 609 (23.7%) | 134 (47.7%) | 64 (43.2%) | 138 (15.3%) | 49 (19.0%) | 88 (18.8%) | 136 (26.5%) |  |  |
| **PA component: Intensity** |  |  |  |  |  |  |  |  |  |
| High Intensity | 1102 (42.9%) | 53 (18.9%) | 22 (14.9%) | 384 (42.6%) | 134 (51.9%) | 221 (47.3%) | 288 (56.1%) | <0.001 | Chi-Square |
| Medium Intensity | 1053 (41.0%) | 119 (42.3%) | 67 (45.3%) | 423 (46.9%) | 108 (41.9%) | 189 (40.5%) | 147 (28.7%) |  |  |
| Low Intensity | 394 (15.3%) | 105 (37.4%) | 55 (37.2%) | 94 (10.4%) | 15 (5.8%) | 57 (12.2%) | 68 (13.3%) |  |  |
| **PA component: Mechanical Strain** |  |  |  |  |  |  |  |  |  |
| High Mech.Strain | 415 (16.2%) | 34 (12.1%) | 29 (19.6%) | 155 (17.2%) | 152 (58.9%) | 38 (8.1%) | 7 (1.4%) | 0.0085 | Fisher's exact |
| Medium Mech.Strain | 1681 (65.5%) | 180 (64.1%) | 103 (69.6%) | 745 (82.7%) | 79 (30.6%) | 359 (76.9%) | 215 (41.9%) |  |  |
| Low Mech.Strain | 453 (17.6%) | 63 (22.4%) | 12 (8.1%) | 1 (0.1%) | 26 (10.1%) | 70 (15.0%) | 281 (54.8%) |  |  |
| **PA component: Turning Actions** |  |  |  |  |  |  |  |  |  |
| High Turning | 558 (21.7%) | 79 (28.1%) | 41 (27.7%) | 166 (18.4%) | 122 (47.3%) | 74 (15.8%) | 76 (14.8%) | <0.001 | Chi-Square |
| Medium Turning | 1621 (63.1%) | 182 (64.8%) | 92 (62.2%) | 610 (67.7%) | 116 (45.0%) | 307 (65.7%) | 314 (61.2%) |  |  |
| Low Turning | 370 (14.4%) | 16 (5.7%) | 11 (7.4%) | 125 (13.9%) | 19 (7.4%) | 86 (18.4%) | 113 (22.0%) |  |  |
| **Functional limitation scale[0-9]** |  |  |  |  |  |  |  |  |  |
| Median [Q1, Q3] | 0 [0, 2.00] | 3.00 [0, 6.00] | 2.00 [0, 5.00] | 0 [0, 1.00] | 0 [0, 1.00] | 0 [0, 1.00] | 0 [0, 2.00] | <0.001 | Kruskal-Wallis |
| **MMSE-score based on maximum spel/num** |  |  |  |  |  |  |  |  |  |
| Mean (sd) | 27.4 (2.62) | 25.7 (3.69) | 26.2 (3.36) | 28.0 (1.93) | 27.2 (3.04) | 27.5 (2.40) | 27.7 (2.15) | <0.001 | ANOVA |
| **Number of Chronic comorbidities** |  |  |  |  |  |  |  |  |  |
| Median [Q1, Q3] | 2.00 [1.00, 3.00] | 2.00 [1.00, 3.00] | 2.00 [1.00, 3.00] | 1.00 [1.00, 2.00] | 1.00 [1.00, 2.00] | 1.00 [1.00, 3.00] | 2.00 [1.00, 3.00] | <0.001 | Kruskal-Wallis |
| **Health problems limit normal activities** |  |  |  |  |  |  |  |  |  |
| Yes, severely | 275 (10.7%) | 56 (19.9%) | 20 (13.5%) | 60 (6.7%) | 23 (8.9%) | 58 (12.4%) | 58 (11.3%) | <0.001 | Chi-Square |
| Yes, slightly | 581 (22.6%) | 85 (30.2%) | 38 (25.7%) | 159 (17.6%) | 55 (21.3%) | 101 (21.6%) | 143 (27.9%) |  |  |
| No | 1707 (66.5%) | 139 (49.5%) | 90 (60.8%) | 679 (75.4%) | 180 (69.8%) | 307 (65.7%) | 312 (60.8%) |  |  |
| **Self-perceived health** |  |  |  |  |  |  |  |  |  |
| Excellent | 347 (13.5%) | 15 (5.3%) | 24 (16.2%) | 139 (15.4%) | 36 (14.0%) | 68 (14.6%) | 65 (12.7%) | 0.0085 | Fisher's exact |
| Good | 1247 (48.6%) | 118 (42.0%) | 60 (40.5%) | 486 (53.9%) | 130 (50.4%) | 222 (47.5%) | 231 (45.0%) |  |  |
| Fair | 601 (23.4%) | 95 (33.8%) | 33 (22.3%) | 183 (20.3%) | 53 (20.5%) | 113 (24.2%) | 124 (24.2%) |  |  |
| Sometimes good/bad | 309 (12.0%) | 37 (13.2%) | 25 (16.9%) | 77 (8.5%) | 36 (14.0%) | 55 (11.8%) | 79 (15.4%) |  |  |
| Poor | 56 (2.2%) | 15 (5.3%) | 5 (3.4%) | 13 (1.4%) | 2 (0.8%) | 8 (1.7%) | 13 (2.5%) |  |  |

#### Table F7 – Turning Actions trajectories in the sample of Men

|  |  | **Turning Actions trajectory clusters** | | | | | |  | | |
| --- | --- | --- | --- | --- | --- | --- | --- | --- | --- | --- |
| Men | **Overall (N=2395)** | **Deceased Early (N=364)** | **Deceased Late (N=293)** | **Predominantly Low (N=587)** | **Predominantly Medium (N=591)** | **Decreasing High 🡪 Med (N=346)** | **Increasing Med 🡪 High (N=214)** | **Adjusted p-value** |  | |
| **Recruitment cohort** |  |  |  |  |  |  |  |  | |  |
| Cohort 1 (1992) | 1432 (59.8%) | 329 (90.4%) | 256 (87.4%) | 283 (48.2%) | 322 (54.5%) | 141 (40.8%) | 101 (47.2%) | <0.001 | | Chi-Square |
| Cohort 2 (2002) | 470 (19.6%) | 20 (5.5%) | 22 (7.5%) | 147 (25.0%) | 119 (20.1%) | 109 (31.5%) | 53 (24.8%) |  | |  |
| Cohort 3 (2012) | 493 (20.6%) | 15 (4.1%) | 15 (5.1%) | 157 (26.7%) | 150 (25.4%) | 96 (27.7%) | 60 (28.0%) |  | |  |
| **Years participating in the study** |  |  |  |  |  |  |  |  | |  |
| Mean (sd) | 9.95 (7.84) | 1.19 (1.60) | 4.45 (2.98) | 14.0 (7.16) | 13.3 (6.68) | 12.4 (7.70) | 7.80 (6.91) | <0.001 | | ANOVA |
| **Age at baseline** |  |  |  |  |  |  |  |  | |  |
| Mean (sd) | 66.4 (8.68) | 74.8 (8.34) | 73.0 (8.29) | 63.2 (6.25) | 64.3 (7.30) | 62.4 (6.58) | 64.3 (7.43) | <0.001 | | ANOVA |
| **Partner status** |  |  |  |  |  |  |  |  | |  |
| No partner | 375 (15.7%) | 102 (28.0%) | 60 (20.5%) | 56 (9.5%) | 78 (13.2%) | 48 (13.9%) | 31 (14.5%) | <0.001 | | Chi-Square |
| Partner (co-residence) | 1924 (80.3%) | 252 (69.2%) | 222 (75.8%) | 510 (86.9%) | 485 (82.1%) | 282 (81.5%) | 173 (80.8%) |  | |  |
| Partner (outside household) | 95 (4.0%) | 10 (2.7%) | 11 (3.8%) | 21 (3.6%) | 28 (4.7%) | 15 (4.3%) | 10 (4.7%) |  | |  |
| **Educational level** |  |  |  |  |  |  |  |  | |  |
| Lower Education | 1110 (46.3%) | 229 (62.9%) | 161 (54.9%) | 254 (43.3%) | 247 (41.8%) | 119 (34.4%) | 100 (46.7%) | <0.001 | | Chi-Square |
| Intermediate Education | 732 (30.6%) | 80 (22.0%) | 81 (27.6%) | 186 (31.7%) | 198 (33.5%) | 129 (37.3%) | 58 (27.1%) |  | |  |
| Higher Education | 553 (23.1%) | 55 (15.1%) | 51 (17.4%) | 147 (25.0%) | 146 (24.7%) | 98 (28.3%) | 56 (26.2%) |  | |  |
| **Number of Sports** |  |  |  |  |  |  |  |  | |  |
| 0 | 1028 (42.9%) | 220 (60.4%) | 163 (55.6%) | 199 (33.9%) | 266 (45.0%) | 82 (23.7%) | 98 (45.8%) | <0.001 | | Chi-Square |
| 1 | 700 (29.2%) | 98 (26.9%) | 72 (24.6%) | 185 (31.5%) | 151 (25.6%) | 125 (36.1%) | 69 (32.2%) |  | |  |
| 2+ | 667 (27.8%) | 46 (12.6%) | 58 (19.8%) | 203 (34.6%) | 174 (29.4%) | 139 (40.2%) | 47 (22.0%) |  | |  |
| **Total PA duration: mean min/day** |  |  |  |  |  |  |  |  | |  |
| Mean (sd) | 134 (102) | 119 (97.0) | 123 (95.2) | 139 (104) | 144 (103) | 138 (106) | 131 (99.5) | 0.042 | | ANOVA |
| **PA component: Duration** |  |  |  |  |  |  |  |  | |  |
| Long duration | 512 (21.4%) | 60 (16.5%) | 49 (16.7%) | 133 (22.7%) | 147 (24.9%) | 74 (21.4%) | 49 (22.9%) | 0.433 | | Chi-Square |
| Medium duration | 830 (34.7%) | 131 (36.0%) | 95 (32.4%) | 207 (35.3%) | 211 (35.7%) | 120 (34.7%) | 66 (30.8%) |  | |  |
| Short duration | 1039 (43.4%) | 171 (47.0%) | 146 (49.8%) | 247 (42.1%) | 228 (38.6%) | 149 (43.1%) | 98 (45.8%) |  | |  |
| **PA component: Muscle Strength** |  |  |  |  |  |  |  |  | |  |
| High Strength | 1123 (46.9%) | 117 (32.1%) | 95 (32.4%) | 335 (57.1%) | 300 (50.8%) | 183 (52.9%) | 93 (43.5%) | <0.001 | | Chi-Square |
| Medium Strength | 676 (28.2%) | 110 (30.2%) | 79 (27.0%) | 144 (24.5%) | 170 (28.8%) | 113 (32.7%) | 60 (28.0%) |  | |  |
| Low Strength | 582 (24.3%) | 135 (37.1%) | 116 (39.6%) | 108 (18.4%) | 116 (19.6%) | 47 (13.6%) | 60 (28.0%) |  | |  |
| **PA component: Intensity** |  |  |  |  |  |  |  |  | |  |
| High Intensity | 1036 (43.3%) | 77 (21.2%) | 89 (30.4%) | 322 (54.9%) | 239 (40.4%) | 212 (61.3%) | 97 (45.3%) | <0.001 | | Chi-Square |
| Medium Intensity | 1020 (42.6%) | 195 (53.6%) | 138 (47.1%) | 212 (36.1%) | 287 (48.6%) | 111 (32.1%) | 77 (36.0%) |  | |  |
| Low Intensity | 325 (13.6%) | 90 (24.7%) | 63 (21.5%) | 53 (9.0%) | 60 (10.2%) | 20 (5.8%) | 39 (18.2%) |  | |  |
| **PA component: Mechanical Strain** |  |  |  |  |  |  |  |  | |  |
| High Mech.Strain | 474 (19.8%) | 74 (20.3%) | 40 (13.7%) | 86 (14.7%) | 65 (11.0%) | 184 (53.2%) | 25 (11.7%) | <0.001 | | Chi-Square |
| Medium Mech.Strain | 1393 (58.2%) | 219 (60.2%) | 153 (52.2%) | 351 (59.8%) | 405 (68.5%) | 136 (39.3%) | 129 (60.3%) |  | |  |
| Low Mech.Strain | 514 (21.5%) | 69 (19.0%) | 97 (33.1%) | 150 (25.6%) | 116 (19.6%) | 23 (6.6%) | 59 (27.6%) |  | |  |
| **PA component: Turning Actions** |  |  |  |  |  |  |  |  | |  |
| High Turning | 355 (14.8%) | 63 (17.3%) | 33 (11.3%) | 22 (3.7%) | 0 (0%) | 232 (67.1%) | 5 (2.3%) | 0.0085 | | Fisher's exact |
| Medium Turning | 1309 (54.7%) | 248 (68.1%) | 131 (44.7%) | 260 (44.3%) | 457 (77.3%) | 77 (22.3%) | 136 (63.6%) |  | |  |
| Low Turning | 717 (29.9%) | 51 (14.0%) | 126 (43.0%) | 305 (52.0%) | 129 (21.8%) | 34 (9.8%) | 72 (33.6%) |  | |  |
| **Functional limitation scale[0-9]** |  |  |  |  |  |  |  |  | |  |
| Median [Q1, Q3] | 0 [0, 1.00] | 1.00 [0, 3.00] | 0 [0, 2.00] | 0 [0, 0] | 0 [0, 0] | 0 [0, 0] | 0 [0, 1.00] | <0.001 | | Kruskal-Wallis |
| **MMSE-score based on maximum spel/num** |  |  |  |  |  |  |  |  | |  |
| Mean (sd) | 27.4 (2.53) | 26.1 (3.62) | 26.6 (2.84) | 27.9 (1.88) | 27.9 (2.03) | 28.0 (2.01) | 27.4 (2.26) | <0.001 | | ANOVA |
| **Number of Chronic comorbidities** |  |  |  |  |  |  |  |  | |  |
| Median [Q1, Q3] | 1.00 [0, 2.00] | 2.00 [1.00, 3.00] | 1.00 [0, 2.00] | 1.00 [0, 2.00] | 1.00 [0, 2.00] | 1.00 [0, 2.00] | 1.00 [0, 2.00] | <0.001 | | Kruskal-Wallis |
| **Health problems limit normal activities** |  |  |  |  |  |  |  |  | |  |
| Yes, severely | 255 (10.6%) | 80 (22.0%) | 39 (13.3%) | 48 (8.2%) | 33 (5.6%) | 24 (6.9%) | 31 (14.5%) | <0.001 | | Chi-Square |
| Yes, slightly | 435 (18.2%) | 73 (20.1%) | 54 (18.4%) | 101 (17.2%) | 112 (19.0%) | 57 (16.5%) | 38 (17.8%) |  | |  |
| No | 1699 (70.9%) | 210 (57.7%) | 198 (67.6%) | 436 (74.3%) | 446 (75.5%) | 264 (76.3%) | 145 (67.8%) |  | |  |
| **Self-perceived health** |  |  |  |  |  |  |  |  | |  |
| Excellent | 365 (15.2%) | 33 (9.1%) | 39 (13.3%) | 94 (16.0%) | 100 (16.9%) | 57 (16.5%) | 42 (19.6%) | 0.0085 | | Fisher's exact |
| Good | 1265 (52.8%) | 174 (47.8%) | 146 (49.8%) | 332 (56.6%) | 323 (54.7%) | 190 (54.9%) | 100 (46.7%) |  | |  |
| Fair | 502 (21.0%) | 97 (26.6%) | 66 (22.5%) | 117 (19.9%) | 119 (20.1%) | 66 (19.1%) | 37 (17.3%) |  | |  |
| Sometimes good/bad | 196 (8.2%) | 40 (11.0%) | 26 (8.9%) | 40 (6.8%) | 42 (7.1%) | 26 (7.5%) | 22 (10.3%) |  | |  |
| Poor | 61 (2.5%) | 19 (5.2%) | 14 (4.8%) | 4 (0.7%) | 6 (1.0%) | 5 (1.4%) | 13 (6.1%) |  | |  |

#### Table F8 – Turning Actions trajectories in the sample of Women

|  |  | **Turning Actions** **trajectory clusters** | | | | | |  | | |
| --- | --- | --- | --- | --- | --- | --- | --- | --- | --- | --- |
| Women | **Overall (N=2568)** | **Deceased Early (N=258)** | **Predominantly Medium (N=509)** | **Decreasing High-Med (N=444)** | **Decreasing Med-Low (N=588)** | **Increasing Low-Med (N=361)** | **Increasing Med-High (N=408)** | **Adjusted p-value** |  | |
| Cohort 1 (1992) | 1522 (59.3%) | 224 (86.8%) | 288 (56.6%) | 278 (62.6%) | 303 (51.5%) | 142 (39.3%) | 287 (70.3%) | <0.001 | | Chi-Square |
| Cohort 2 (2002) | 521 (20.3%) | 22 (8.5%) | 112 (22.0%) | 102 (23.0%) | 134 (22.8%) | 89 (24.7%) | 62 (15.2%) |  | |  |
| Cohort 3 (2012) | 525 (20.4%) | 12 (4.7%) | 109 (21.4%) | 64 (14.4%) | 151 (25.7%) | 130 (36.0%) | 59 (14.5%) |  | |  |
| **Years participating in the study** |  |  |  |  |  |  |  |  | |  |
| Mean (sd) | 11.6 (8.23) | 1.35 (1.58) | 15.8 (7.00) | 13.8 (7.93) | 12.0 (7.85) | 12.6 (7.67) | 8.93 (7.31) | <0.001 | | ANOVA |
| **Age at baseline** |  |  |  |  |  |  |  |  | |  |
| Mean (sd) | 66.1 (8.55) | 74.7 (8.69) | 63.6 (6.81) | 66.6 (8.52) | 64.7 (7.62) | 62.6 (6.19) | 68.5 (9.22) | <0.001 | | ANOVA |
| **Partner status** |  |  |  |  |  |  |  |  | |  |
| No partner | 940 (36.6%) | 158 (61.2%) | 162 (31.8%) | 166 (37.4%) | 198 (33.7%) | 84 (23.3%) | 172 (42.2%) | 0.0085 | | Fisher's exact |
| Partner (co-residence) | 1559 (60.7%) | 99 (38.4%) | 331 (65.0%) | 265 (59.7%) | 376 (63.9%) | 261 (72.3%) | 227 (55.6%) |  | |  |
| Partner (outside household) | 69 (2.7%) | 1 (0.4%) | 16 (3.1%) | 13 (2.9%) | 14 (2.4%) | 16 (4.4%) | 9 (2.2%) |  | |  |
| **Educational level** |  |  |  |  |  |  |  |  | |  |
| Lower Education | 1441 (56.1%) | 191 (74.0%) | 270 (53.0%) | 237 (53.4%) | 326 (55.4%) | 165 (45.7%) | 252 (61.8%) | <0.001 | | Chi-Square |
| Intermediate Education | 796 (31.0%) | 47 (18.2%) | 156 (30.6%) | 162 (36.5%) | 180 (30.6%) | 140 (38.8%) | 111 (27.2%) |  | |  |
| Higher Education | 328 (12.8%) | 20 (7.8%) | 83 (16.3%) | 44 (9.9%) | 80 (13.6%) | 56 (15.5%) | 45 (11.0%) |  | |  |
| **Number of Sports** |  |  |  |  |  |  |  |  | |  |
| 0 | 1015 (39.5%) | 146 (56.6%) | 253 (49.7%) | 100 (22.5%) | 248 (42.2%) | 51 (14.1%) | 217 (53.2%) | <0.001 | | Chi-Square |
| 1 | 811 (31.6%) | 77 (29.8%) | 116 (22.8%) | 184 (41.4%) | 175 (29.8%) | 144 (39.9%) | 115 (28.2%) |  | |  |
| 2+ | 742 (28.9%) | 35 (13.6%) | 140 (27.5%) | 160 (36.0%) | 165 (28.1%) | 166 (46.0%) | 76 (18.6%) |  | |  |
| **Total PA duration: mean min/day** |  |  |  |  |  |  |  |  | |  |
| Mean (sd) | 199 (111) | 166 (103) | 211 (111) | 193 (107) | 199 (110) | 214 (113) | 196 (116) | <0.001 | | ANOVA |
| **PA component: Duration** |  |  |  |  |  |  |  |  | |  |
| Long duration | 1165 (45.4%) | 84 (32.6%) | 257 (50.5%) | 194 (43.7%) | 262 (44.6%) | 185 (51.2%) | 183 (44.9%) | <0.001 | | Chi-Square |
| Medium duration | 962 (37.5%) | 107 (41.5%) | 184 (36.1%) | 173 (39.0%) | 220 (37.4%) | 137 (38.0%) | 141 (34.6%) |  | |  |
| Short duration | 422 (16.4%) | 64 (24.8%) | 65 (12.8%) | 74 (16.7%) | 98 (16.7%) | 39 (10.8%) | 82 (20.1%) |  | |  |
| **PA component: Muscle Strength** |  |  |  |  |  |  |  |  | |  |
| High Strength | 1025 (39.9%) | 46 (17.8%) | 213 (41.8%) | 141 (31.8%) | 233 (39.6%) | 263 (72.9%) | 129 (31.6%) | <0.001 | | Chi-Square |
| Medium Strength | 915 (35.6%) | 85 (32.9%) | 214 (42.0%) | 186 (41.9%) | 236 (40.1%) | 36 (10.0%) | 158 (38.7%) |  | |  |
| Low Strength | 609 (23.7%) | 124 (48.1%) | 79 (15.5%) | 114 (25.7%) | 111 (18.9%) | 62 (17.2%) | 119 (29.2%) |  | |  |
| **PA component: Intensity** |  |  |  |  |  |  |  |  | |  |
| High Intensity | 1102 (42.9%) | 47 (18.2%) | 207 (40.7%) | 196 (44.1%) | 260 (44.2%) | 275 (76.2%) | 117 (28.7%) | <0.001 | | Chi-Square |
| Medium Intensity | 1053 (41.0%) | 111 (43.0%) | 247 (48.5%) | 184 (41.4%) | 247 (42.0%) | 72 (19.9%) | 192 (47.1%) |  | |  |
| Low Intensity | 394 (15.3%) | 97 (37.6%) | 52 (10.2%) | 61 (13.7%) | 73 (12.4%) | 14 (3.9%) | 97 (23.8%) |  | |  |
| **PA component: Mechanical Strain** |  |  |  |  |  |  |  |  | |  |
| High Mech.Strain | 415 (16.2%) | 34 (13.2%) | 44 (8.6%) | 185 (41.7%) | 98 (16.7%) | 14 (3.9%) | 40 (9.8%) | <0.001 | | Chi-Square |
| Medium Mech.Strain | 1681 (65.5%) | 172 (66.7%) | 389 (76.4%) | 180 (40.5%) | 399 (67.9%) | 241 (66.8%) | 300 (73.5%) |  | |  |
| Low Mech.Strain | 453 (17.6%) | 49 (19.0%) | 73 (14.3%) | 76 (17.1%) | 83 (14.1%) | 106 (29.4%) | 66 (16.2%) |  | |  |
| **PA component: Turning Actions** |  |  |  |  |  |  |  |  | |  |
| High Turning | 558 (21.7%) | 73 (28.3%) | 0 (0%) | 347 (78.2%) | 109 (18.5%) | 0 (0%) | 29 (7.1%) | 0.0085 | | Fisher's exact |
| Medium Turning | 1621 (63.1%) | 167 (64.7%) | 505 (99.2%) | 82 (18.5%) | 467 (79.4%) | 36 (10.0%) | 364 (89.2%) |  | |  |
| Low Turning | 370 (14.4%) | 15 (5.8%) | 1 (0.2%) | 12 (2.7%) | 4 (0.7%) | 325 (90.0%) | 13 (3.2%) |  | |  |
| **Functional limitation scale[0-9]** |  |  |  |  |  |  |  |  | |  |
| Median [Q1, Q3] | 0 [0, 2.00] | 2.50 [0, 6.00] | 0 [0, 1.00] | 0 [0, 2.00] | 0 [0, 1.00] | 0 [0, 0] | 0.500 [0, 3.00] | <0.001 | | Kruskal-Wallis |
| **MMSE-score based on maximum spel/num** |  |  |  |  |  |  |  |  | |  |
| Mean (sd) | 27.4 (2.62) | 25.7 (3.76) | 28.1 (1.91) | 27.4 (2.63) | 27.5 (2.41) | 28.0 (1.89) | 27.0 (2.83) | <0.001 | | ANOVA |
| **Number of Chronic comorbidities** |  |  |  |  |  |  |  |  | |  |
| Median [Q1, Q3] | 2.00 [1.00, 3.00] | 2.00 [1.00, 3.00] | 1.00 [1.00, 2.00] | 2.00 [1.00, 3.00] | 2.00 [1.00, 3.00] | 1.00 [0, 2.00] | 2.00 [1.00, 3.00] | <0.001 | | Kruskal-Wallis |
| **Health problems limit normal activities** |  |  |  |  |  |  |  |  | |  |
| Yes, severely | 275 (10.7%) | 49 (19.0%) | 33 (6.5%) | 57 (12.8%) | 51 (8.7%) | 28 (7.8%) | 57 (14.0%) | <0.001 | | Chi-Square |
| Yes, slightly | 581 (22.6%) | 79 (30.6%) | 114 (22.4%) | 94 (21.2%) | 130 (22.1%) | 72 (19.9%) | 92 (22.5%) |  | |  |
| No | 1707 (66.5%) | 129 (50.0%) | 361 (70.9%) | 291 (65.5%) | 406 (69.0%) | 261 (72.3%) | 259 (63.5%) |  | |  |
| **Self-perceived health** |  |  |  |  |  |  |  |  | |  |
| Excellent | 347 (13.5%) | 11 (4.3%) | 88 (17.3%) | 58 (13.1%) | 69 (11.7%) | 64 (17.7%) | 57 (14.0%) | 0.0085 | | Fisher's exact |
| Good | 1247 (48.6%) | 110 (42.6%) | 244 (47.9%) | 212 (47.7%) | 322 (54.8%) | 190 (52.6%) | 169 (41.4%) |  | |  |
| Fair | 601 (23.4%) | 90 (34.9%) | 122 (24.0%) | 111 (25.0%) | 110 (18.7%) | 65 (18.0%) | 103 (25.2%) |  | |  |
| Sometimes good/bad | 309 (12.0%) | 33 (12.8%) | 46 (9.0%) | 52 (11.7%) | 74 (12.6%) | 36 (10.0%) | 68 (16.7%) |  | |  |
| Poor | 56 (2.2%) | 14 (5.4%) | 9 (1.8%) | 9 (2.0%) | 11 (1.9%) | 3 (0.8%) | 10 (2.5%) |  | |  |
